# Long-read metagenomics and methylation-based binning allow the description of the emerging high-risk antibiotic resistance genes and their hidden hosts in complex communities

**DOI:** 10.64898/2026.02.18.26346558

**Authors:** Melina Markkanen, Heidi Putkuri, Dovydas Kičiatovas, Ville Mustonen, Marko Virta, Antti Karkman

## Abstract

Antibiotic resistance genes (ARGs) circulating among clinically relevant bacteria pose serious challenges to public health. Given the ancient and environmental bacterial origins of ARGs, a better understanding of the carriers of ARGs beyond the clinically most relevant species is urgently needed for more farsighted resistance monitoring and intervention measures. While the risks of emerging ARGs from environmental sources have been recognized, the identification bottlenecks stem from the limitations of shotgun metagenomic sequencing and bioinformatic methods. Here, we used long-read metagenomic sequencing and bacteria-specific methylation profiles to re-establish the links between established (well-described) or latent (absent in databases) ARGs and their bacterial and genetic contexts in wastewater. The base modification data produced by PacBio SMRT sequencing was analyzed by an in-house pipeline utilizing position weight matrices and UMAP visualizations. The approach was validated by a synthetic community with known bacterial composition. Our analysis revealed several previously unreported ARGs and their hosts with varying risk levels defined by their potential as emerging public health threats. For instance, *Arcobacter*, as one of the prevalent taxa in influent wastewater, was shown to carry a latent beta-lactamase gene with high predicted mobility potential. Of the other emerging beta-lactamases, we provided a real-life example of ongoing p*dif* module-mediated genetic reshuffling of the *bla*MCA gene occurring at least within *Acinetobacter* hosts in our samples. Additionally, we identified *Simplicispira*, Phycisphaerae, and environmental groups of the Bacteroidales order as the carriers of established, clinically important ARGs. These findings support the intermediate host roles of strictly environmental bacteria for the further dissemination of mobilized ARGs, highlighting the importance of exploring the uncultivated, or non-pathogenic, carriers of ARGs for the early detection of newly arising ARGs and mobility mechanisms.

## Introduction

The introduction of antibiotics into therapeutic practice represents a pivotal milestone in the advancement of medicine (1). However, the simultaneous increase in emergence and circulation of antibiotic resistance genes (ARGs) has severely diminished the efficacy of the antibiotic drugs available in human and veterinary medicine, challenging the entire health care system. According to the most recent estimates, deaths of 4.71 million people alone in 2021 were associated with bacterial resistance, whereas for the year 2050, the estimate stands at 8.2 million (2). Although the ARGs are ancient components of some bacterial genomes owing to the “producer hypothesis” (3–5), their rapid increase and spread across bacterial lineages are driven by the gene mobilization events occurring between bacteria, which is fueled by the selection pressures arising from the clinical use of antibiotics (6). ARG mobilization is enabled by horizontally transferred mobile genetic elements (MGEs) and their adjacent genes. Sequential ARG mobilization events, starting from the source organism and proceeding further toward the putative opportunistic pathogen hosts, may include several steps of so-called intermediate carriers of ARGs (7, 8).

The resistance features carried by the most common opportunistic pathogens are relatively well described owing to their immediate clinical relevance, but also due to practical reasons, as many of these bacteria are cultivable. However, given the complexity of the resistance gene dynamics – including non-pathogenic bacteria – and rapidly evolving resistance situation, a better understanding of the ARG mobility mechanisms and ARG carriers beyond the cultivable clinical strains is urgently needed (7). The study of environmental resistomes enables the identification of both established and latent ARGs – that is, those that are well-characterized in databases and those that are largely unexplored – regardless of the cultivability of their host bacteria (9).

For instance, both established and predicted versatile beta-lactamase genes are highly frequent among environmental bacteria, especially within wastewater communities (5, 9, 10). Indeed, wastewater has been identified as one of the key environments regarding the presence of latent ARGs (9) with elevated risk of becoming clinically problematic if mobilized into relevant opportunistic pathogen hosts (8, 9). This is not difficult to envision due to the high diversity of bacteria present in wastewater originating from both the human body and various environmental sources, striving to persist despite a range of environmental stressors (11). Newly emerging ARGs are expected to arise from the largely unknown repertoire of bacteria inhabiting external environments and commonly referred to as environmental bacteria (8, 12), whose description and detection are still limited in contrast to cultivable human health-related species. The transition from a non-mobile context in environmental bacteria to clinically relevant ARGs circulating among human pathogens contains multiple gene decontextualization and host transition events by intermediate hosts (7, 8). Unfortunately, considering the prevailing conditions of the post-antibiotic era, this transfer may be irreversible (13). Several environmental origin bacteria and intermediate hosts (or recent hosts) of the current clinical ARGs have been resolved (3, 5, 6, 14–16). Although due to detection difficulties (7), this is typically done retrospectively. Knowledge about the intermediate ARG carriers is particularly important, as they may supply the foundational genetic material for the emergence of clinically relevant resistant bacteria (17).

Ranking ARGs in environmental resistomes by their impact on human health is inherently difficult and remains debatable (12, 18). The risk–ranking schemes that have given prominence to mobile ARGs that are already present in pathogens (18, 19) have been challenged by more farsighted frameworks, which define novel resistance determinants arising from environmental microbiomes, especially with the implications of gene mobility, as the most critical contributors to the current rapidly developing resistance problem. These schemes aim for early capture of emerging resistance threats by assessing the likelihood of ARG transmission from environmental species to clinically relevant bacteria and the severity of such an event (12), using ARG mobility as the proxy for this (7).

However, the inability of shotgun metagenomic sequencing (20) and assembly (21) in linking the ARGs, especially those encoded by MGEs, to their host bacteria has substantially limited our understanding of the non-pathogenic or environmental carriers of ARGs and has hindered the risk assessment (7, 12). Even though metagenomic assembly with long-reads may allow simultaneous identification of ARGs and their near genetic context, their hosts remain unresolved, especially in highly complex communities, such as influent wastewater (7). The current metagenomic binning algorithms, relying on sequence composition and coverage, tend to overlook the ARGs residing within MGEs, for instance, by the inability to connect plasmids to their carrier bacteria due to differential coverage (22, 23). This remains one of the most significant challenges in the entire field of microbial metagenomics and antibiotic resistance research, as plasmids carry vital adaptation-increasing and evolution-driving features (23, 24). Additionally, binning algorithms are confused by the presence of multiple strains of one species, biased toward the high-abundance taxa while ignoring the genomes of rare species, which the ARG-carrying bacteria commonly are (9, 25). Consequently, these biases leave us with persistent uncertainties regarding the role of environmental lineages on the ARG carriage and spread.

Bacterial DNA methylation signals are base modifications mediated by target sequence motif-specific methyltransferase enzymes, often acting as part of the restriction-modification (R-M) systems (23). As the main epigenetic modification mechanism of bacterial genomes, methylation has roles in various regulatory functions, gene expression (26, 27), and genome defense against phages (28) by enabling the distinction between own and foreign DNA (27). Due to the unique set of methylation-mediating enzymes and their unique target motifs, all sequence material of a bacterial strain (chromosomes and plasmids) is expected to show strain-specific methylation profiles (23), allowing their bioinformatic reconnection into bins of connected contigs (14, 23). So far, the bacterial methylation detection with the latest technology by PacBio and Nanopore, allowing direct methylation detection without pretreatment (27), has been focused on isolates, while fewer analysis tools and workflows are available for metagenomic datasets, except those with low bacterial complexity (23, 29–32). For instance, the computational demands required for a comprehensive analysis of highly complex wastewater microbial communities with the MultiMotifMaker algorithm (33) extend beyond what is practically achievable. Tools similar to Nanomotif (22), which is specifically designed for Nanopore data, are greatly needed to take advantage of the kinetic data produced by PacBio SMRT sequencing.

Here, we present a pipeline for summarizing the PacBio base modification data using position weight matrices (PWMs) and Uniform Manifold Approximation and Projection (UMAP) to cluster metagenomic contigs based on their methylation profiles to generate bins of connected contigs (or “genome bins” hereafter) and thereby reveal the ARG-carrying bacteria present in wastewater. A synthetic community with known bacterial composition and available sequence data was used to validate our methodology. In contrast to individual metagenomic contigs, the ability to form genome bins, even in incomplete form, provides sufficient genomic context to support predictions of ARG mobility and host species assignment. We focused on carriers of both established and latent ARGs detected in influent, effluent, and dried sludge to expand our understanding of the (hidden) environmental bacterial diversity responsible for the carriage and spread of ARG, as well as the hosts of uncharacterized ARGs showing potential for transmission to clinically relevant bacteria.

## Materials and Methods

### Sample collection and DNA extraction

Detailed sample descriptions of the wastewater metagenomes (PRJEB106782, PRJEB83306) investigated here are provided in our previous work (14). Briefly, composite samples were collected from influent (INF1, INF2, INF3), effluent (EFF1, EFF2, EFF3), and dried sludge (SLU1, SLU2, SLU3) at one of the two wastewater treatment plants responsible for processing wastewater in the Helsinki metropolitan area in Finland during three days in 2019 (18.3.2019, 20.3.2019, 26.3.2019). For wastewater samples, after filtration through a 0.22 µm membrane, the DNA was extracted from the filter using the Qiagen DNeasy PowerWater kit with bead beating.

Synthetic community data (PRJEB85727) of the study by Partanen and colleagues (34) and associated genome sequence data (PRJNA1047486) by Hogle and colleagues (35) were utilized as the validation data for the correctly reconnected metagenomic contigs. A comprehensive description of the synthetic communities is provided in the work by Partanen (34) and Hogle and colleagues (35) and **Supplementary Methods**.

### Sequencing and preparatory analyses

Library preparation and PacBio Single Molecule Real-Time (SMRT) sequencing for metagenomic long-reads of wastewater communities were performed at the Institute of Biotechnology, University of Helsinki. Highly accurate long reads (HiFi reads) of metagenomic community samples with kinetic tags were generated from the subreads using PacBio BAM toolkit programs (https://github.com/PacificBiosciences/pbtk) to allow methylation detection in downstream analyses. Instead, HiFi reads without the tags were used for metagenomic assembly. Each of the synthetic and wastewater community samples (five and nine samples, respectively) was assembled separately using hifiasm-meta (v.0.18.0) (36). Detailed Snakemake (v7.17.1) (37) workflows are found in https://github.com/melinamarkkanen/Methylation.

### Primary data analyses of the metagenomic contigs

HiFi reads with attached kinetic tags were aligned to assembled metagenomic contigs using the PacBio program pbmm2 (v1.10) (https://github.com/PacificBiosciences/pbmm2/). Comparison of the observed interpulse duration (IPD) values of each sequence position from aligned reads to those expected for unmodified sequences allowed the detection of base modifications by the ipdSummary program (v.3.0) (38). As a result, statistically significant comparisons are gathered as rows into the resulting General Feature Format (GFF) output files for all contigs separately. Base modifications are identified as either of types m6A or m4C or of unknown type, referred to as ‘modified base’. Along with the contig name and modification type, the position of the methylated base and its flanking region of 20 bp upstream and downstream is recorded for the following steps.

### Methylation data conversion to Position Weight Matrices (PWM)

The obtained contig-wise summaries of the detected methylated bases were converted into Position Weight Matrices (PWM) to be used as unique signatures for further analysis. For that, in-house Python scripts were used to extract the flanking sequences separately for each modification type and sample (5 and 9 synthetic and wastewater community samples, respectively) and then convert those data into PWMs. In the PWM files, each row represented one nucleotide (A, T, G, or C) and a column indicated each position surrounding the methylated base [−20, 20] (more detailed description of the constructed PWMs is provided in the Supplementary Methods). To discover the balance between noise and an adequate amount of data (given the high complexity of wastewater samples, especially influent) for downstream methylation analysis, the minimum thresholds for detected instances of modified bases per contig were explored with the data of known bacterial composition (**Table S1**). Of the tested values (20, 50, 100, 200, and 500), 50 was selected as it provided the highest model accuracy values for the Random Forest classifier (**Table S2**).

### Random Forest classifier

To test our methylation-based approach and the high-dimensional data presentation using PWMs described above, a Random Forest model was trained and implemented using Scikit-learn (v1.4.2) (39) Python library on synthetic community data with *a priori* known taxonomical composition (**Supplementary Methods**). Random Forest model was chosen due to its ability to generate feature importance scores, offering insights into the relative contribution of individual methylation types, positions, and bases to the prediction of the origin of bacterial genomes of metagenomic contigs. Model performance was evaluated using a confusion matrix, which summarizes the number of correct and incorrect predictions. Overall accuracy was calculated as the proportion of correctly classified instances to the total number of instances.

### Visualization of PWMs with the synthetic community data

UMAP (Uniform Manifold Approximation and Projection) (40) was applied to visualize and reduce the complexity of the high-dimensional data by projecting it into two dimensions. By the non-linear dimensionality reduction technique of UMAP, known for its ability to preserve both local and global structures in data (40), this approach allowed us to explore the clusters represented by methylation profiles. In addition to visual inspection of the clustering of species-wise colored contigs in UMAP, contigs clustering together were combined into (sample-wise) bins of connected contigs to confirm their completeness and putative redundancy, which were evaluated by CheckM2 (v 1.0.1) (41). GTDB-Tk (v2.4.1, database release 220) (42) was used to confirm the taxonomical identity of the genome bins. We also investigated whether contigs within the same clusters exhibit similar nucleotide information content by generating sequence logos showing the nucleotide preferences across the context sequences at each position (43), which, in our case, illustrates the methylation target motif sequences (**Fig. S4**).

### Identification of established and latent ARG-carrying genome bins in wastewater

Established and latent ARGs present in the contigs were predicted using ResFinder (v 4.7.0) (44) and fARGene (Fragmented Antibiotic Resistance Gene iENntifiEr) (v0.1) (45), respectively. By color-coding the contigs with ARGs in the UMAP visualization, we directed the focus on the formation of genome bins with ARGs. The need for manual processing in methylation-based binning restricted the volume of data that could be examined. This was especially highlighted regarding the latent ARGs detected by fARGene, with which we decided to focus on the putative beta-lactamases due to their higher likelihood of serving the antibiotic resistance function and thus being mobile in contrast to chromosomal housekeeping genes that are incorrectly assigned as putative resistance genes (10). The clustering contigs were combined similarly to the synthetic community data, and the quality and the taxonomical identity of the resulting genome bins were evaluated similarly as described for the synthetic community above.

### Analysis of ARG mobility potential and genetic context

Five bins of connected contigs with established or latent ARGs were investigated in more detail as example cases representing newly resolved ARG-host linkages. These genome bins were determined using evidence from the initial exploratory phase: The established or latent ARG was detected in multiple different contigs and potentially hosted by different species, indicated by its location in separate clusters of the UMAP visualization (**Fig. S8**).

The following genetic context analysis was performed for all five examined bins of connected contigs with ARGs: In addition to the ARG present in the binned contigs, hits to the same ARGs were queried among all wastewater data using BLAST (46). ARG flanking regions (5 or 10 kb up-and downstream of the ARG) of the ARG of the matching sequences (including both those binned using methylation and those that remained unbinned due to scarce or absent methylation data) were extracted using an in-house script applying SeqKit tools (v 2.5.1) (47). Additional mobility predictions for the entire contigs or extracted flanking regions, respectively, were performed using geNomad (v1.8.0) (48) and pdifFinder (49). Bakta (v1.11.0) (50) was used for annotating the genes in the flanking regions. To examine the diversity within the genetic contexts and to reduce the number of sequences for visualization purposes, the genetic contexts were dereplicated to exclude identical or similar genetic contexts by vsearch (v2.22.1) (51). After removing the redundancy of the contexts, the ARGs in those that remained were extracted for phylogenetic tree analysis. For that, gene alignment was done using MAFFT (v7.505) (52), for which the tree was computed by RAxML (command raxmlHPC-PTHREADS) (v8.2.12) (53) and visualized with Interactive Tree of Life (iTOL) (54).

The flanking sequences were visualized using pyGenomeViz (v1.6.1) (55) using a BLAST (46) query for highlighting the aligned nucleotide sequences between the compared sequences. The insertion sequences located near the investigated ARGs were examined in more detail by the ISfinder (56) online server and the *attC* sites by the HattCI program (57). Protein structures of the *Arcobacter* encoded latent beta-lactamase genes were predicted using AlphaFold (58), and similar structures were queried by FoldSeek (59).

### Taxonomical composition

Relative abundance of genus-level taxa in both synthetic and wastewater communities was analyzed using Sylph (sylph (v 0.8.1) and sylph-tax (v 1.5.1)), with metagenomic reads as the input (60). For the wastewater data, genera with relative abundance below 0.5 % were left out of the visualization.

## Further details

All other figures that were not specifically mentioned above were drawn using Plotly (61) and RStudio (2024.12.1+563.pro5) (62) by applying packages phyloseq (v1.16.2) (63), ggplot2 (v3.5.0) (64), and vegan (v2.6-10) (65). Comprehensive descriptions of the materials and methods can be found in the **Supplementary Methods**.

## Data availability

The subread sequence data from the wastewater metagenomes are available in the European Nucleotide Archive (ENA) under accession number PRJEB106782, along with the corresponding HiFi reads, which can be found under accession number PRJEB83306. The other sequencing data used in this study were accessed via the following accession numbers: synthetic community metagenomes PRJEB85727 (34) and the whole genome sequence (WGS) data of the synthetic communities PRJNA1047486 (35) (**Table S1**). The genome bin sequences are available in https://doi.org/10.5281/zenodo.18239683. The analysis scripts are shared in the publicly available GitHub repository at https://github.com/melinamarkkanen/Methylation.

## Results

### Position Weight Matrix (PWM) -based method for linking contigs by their shared methylation signatures

In the absence of suitable tools for methylome analysis of PacBio SMRT sequencing data from complex microbial community samples, such as wastewater, a new workflow was established here (**Fig. 1**). In our approach, the per-contig base modifications were converted into position weight matrices (PWMs), allowing their clustering according to their origin bacterial genome by Uniform Manifold Approximation and Projection (UMAP) to form bins of connected contigs, relying on the assumption that methylation profiles are unique among bacterial strains. We validated our workflow with a synthetic community of known bacterial composition, of which whole genome sequence (WGS) data were available for half of the community members (**Table S1**, **Fig. S1**). Moreover, preliminary analyses with the validation data enabled the exploration of the most appropriate signal-to-noise ratio regarding the amount of the input methylation data (>50 base modifications per contig) for the downstream Random Forest analysis (**Table S2**).

**Fig. 1.**
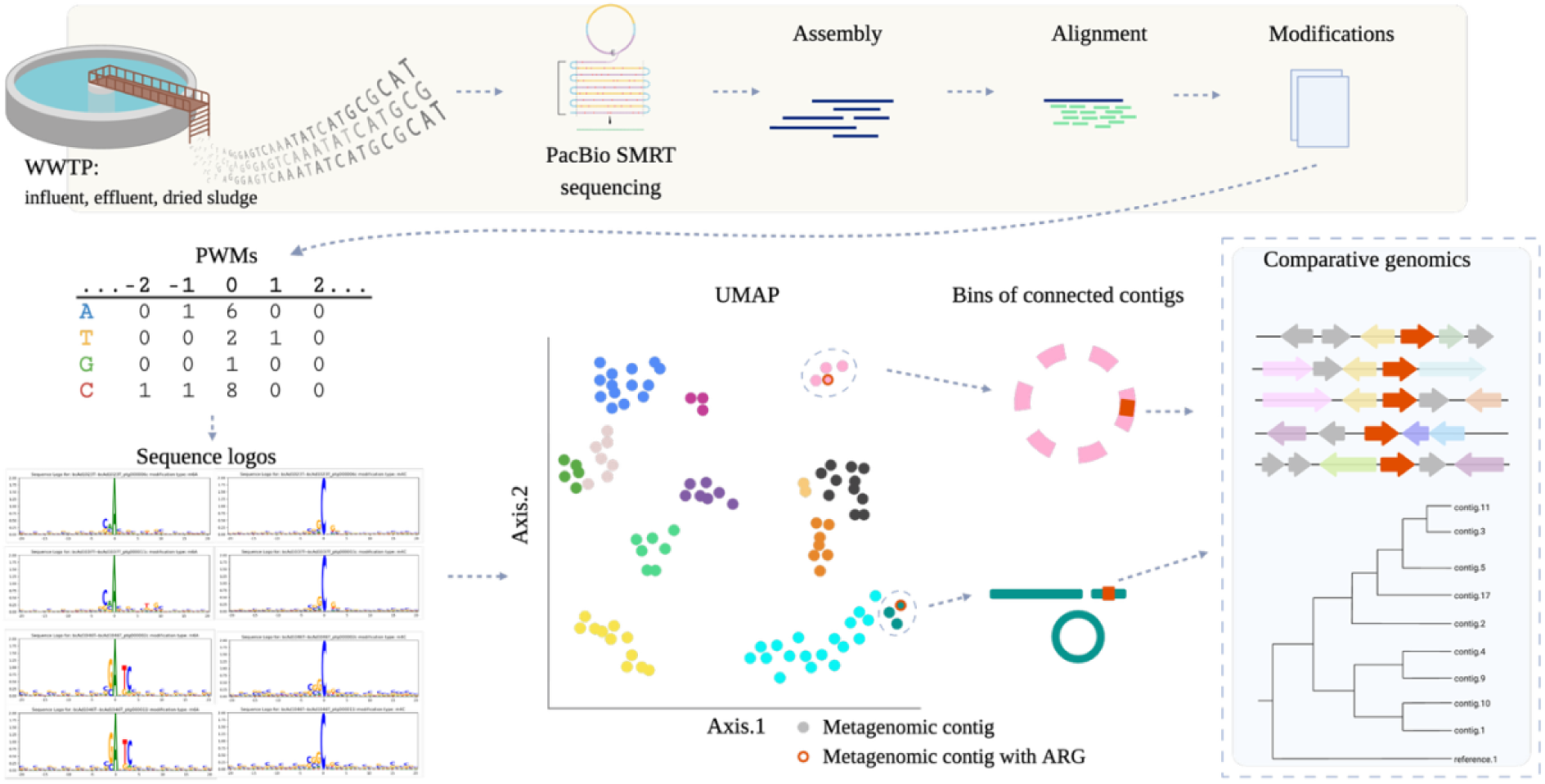
Schematic representation of the key steps in the workflow. Before the analysis of the wastewater samples, the workflow (until the formation of bins of connected contigs step) was applied on a synthetic community with known bacterial composition as a method validation procedure. Figure created with BioRender.com.

The validation results confirmed that, at least for most community members, the species identity of a metagenomic contig can be predicted based on its methylation profile presented by PWMs (**Fig. S2**, **Table S2**). This was confirmed by the Random Forest confusion matrix, which summarized true and false predictions and provided the basis for calculating test accuracy. The model showed strong performance, with a test accuracy value of 0.88 at the species level (**Table S2**). For the remaining cases, prediction accuracy was low due to misclassifications across many different species, rather than repeated specific incorrect predictions, suggesting rather heterogeneous and inconclusive methylation profiles instead of substantially overlapping profiles between multiple species (**Fig. S2, Table S2**). This was reflected in the UMAP visualization as a cloud of contigs that were not unique to any single species (**Fig. S3A**). However, as this miscellaneous group of contigs was clearly separate from the more defined, distinct clusters, the risk of false positives coming from this group is diminished (loss of data rather than false positives). In some but not all cases, a higher amount of input methylation data or the contig length seemed to be descriptive for the more distinct species-wise clusters (**Fig. S3B**).

Metagenomic contigs from synthetic communities were clustered based on their methylation profiles, aiming to recover contig relationships lost during shotgun sequencing. To test whether the contig clusters correspond to the original community member genomes, completion and redundancy values, computed by CheckM2 (v 1.0.1) (41), of some selected species metagenome-assembled genomes (MAGs) or bins of connected contigs were examined (**Table S3**). Fully complete MAGs containing both chromosomal and plasmid sequences were resolved for multiple species and samples (**Table S3**). This was also seen in the sequence logos of chromosomal and plasmid contigs showing ± 20 bases surrounding the modified base, differentially sized according to their information content (**Fig. S4**). However, in some cases, only partial genome bins could be curated due to too few methylation information observed as the absence of contigs (exclusion by initial filtering by >50 base modifications/contig) or located apart from the main cluster due to incomplete methylation profile (**Fig. 3A**, **Table S3**). Altogether, we concluded that our approach could reconnect metagenomic contigs into species-wise bins of connected contigs to allow the establishment of ARG-host linkages and description of previously unknown contributors to resistance carriage in wastewater.

### General overview of the wastewater bins of connected contigs with ARGs

The miscellaneous clusters of metagenomic contigs in UMAP visualizations of wastewater samples were excluded to decrease noise and obtain a more detailed view of distinctive clusters for the manual bin curation step (**Fig. S7**). Aiming to re-establish the links between resistance genes and their wider contexts, we focused on clusters containing contigs with established or latent ARGs (**Fig. S8**). After quality assessment, 35 genome bins with established (**Table 1**) and 53 with latent (**Table 1**) ARGs were described. The average completion and redundancy values were 55.40 and 2.17, respectively. Of all genome bins, 31 were obtained from influent, 15 from effluent, and 42 from dried sludge (**Table 1**). However, due to the non-quantitative nature of the approach and the included manual steps, the resulting bins of connected contigs cannot be considered a quantitative measure of ARG-encoding bacteria in the samples. On average, the genome bins were composed of 12 contigs (range from 2 to 44) (**Table 1**). In addition, the GC-content of the genome bin corresponded to that of the bin’s assigned taxa and their closest matching reference genome by GTDB-tk (**Table 1**).

**Table 1.**
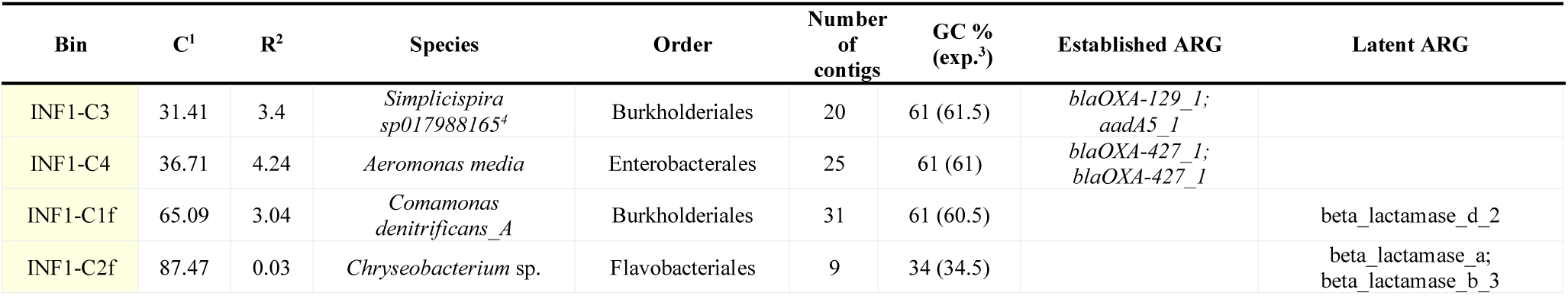
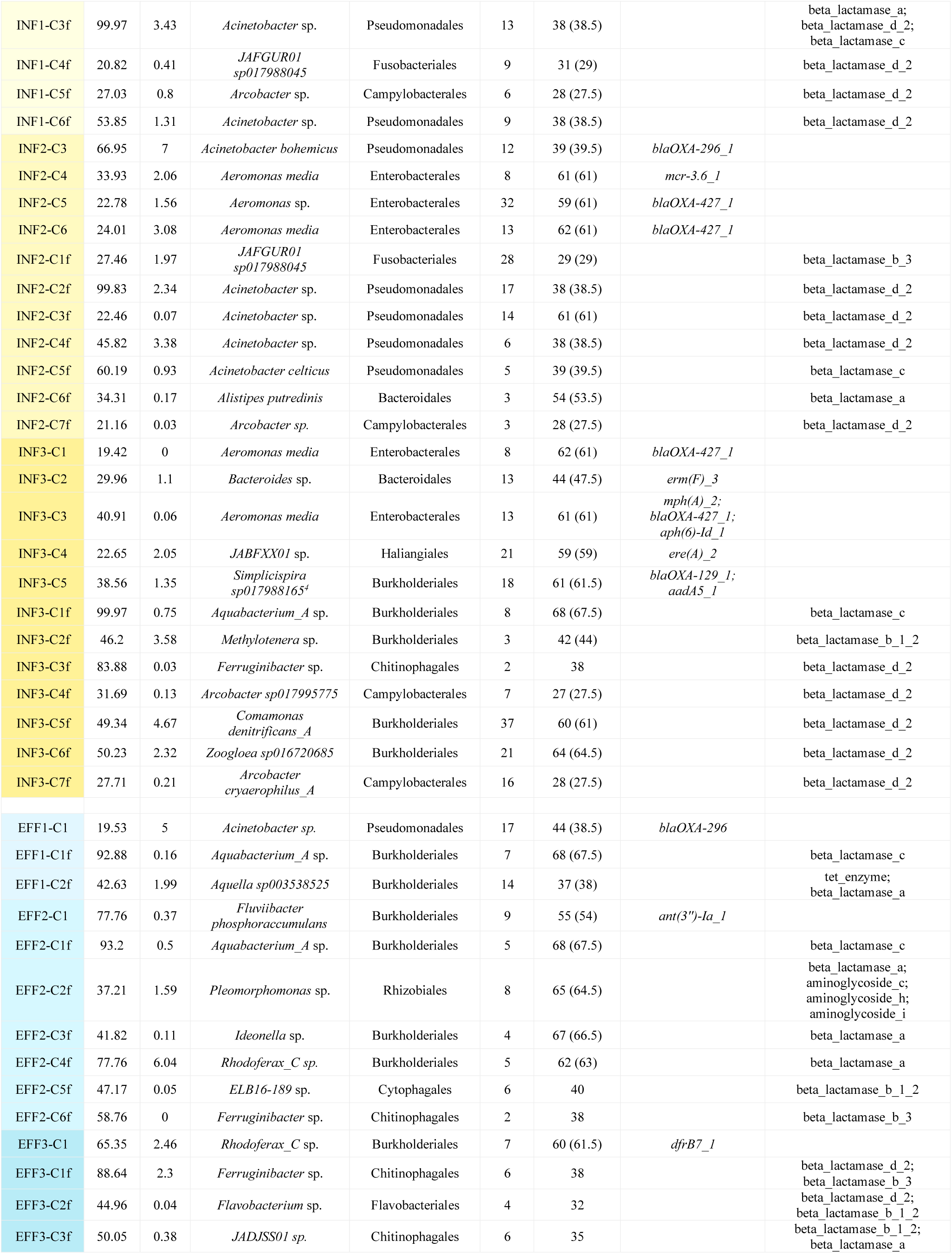
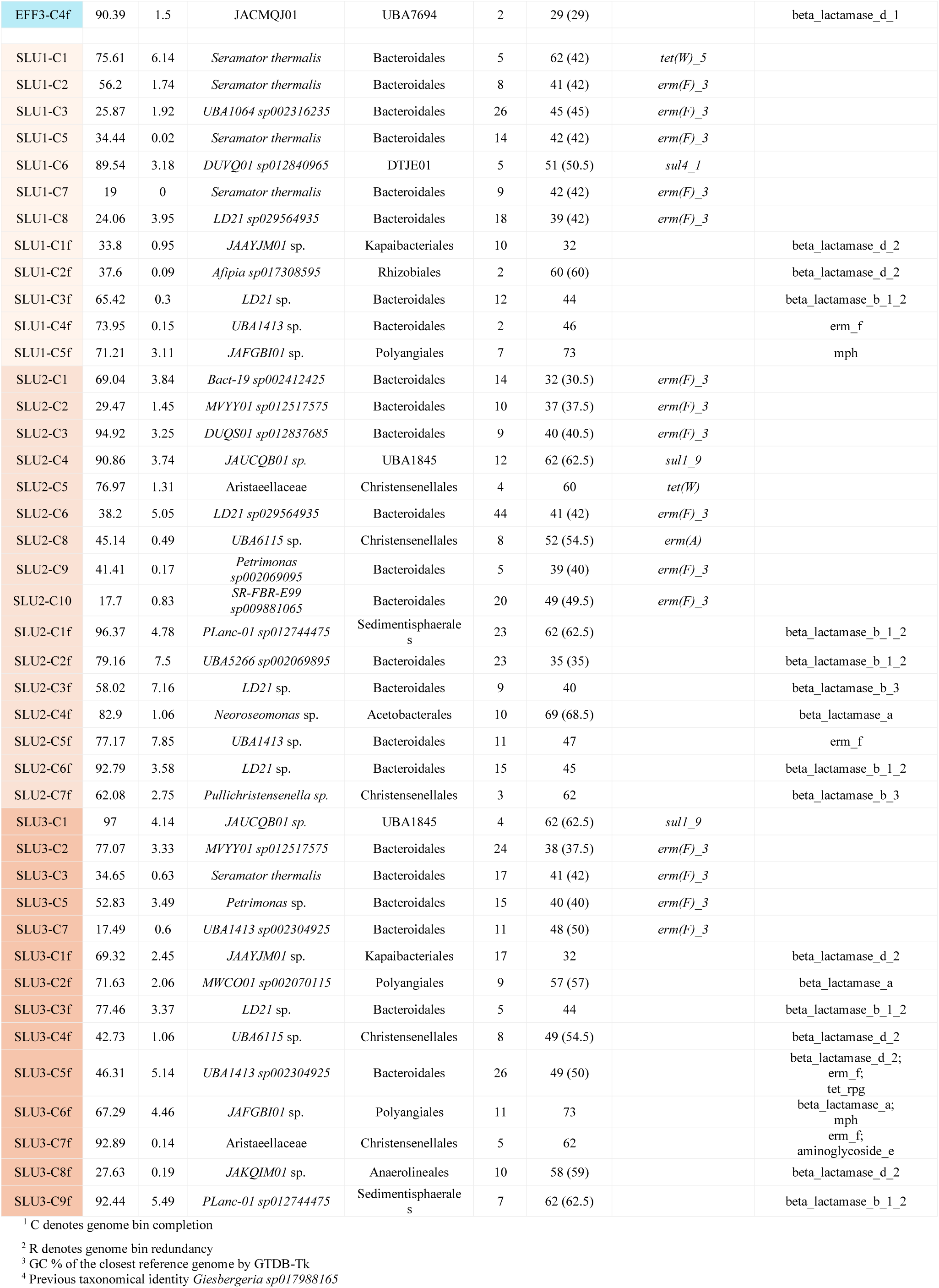
Methylation-based bins of connected contigs encoding latent and established ARGs.

Given the pivotal role of plasmid-encoded ARGs for the antibiotic resistance problem and the difficulties related to their characterization within whole-community data, we were particularly interested in exploring the plasmid sequences that were linked to their host genome bins by the methylation analysis. We were not able to link any ARG-encoding plasmids to their wider, genome bin context (**Table 1**). However, we could find linkages between non-ARG-carrying plasmids and their host genome bins in several cases (**Table S4**). For instance, two to five plasmids were linked to *Acinetobacter* sp. in three different genome bins from influent (bins INF1-C3f, INF1-C6f, and INF2-C2f) (**Table S4**). Furthermore, single plasmids were identified, for example, for *Aquabacterium_A* sp. (bin EFF2-C1f) and *Aeromonas media* (bin INF1-C4) (**Table S4**). Querying these contigs against the NCBI database with BLAST resulted in matches with plasmid sequences (data not shown), confirming our ability to connect chromosomal and plasmid sequences.

*bla*OXA genes carrying *Aeromonas* sp. were most commonly identified as established ARG carriers in influent, while Bacteroidales order bacteria carrying *erm*(F) genes were descriptive for dried sludge. The fewer genome bins with established ARGs obtained from effluent were mainly Burkholderia order bacteria carrying various established ARGs. Regarding latent ARGs, beta-lactamases of classes A, B 1-3, C, and D2, most notably of the latter, were identified in more diverse hosts, including bacteria belonging to 16 different orders such as Burkholderiales, Bacteroidales, Pseudomonadales, Chitinophagales, Campylobacterales, and Christensenellales, to name the most common groups. In general, the latent ARG host species included more members of the uncultivated bacterial lineages compared to the identified carriers of established ARGs. We next focused on a couple of the most interesting newly identified carriers of established and latent ARGs (**Table 1**).

### A subset of the latent class D beta-lactamase variants encoded by *Arcobacter* exhibited high mobility potential

*Arcobacter* species of the Campylobacterota phylum were one of the major bacterial groups detected in influent wastewater (**Fig. S6**). Bins of connected contigs (n = 6, 3, 7, 16) assigned as *Arcobacter* spp. and hosting latent class D beta-lactamase genes were recovered from each of the influent samples (bins INF1-C5f, INF2-C7f, INF3-C4f, and INF3-C7f, **Table 1**). The genome bins showed completeness and redundancy values ranging from 21.16 to 31.69 and 0.03 to 0.8, respectively, and they were assigned as *Arcobacter* sp., *Arcobacter* sp., *Arcobacter sp017995775*, and *Arcobacter cryaerophilus A,* respectively (**Table 1**). (The previously distinct genera *Arcobacter* and *Aliarcobacter* are referred here as a single genus *Arcobacter* according to the latest update on nomenclature and taxonomical classification (66).) Identical or similar ARG sequences were also found among unbinned contigs of influent wastewater, and these were included in the gene tree and the genetic context analysis, along with the reference sequence *bla*OXA-464, showing the closest similarity of the established beta-lactamase genes (**Fig. 2**, **Table S5**).

**Fig. 2.**
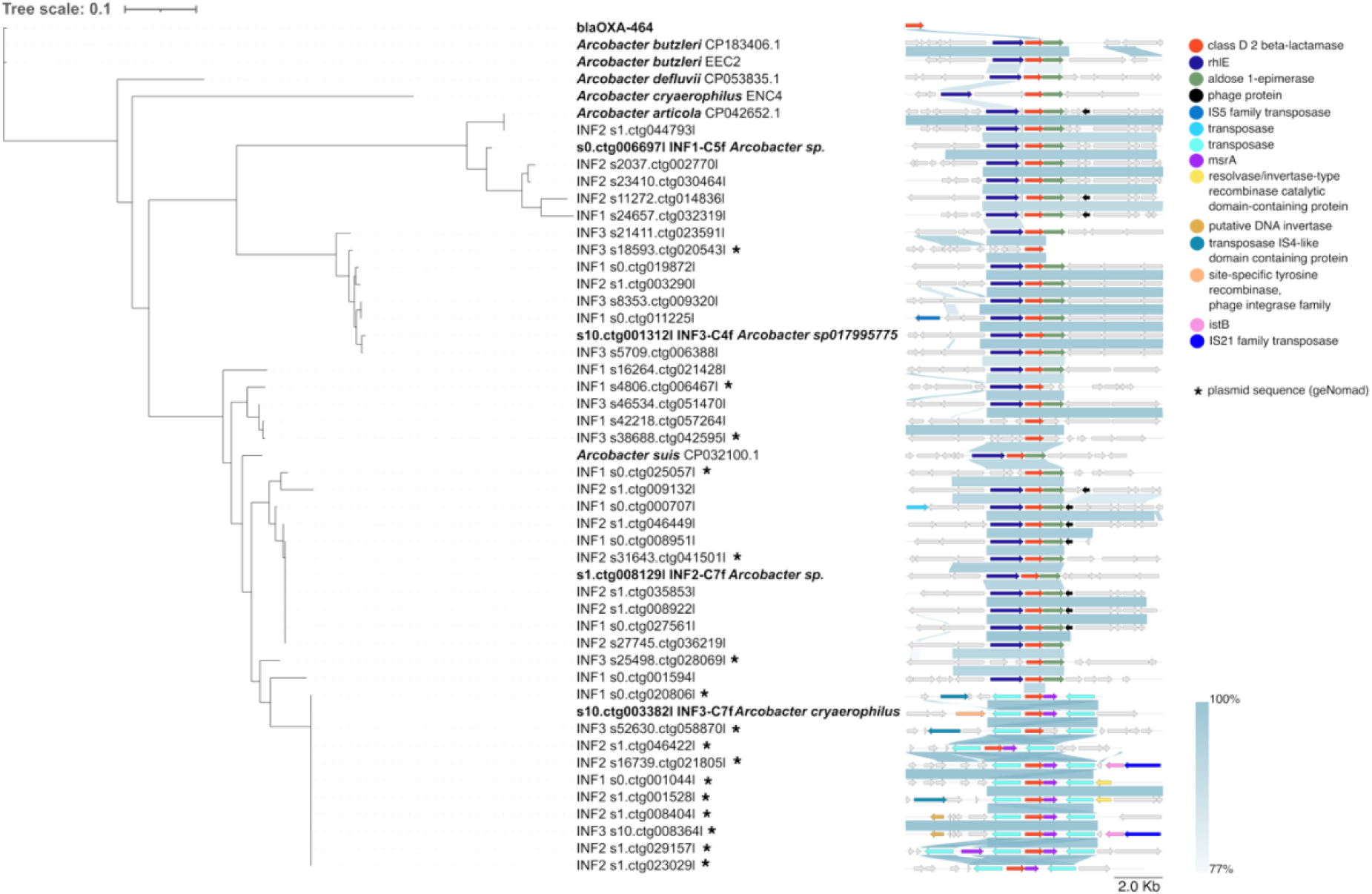
Gene phylogeny tree of the latent class D beta-lactamase gene (shown in red) found in the bins of connected contigs of *Arcobacter* species and unbinned sequences from influent wastewater. Flanking regions of 5 kb upstream and downstream of the class D beta-lactamase were included in the genetic context visualization. Reference sequences of class D beta-lactamase and their contexts were embedded in the analysis.

The class D beta-lactamases carried by each of the four genome bins formed distinct branches of the tree (**Fig. 2**). Moreover, they primarily differed from those of the reference strains. Of these, *A. butzleri* carried a gene identical to the variant *bla*OXA-464, while the beta-lactamases of *A. defluvii*, *A. cryaerophilus*, and *A. articola* showed more dissimilarities to any other variants (**Fig. 2**). On the contrary, the beta-lactamase encoded by *A. suis* showed the closest similarity to that in one of our genome bins (**Fig. 2**). Despite differences in the beta-lactamase sequences, their immediate flanking region showed substantial conservation across most hosts and branches of the phylogenetic tree (**Fig. 2**, **Table 1**). The conserved region consisted of genes *rhl*E and aldose 1-epimerase (mutarotase), encoding core functions related to RNA (67) and carbohydrate metabolism (68), respectively, further supporting the non-mobile context of the beta-lactamase gene among these sequences (**Fig. 2**).

As a notable exception to the sequences described above, the branch represented by the INF3-C7f genome bin assigned as *A. cryarophilus*, harbored identical beta-lactamase genes but lacked the conserved core region descriptive for all other branches (**Fig. 2**). Instead, the beta-lactamase gene, together with peptide methionine sulfoxide reductase gene *msrA* involved in oxidative stress protection (69) (not to be mixed with the established ARG *msr*(A)) were flanked by transposase genes on both sides (**Fig. 2**). Additionally, IS21 family transposases were detected in some of these contexts within this branch, but not among the other branches (**Fig. 2**). While no similar context sequences of the entire module were detected in public sequence repositories, matching transposase genes were found in various Campylobacterales such as *Sulfurospirillum* and *Arcobacter* species. Furthermore, all except the bin INF3-C7f-linked sequences of the branch were predicted as plasmid-borne, implying higher mobility potential of these sequences, unlike most of the conserved contexts (**Fig. 2**, **Table S5**).

The amino-acid sequence of this beta-lactamase showed low sequence similarity with *bla*OXA-464 (72 % 182/254 AA) and the best matches for its predicted protein structure were the beta-lactamases present in *Sulfurospirillum* sp. hDNRA2 (AF-A0AAU8L8S4-F1, e-value 8.27e-33; sequence identity 50.9%) by AlphaFold Database clustered to 50% sequence identity and *Enterobacter cloacae* (9h15_A_1 e-value 1.33e-29; sequence identity 43.8%) by the Protein Data Bank.

Altogether, these findings suggested that this particular class D beta-lactamase represented a putative novel mobile ARG carried and possibly horizontally transferred at least among *Arcobacter* and other Campylobacterales.

### Class C beta–lactamase genes (*bla*MCA) are reshuffled by p*dif* modules across at least plasmids and chromosomes of *Acinetobacter* species

One class C beta-lactamase gene-carrying, high-quality (99.97 completeness and 3.43 redundancy) *Acinetobacter* sp. genome bin INF1-C3f was characterized through the methylation analysis (**Fig. 3A**, **Table 1**, **Table S6**). This beta-lactamase was predicted by the fARGene tool, which excludes the established ARGs from further analysis steps (45), suggesting that the gene is previously undescribed. However, it was identified in one database, the AMRFinderPlus of the NCBI applied by Bakta (50), where it appeared under the name *bla*MCA. We thus refer to the gene by the name ‘*bla*MCA’ from hereon. Matching genes were found in unbinned contigs from influent samples and were predominantly predicted to be plasmid-derived (**Fig. 3A**, **Table S6**).

**Fig. 3.**
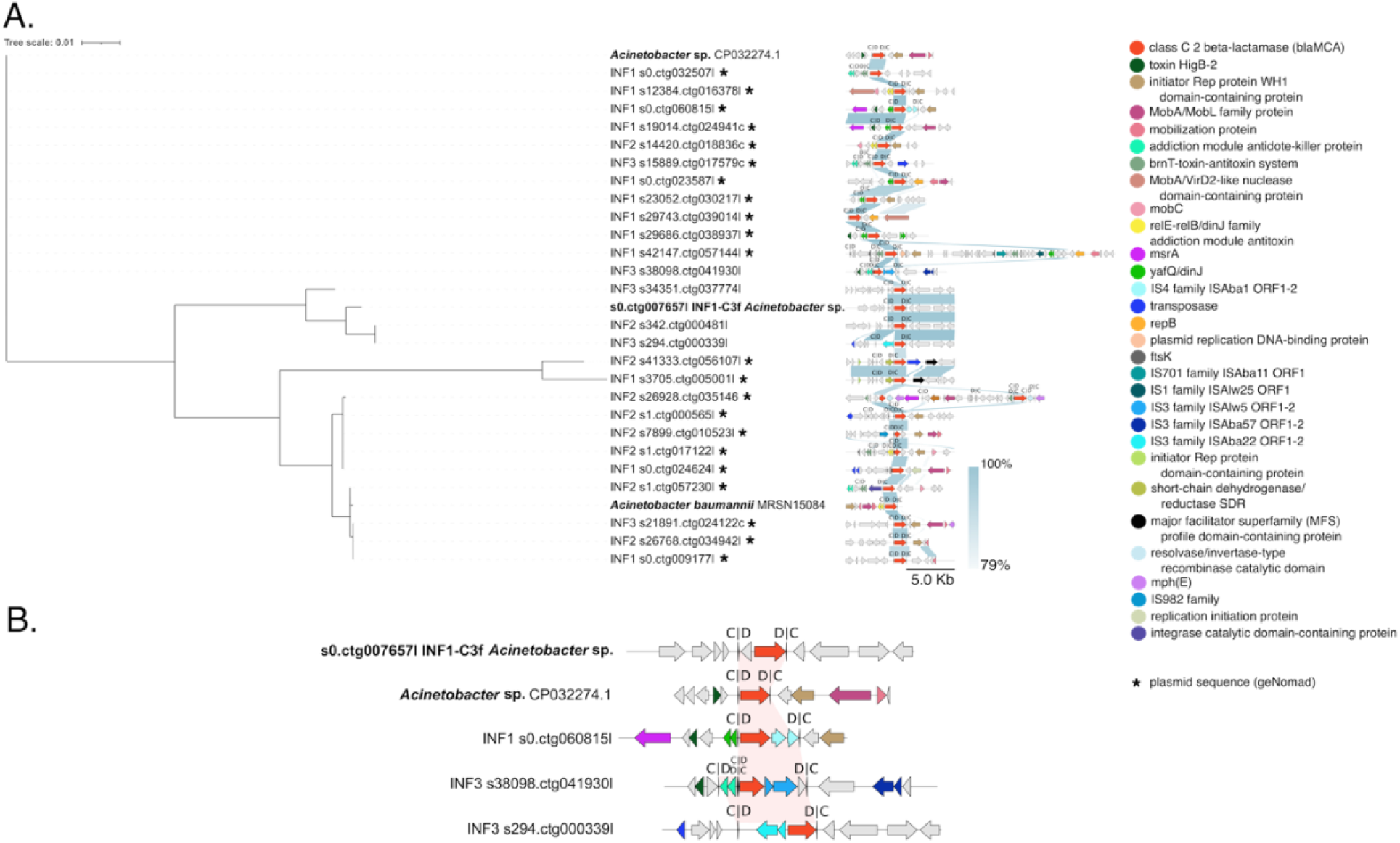
**A.** The phylogeny tree of class C beta-lactamase genes (also known as *bla*MCA) (shown in red) in the methylation-based bins of connected contigs and unbinned contigs with matching genes. *Acinetobacter baumannii* (MRSN15084) and *Acinetobacter* sp. (CP032274.1) were included as references. The annotations of the target gene flanking sequences are shown for 5 kb upstream and downstream. **B.** More detailed visualization shows the gene arrangement of the p*dif*-modules containing IS3 or IS4 family transposases alongside the *bla*MCA gene. Bin INF1-C3f and *Acinetobacter* sp. (CP032274.1) sequences are included in the figure for comparison.

One major branch of the *bla*MCA gene tree was formed by the reference sequence *Acinetobacter* sp. (CP032274.1) and unbinned 12 predicted plasmid sequences from influent wastewater possessing identical ARG sequences but highly diverse genetic contexts (**Fig. 3A**). Of the two other main tree branches, the *bla*MCA gene carried by *Acinetobacter* sp. genome bin INF1-C3f showed 92 % (1122/1215 bp) sequence similarity with both references (MRSN15084 and CP032274.1), while the beta-lactamases carried by the references (MRSN15084 and CP032274.1), that were most distantly related within the tree, showed 89 % (1081/1216 bp) sequence similarity with each other (**Table 1**, **Fig. 3A**).

The non-plasmid context, as well as the conserved gene arrangement across the sequences of the branch that was represented by genome bin INF1-C3f *Acinetobacter* sp. (**Table 1**), suggested a chromosomal context for *bla*MCA in these sequences (**Fig. 3A**, **Table S6**). This was contrary to what was observed for the remaining sequences, where the flanking regions are variable despite the identical ARG sequences (**Fig. 3A**), indicating recent decontextualization events. Nevertheless, common to all sequences were the presence of p*dif* sites (**Fig. 3**, **Table S6**, **Table S7**). More specifically, in most cases, these C|D and D|C-oriented site-specific recombination sites were detected immediately bordering the *bla*MCA gene, forming a *bla*MCA-p*dif* module, which was, in a few cases, present multiple times along the investigated sequence region (**Fig. 3A**).

In three instances, different IS3 or IS4 family transposase genes co-occurred with *bla*MCA in the same p*dif* modules suggesting their possible role in the transmission or expression of *bla*MCA gene (**Fig. 3B**). For the INF1-C3f *Acinetobacter* sp. genome bin and its cluster companions, genes encoding proteins of unknown function were located upstream *bla*MCA of (**Fig. 3**). Overall, the studied regions contained a high number of p*dif* sites proximal to not only *bla*MCA, but also other genes (**Fig. 3**). For instance, p*dif* modules encoding *brnT* and toxin-antitoxin system genes, referring to plasmid maintenance functions, with or without integrase catalytic domain containing protein encoding gene were repeatedly observed in multiple sequences (**Fig. 3A**). Altogether, the high frequency of p*dif* modules likely contributes to the extensive gene content variation seen across the studied contexts suggesting active shuffling of *bla*MCA not only among plasmid but also chromosomal contexts at least among *Acinetobacter* species (**Fig. 3**).

### Identification of *Simplicispira* sp. as a newly described host for the integron–encoded ESBL gene *bla*OXA-129

The established extended-spectrum beta-lactamase (ESBL) phenotype encoding gene *bla*OXA-129 was identified in two methylation-based genome bins assigned as *Simplicispira sp017988165* (prev. *Giesbergeria sp017988165*) from two separate influent samples (bins INF1-C3, INF3-C5) (**Fig. 4**, **Table 1**, **Table S8**). The bins of connected contigs were composed of 20 and 18 contigs and showed 31.41 and 38.56 completion and 3.4 and 1.35 redundancy, respectively (**Table 1**). Further exploration of matching genes in the full data revealed two more occurrences of this gene among the unbinned contigs from influent samples (**Fig. 4**, **Table S8**). The ARGs in the genome bins were 99.90-100 % and in the unbinned contigs 99.75-100 % identical (802-804/804 bp) to the *bla*OXA-129 genes in the reference sequences of clinical class 1 integron encoded genes carried by *E. hormaechei* (FJWZ01000025, the reference gene in the ResFinder database) and of *Pseudomonas aeruginosa* (CP031449.2).

**Fig. 4.**
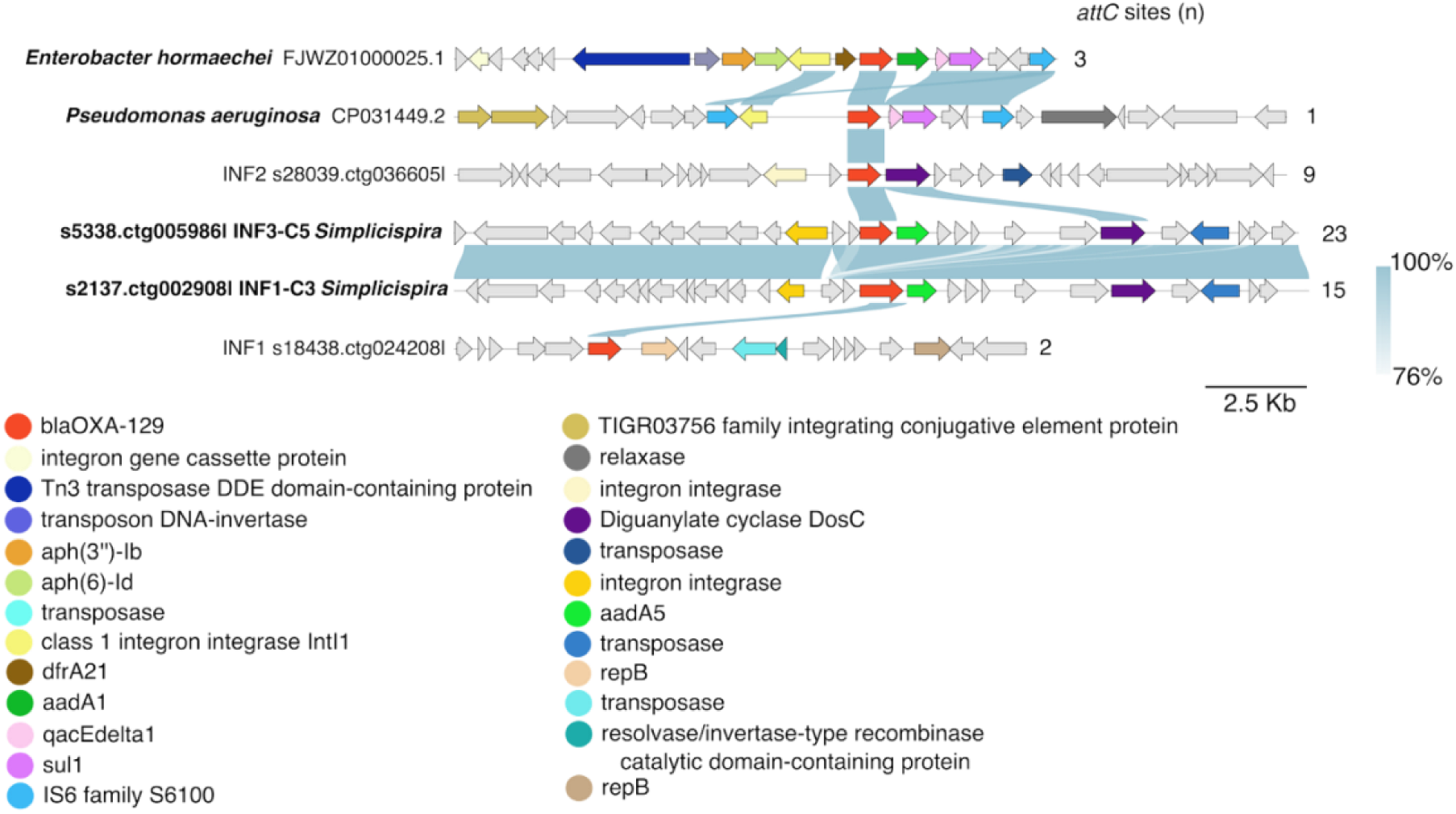
A comparative analysis of genetic features of the *bla*OXA-129 gene (shown in red) hosts identified in sequences of this study and previously described isolates of *Enterobacter hormaechei* (FJWZ01000025.1) and *Pseudomonas aeruginosa* (CP031449.2). The annotations of the target gene flanking sequences are shown for 10 kb upstream and downstream.

The flanking region of the *bla*OXA-129 displayed high similarity between the two *Simplicispira* sp. genome bins but differed more from the reference sequences (**Fig. 4**). However, a common feature among all but one of the sequences was the presence of traits indicating integron gene cassette context of the *bla*OXA-129 gene (**Fig. 4**). These included the presence of a gene annotated as integron integrase upstream of *bla*OXA-129, detection of aminoglycoside resistance gene *aadA* (*aadA1* in *P. aeruginosa* and *aadA5* in *Simplicispira* sp.) downstream of *bla*OXA-129 and finally numerous *attC* sites flanking the genes which were numerous especially in the *Simplicispira* sp. sequences (**Fig. 4**). The integrases of *Simplicispira* sp. were dissimilar to those in *E. hormaechei* and *P. aeruginosa* which in turn resembled each other (**Fig. 4**). However, according to BLAST search against public sequence databases, similar integron integrase genes to those in *Simplicispira* sp. are found in multiple other bacterial species such as *Klebsiella* sp. and *Acidovorax* sp. (data not shown). Another notable difference between the context regions of the clinical *E. hormaechei* and of *Pseudomonas aeruginosa* integron cassettes and those in *Simplicispira* sp. was the presence or absence of IS6 family IS6100 transposases, respectively (**Fig. 4**). Particularly, the arrangement of the two identical IS6100 genes flanking entire the integron gene cassette in composite transposon orientation in *Pseudomonas aeruginosa* may have a key role in mobilizing the cassette (**Fig. 4**). While one copy of IS6100 is observed also in *E. hormaechei*, none are found in the cassettes of this study (**Fig. 4**).

### Class 1 integron cassette encoded *sul1* was hosted by scarcely studied bacteria of the order UBA1845 (class Phycisphaerae)

Sulfonamide resistance gene variant *sul1-9* was detected within clinical class 1 integron gene cassette in two high-quality genome bins (bins SLU2-C4, SLU3-C1) from two separate sludge samples (**Table 1**, **Table S9**, **Fig. 5**). These bins constituting of 12 and 4 contigs showed high completion and redundancy values of 90.86 and 97 and 3.74 and 4.14 respectively and were assigned as *JAUCQB01* sp. of UBA1845 order within Phycisphaerae class and Planctomycetota phylum (**Table 1**, **Table S9**). Gene *sul1-9* differs from its most similar variant *sul1-2* by an 11 bp long tail and one nucleotide substitution.

**Fig. 5.**
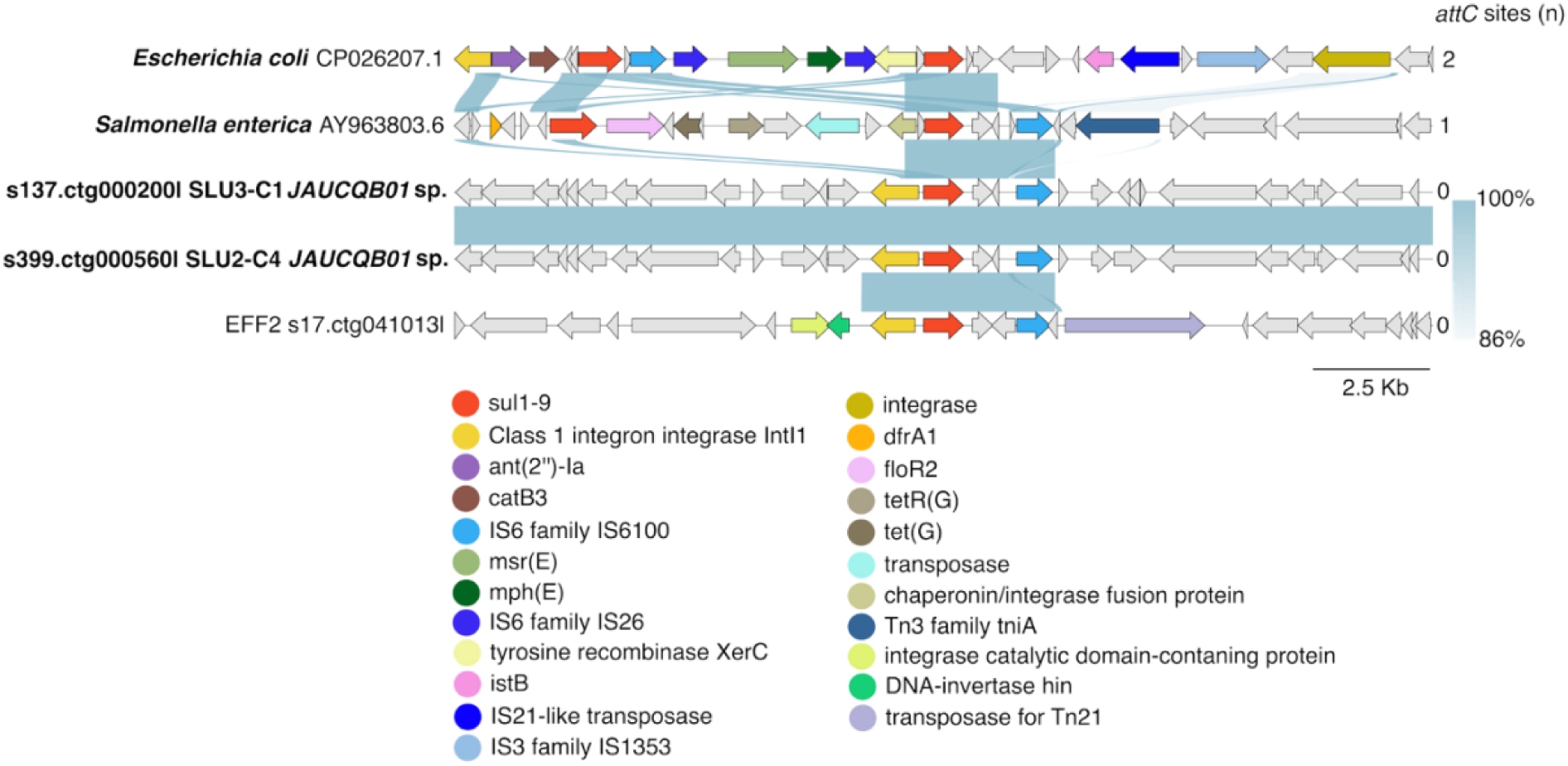
The phylogeny tree of *sul1* genes (shown in red) in the methylation-based bins of connected contigs and unbinned contigs. Reference sequences of *Escherichia coli* (CP026207.1), *Klebsiella* sp. (CP055486.1), and *Pseudomonas aeruginosa* (CP021775.1) were included in the comparative genetic analysis of the *sul1-9* flanking region of 10 kb up- and downstream length.

Exploring the *sul1-9* contexts of the sequences of this study and of the previously reported hosts of *Escherichia coli* (CP026207.1) and *Salmonella enterica* (AY963803.6, the reference gene in the ResFinder database) revealed that common to all these sequences was the presence of the *intI1* gene, located adjacent to the *sul1-9* (**Fig. 5**, **Table S9**). Moreover, similarly to the *bla*OXA-129 encoding integron gene cassettes, IS6 family IS6100 transposase was detected in all investigated sequences including both reference sequences from public databases as well as the newly described *sul1-9* hosts of *JAUCQB01* sp. and unbinned contig characterized in this study (**Fig. 5**). In contrast to the *E. coli* and *S. enterica* reference strains, no additional ARGs were present in the integron cassettes of this study (**Fig. 5**). The partially shared sequence between the *JAUCQB01* sp. genome bin sequences and the unbinned contig described in this study and extending the core structure of the integron supports the wide occurrence and hence mobility of this *sul1-9* cassette with IS6100 in different contexts (**Fig. 5**). However, unlike for the reference strains, no *attC* sites were reported for the sequences of this study (**Fig. 5**).

### Gene *erm*(F) is carried by multiple different environmental Bacteroidales species in the sludge

The acquired macrolide–lincosamide–streptogramin B (MLSB) resistance gene *erm*(F) was the most common established resistance gene present among the ARG-encoding genome bins and was also detected in numerous unbinned contigs (**Table 1**, **Fig. 6**, **Table S10**). The 13 visualized *erm*(F) encoding bins of connected contigs were primarily derived from sludge samples, but one influent derived *erm*(F) was also reported (**Table 1**, **Fig. 6**). The completeness and redundancy values of the *erm*(F) genome bins ranged from 17.49 to 94.92 and 0.02 to 3.84 (average completion 58.51 and redundancy 2.11), and the number of included contigs per genome bin was on average 11 (**Table 1**). Altogether, these genome bins represented 7 different bacterial families within the Bacteroidales order (**Table 1**). Of these, only the one genome bin represented the human gut-associated *Bacteroidetes* sp. (*Bacteroidaceae* family) *erm*(F) host, and this bin was recovered from the influent. Instead, the remaining bins of connected contigs from sludge represented the so-called environmental lineages of Bacteroidales, including families *Dysgonomonadaceae, Paludibacteraceae, Tenuifilaceae, Prolixibacteraceae, VadinHA17,* and *ML635J-15* (**Fig. 6**, **Table S10**). These findings align with the observed Bacteroidota phylum dynamics throughout the wastewater treatment process (**Fig. S6**, **Fig. S9**): While the *Bacteroidaceae* family was clearly dominant in the influent, the taxa profile was substantially different in sludge (**Fig. S5**, **Fig. S6**), where multiple other Bacteroidales families were present in high relative abundances, particularly the *erm*(F) hosting families in the sludge (**Fig. 6**, **Fig. S6**, **Fig. S9**).

**Fig. 6.**
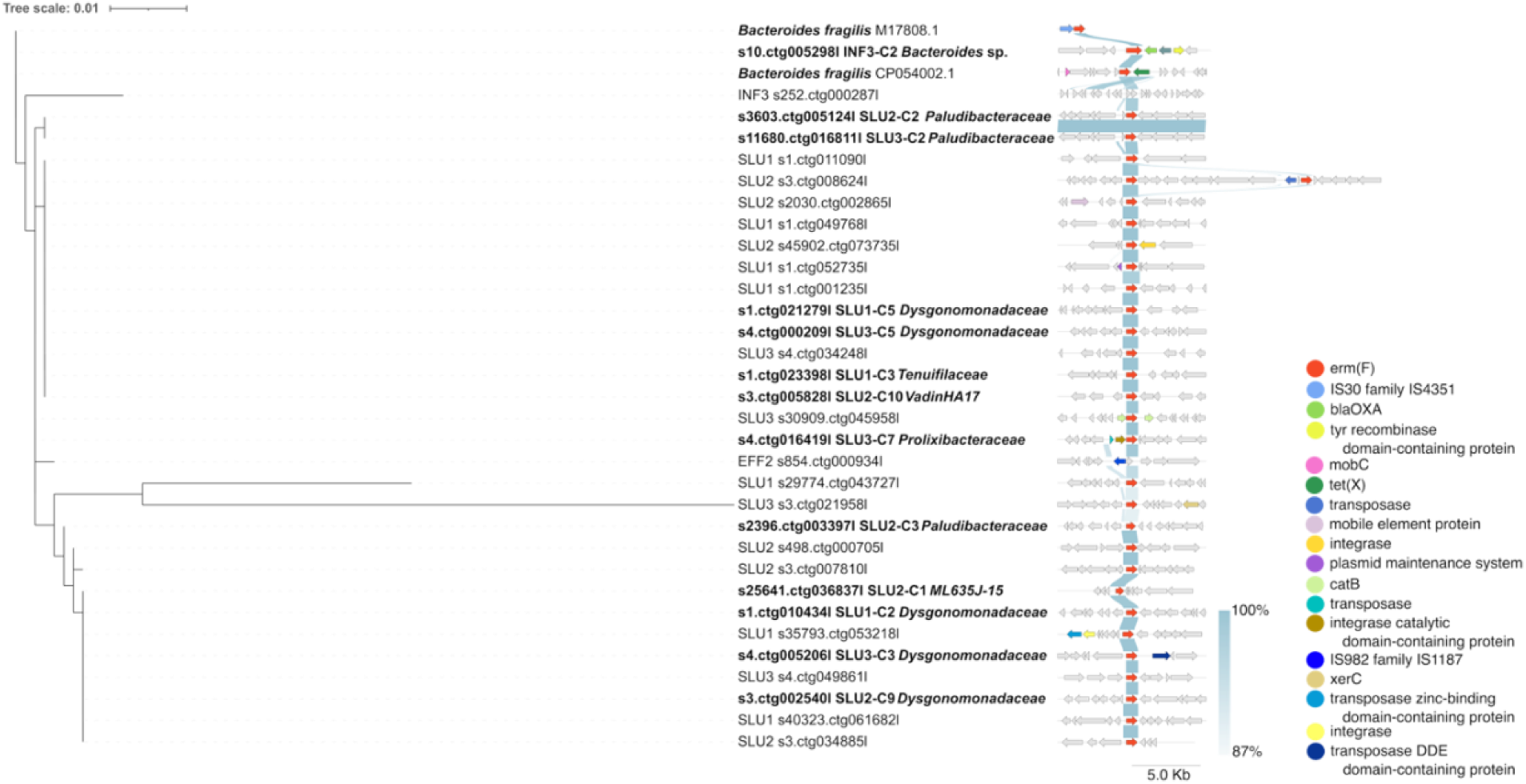
Gene phylogeny tree of the *erm*(F) gene (shown in red) found in genome bins of different Bacteroidales order bacteria and unbinned sequences from wastewater. The *erm*(F) genes encoded by *Bacteroides fragilis* (M17808.1) and *B. fragilis* (CP054002.1) and their flanking regions were included as references. Flanking regions of 5 kb upstream and downstream of the *erm*(F) were included in the genetic context visualization. Family-level taxonomical identity of the genome bins is shown in bolded text.

Phylogenetic analysis revealed that the identified *erm*(F) genes were highly similar despite the variable genetic context indicative of horizontal gene transfer (HGT) (**Fig. 6**). Most of the genes shared 99-100 % sequence identity with the reference *erm*(F) gene (*Bacteroides fragilis* (CP054002.1 and M17808.1, reference gene in ResFinder database) while for the two most dissimilar genes the similarity to reference gene was lower (95% and 92%) (**Fig. 6**). High plasticity among the *erm*(F) flanking regions was emphasized by the observation of clearly distinct context sequences, even when originating from genome bins of the same species (*Seramator thermalis* bins SLU1-C5, SLU1-C2, SLU3-C3) (**Fig. 6**, **Table S10**). Moreover, in some cases, MGEs such as transposases of IS30 and IS987 families are detected near *erm*(F), supporting the rearrangement of genes. However, no CTnDOT transposases, typically explaining the transfer of *erm*(F) among human gut-associated *Bacteroidales* (70), were detected. In the *Bacteroides* sp. genome bin (bin INF3-C2), the *erm*(F) seems to be part of an integron gene cassette along with *aadS* and *bla*OXA ARGs (**Fig. 6**). Altogether, both the high diversity of *erm*(F) hosts and contexts suggested that *erm*(F) is horizontally moving between various, not only human-gut-associated *Bacteroidales* hosts.

## Discussion

Although the novel resistance determinants arising from environmental microbiomes, especially with the implications of gene mobility, have been assigned as critical contributors to the current rapidly developing resistance problem, methodological limitations have impaired our ability to recognize them (7, 12). While a wide variety of ARGs are being detected in diverse environments and microbiomes, the crucial information regarding their genetic and bacterial contexts is often missing (12, 18). Instead, in many metagenomic studies, the highest-abundance taxa are often reported separately, although those bacteria rarely represent the ARG carriers in those communities (9, 25). In this study, by applying metagenomic long-read sequencing and bacteria-specific methylation signals, we were able to identify the host bacteria of the emerging resistance threats whose characterization has been delayed by technical limitations and prioritized clinical settings.

The latent class D beta-lactamase gene that was carried by *Arcobacter cryaerophilus* showed potential as a newly emerging, uncharacterized ARG that could be transferred among *Arcobacter* or related species primarily due to the composite structure of the flanking transposase genes, similar to what was reported before with other ARGs (71–73). The IS21 family transposase situated in proximity may also have a role in the mobilization (74). Although the studied beta-lactamase did not match any of the existing proteins in the databases, the best matches included proteins encoded by *Sulfurospirillum,* which, similarly to *Arcobacter,* represents the Campylobacterales order. Moreover, the observation of similar transposase genes but not their adjacent genes, both in *Sulfurospirillum* and *Arcobacter* sequences of the public sequence databases, could suggest that transmission of genes facilitated by these composite transposases occurs at least between closely related species. These observations may also point to *Sulfurospirillum* as the ancestral host of the beta-lactamase described here before its transfer to the species *Arcobacter cryaerophilus*.

According to our data, the remaining class D beta-lactamase genes represented likely chromosomal, non-mobile variants of this gene, indicated by the conserved context across different *Arcobacter* species, including *A. butzleri*, *A. defluvii, A. articola*, *A. sp017995775, A. suis,* and unidentified species of *Arcobacter*. Of these genes, only the one hosted by *A. butzleri* showed similarity to any previously established ARGs, namely the *bla*OXA-464 gene, reported previously in wastewater (75) (Dekic Rozman *et al.* manuscript titled “Emerging Food- and Waterborne Pathogen *Arcobacter* in Wastewater: Diversity and Antibiotic Resistance” under review).

Although *Arcobacteraceae* are among the most dominant families in influent wastewater (76, 77), a substantial diversity within this bacterial group has remained uncharacterized (Dekic Rozman *et al.* manuscript under review). At the same time, indications of the contribution of *Arcobacter* species to the ARG load in wastewater have been reported, yet the extent and underlying mechanisms remain poorly understood (76, 77). The lack of standardized cultivation and phenotyping protocols (Dekic Rozman *et al.* manuscript under review) and the genome plasticity, challenging the genome-resolved metagenomics for *Arcobacter* species (78), have delayed the description of genotypic and phenotypic resistance features of *Arcobacter* species. Therefore, the description of the emerging mobile class D beta-lactamase here elucidates the less-understood genetic mechanisms underlying resistance of *Arcobacter* species. In addition to the human clinical cases and wastewater, where they act as anoxic denitrifiers (79), *Arcobacter* have been isolated from food and animals, for which they are usually asymptomatic, driving their unnoticed dissemination (80).

The widespread distribution of *Arcobacteria* spp. across both external environments and human activity-linked sources highlights the significance of the *Arcobacter* species serving as the mediators of the established and emerging ARGs between non-pathogenic and pathogenic species. Given that *A. cryaerophilus* is one of the *Arcobacter* species most commonly isolated in clinical cases (80), the mobile beta-lactamase gene described in this study within *A. cryaerophilus* host could cautiously be interpreted as an example of the ARG decontextualization from its original genomic context and further into a pathogenetic background by MGEs. Given the limited evidence presented in this study, we cautiously propose that the potential past host in this case may have represented a species of genus *Sulfurospirillum*, which are taxonomically and functionally related to *Arcobacter* (81).

Our description of the carriage and mobility of the class C beta-lactamase gene *bla*MCA provides another instance of emerging resistance risk that is not yet widely recognized. To our best knowledge, this gene has been referred to only once before (49). Its absence in most of the widely used ARG databases, such as ResFinder (44) and CARD (82), and therefore its scarce representation in the literature would suggest that *bla*MCA is considered rather an intrinsic gene than an acquired ARG. However, based on the active reshuffling of *bla*MCA-encoding p*dif*-modules across diverse genomic contexts demonstrated by our results, we argue that *bla*MCA may show potential as an emerging clinically relevant ARG. While the XerC/XerD (*dif*) binding site-specific recombination is central in chromosomal dimer resolution among all bacterial lineages, p*dif*-module trafficking, which occurs primarily among *Acinetobacter* plasmids (83), allows the rapid acquisition of adaptation-improving genes by site-specific homologous recombination (84). Despite its ancient origin, this mechanism is thought to have gained additional functionality as an ARG mobilizing mechanism along the post-antibiotic era (83). The full understanding of the significance of the p*dif* sites as mobile genetic elements has been impeded by the difficulty in identifying them (49) as well as the detection limitations of the current tools focusing on well-established ARG-modules and pathogens (49, 85), which has restricted the discovery of emerging latent ARGs and their non-pathogenic hosts possibly harboring novel p*dif* modules.

Building on previous knowledge on *bla*MCA that is based on observations on publicly available plasmid sequences, which are likely biased towards opportunistic pathogens (49), the findings presented here add a real–world example of the p*dif*-module rearrangement at a microbial community level, beyond the boundaries of genetic context or taxonomical lineages. This was exemplified here by the detection of *bla*MCA-p*dif* modules within both likely chromosomal *Acinetobacter* sp. host and plasmid-sequence contexts. Chromosome-integrated p*dif*-modules encoding ARGs have been reported before (86), and on the other hand, the so-called additional chromosomal *dif* sites, observed in *Acinetobacter* spp. before (84), may act as reservoirs for adaptation-improving genes transferred to the plasmid p*dif*-modules.

In addition to the p*dif* modules consisting of *bla*MCA formed by the inverted p*dif*-sites (C|D – D|C orientation) flanking *bla*MCA in *Acinetobacter,* reported also earlier (49), we also found modules in which *bla*MCA was coupled with IS elements. The presence of complete sets of ORFs of either IS3 or IS4 family transposases located up- or downstream of *bla*MCA within the same p*dif* module could be an indication for further mobility and transcription of the *bla*MCA gene by an IS-encoded promoter (87). Similar observations of IS elements within other kinds of ARG-p*dif*-modules have been made before (88), but not in association with the *bla*MCA.

While particularly common in *Acinetobacter* plasmids (83), identical p*dif*-modules have been detected across genera (49), confirming that p*dif*-module-mediated spread of ARGs across bacterial lineages does occur, and environments like wastewater could host conditions enabling such cross-species transmission. Unfortunately, we failed to assign taxonomic identity to any other than *Acinetobacter* sp. sequences hosting *bla*MCA. Even though the resistance phenotype of this beta-lactamase gene remains to be confirmed, the frequent rearrangement of *bla*MCA p*dif*-modules suggests that it confers an adaptive advantage to its recipient under selective conditions, such as those caused by the presence of antibiotic residues in the influent wastewater. Overall, our description of the *bla*MCA reshuffling at least among *Acinetobacter* chromosomes and plasmids, supported by previous findings (49), adds to the extensive list of antibiotic resistance features typical for this genus, which comprises members across ecological niches from those flagged as the top priority pathogens regarding multidrug resistance by WHO (89) to the non-pathogenic environmental species.

The risk potential of environmental microbiome origin ARGs is defined by the likelihood and consequences of their transmission to human pathogens (7). As both hosts of the latent ARGs described above, *Arcobacter* and *Acinetobacter*, represent genera that include both opportunistic pathogens and environmental lineages, the frequency of HGT and likelihood of ARG transmission across pathogenic and non-pathogenic strains is less rare than is considered for ARGs emerging from strictly environmental bacteria (12, 15). This is particularly concerning, given that only one successful transmission to a clinically important strain is required for the emergence of a novel clinically important resistance threat (12).

In addition to the novel, predicted ARGs, our methylation-based binning approach allowed the identification of previously uncharacterized environmental hosts for functionally confirmed ARGs, shedding light on the hidden contributors to total resistance burden. To the best of our knowledge, *Simplicispira sp017988165* (prev. *Giesbergeria sp017988165*) described here as the carrier of the integron-encoded *bla*OXA-129 is the first non-pathogenic host identified for this gene. *Simplicispira* sp. are important denitrifiers under both aerobic and anoxic conditions in wastewater (79, 90, 91). *bla*OXA-129 was first described in integron gene cassettes of clinical isolates of *P. aeruginosa* collected in 2006-07 in China, showing carbapenem-sensitive ESBL phenotype (92) and in other opportunistic pathogens (*Enterobacter* sp.*, P. aeruginosa, K. pneumoniae, S. enterica* (93) and *C. resistens* (CP125947.1)) ever since.

Although the integrase encoding genes in *Simplicispira* and the previously described pathogens differ, the integron gene cassette context with numerous detected *attC* sites supports its possibility of acting as a source of ARGs to be translocated into other contexts. However, integrons are not mobile themselves (94). While two integron gene cassette flanking IS6100 transposases (72) were present in most but not all *bla*OXA-129 sequences reported before in opportunistic pathogens (93, 95), no such genetic elements required for the mobilization of the integron gene cassette were observed in *Simplicispira*. Altogether, our findings broaden the current knowledge about the host range and mobility of this gene and point to the potential of *Simplicispira* sp. acting as an environmental intermediate host of clinically relevant ARGs in wastewater.

Interestingly, according to our results, it appeared that the same IS6100 transposase (72), which has also previously been strongly associated with ARG mobilization (96), would have also been involved in the putative mobility of the class 1 integron-encoded *sul1-9* gene within the strictly environmental class Phycisphaerae bacteria (species *JAUCQB01* sp.) described here. However, detection of only one copy of the IS6100 gene within the *sul1-9* flanking region of both environmental Phycisphaerae and opportunistic pathogenic reference hosts *E. coli* and *S. enterica* may not yet warrant the transfer of its nearby genes (72).

Considering the available evidence, no ARGs have been described in this *JAUCQB01* sp. or within its higher taxonomic rank, order UBA1845 bacteria before. Although Phycisphaerae are being detected in various environments, the understanding of their functional characteristics and ecological roles is held back by the lack of cultivable reference strains, as is the case with order UBA1845 (97). Only recently, Lenferink and colleagues have discovered that the UBA1845 bacteria, mostly residing in engineered environments under anoxic conditions, such as wastewater, thrive in the presence of diverse sugar and complex carbon substrates such as extracellular polymeric substances extracted by primary producers (97). The presence of *sul1-9* as the only ARG in these cassettes of UBA1845 might point to an early evolutionary stage of these integron cassettes, as more ARGs are expected to accumulate in time under certain conditions (94). Our observations could be further explained by their chromosomal context, unlike the reference opportunistic pathogens’ plasmid- or genomic island-encoded (98) integron cassettes containing multiple consecutive ARGs.

The two example cases confirm that even the clinical integron-encoded ARG cassettes are well disseminated to taxa lacking any human health association, challenging the previous conception of integrons encoding ARG cassettes being disseminated mainly among opportunistic pathogens (94). Furthermore, these findings reveal insights into the so-called intermediate hosts and mobilizers of ARGs fostered by the wastewater treatment process before their putative irreversible transfer to human health-associated species (7, 8).

Similarly, our analysis revealed the host dynamics of macrolide-lincosamide-streptogramin resistance phenotype encoding (99) *erm*(F) gene throughout the wastewater treatment. While the identified *erm*(F) carrier in influent represented gut–associated Bacteroidales, no human-associated taxa but numerous other members of this order were distinguishable in the outlet sludge. The particularly well-distinctive value of methylation profiles in the detection of Bacteroidales has also been noted before, although in considerably less complex gut microbiomes (23). The *erm*(F) hosts in the sludge comprised 7 different families and 10 genera representing environmental groups of Bacteroidales. These species, such as those belonging to families *Dysgonomonadaceae*, *Paludibacteraceae,* and *Tenuifilaceae,* are less studied but common members of the bioreactor systems, specifically wastewater sludge (100–102). They are characterized by their metabolically versatile functions, for instance, denitrification and the capability for fermenting a wide range of mono- and disaccharides (79, 100–102).

The expanding host range and the highly variable genetic contexts in the sludge point to the horizontal mobility of the *erm*(F) among different bacteria, as noted before (103). However, as the host range revealed here and in previous work (15, 103) suggests, certain HGT barriers seem to restrict the transmission to Bacteroidota. Although various MGEs are detected among the gut-unrelated Bacteroidales hosts, no single specific mobility mechanism responsible for the horizontal mobility of *erm*(F) could be identified here or by previous work (103). While the CTnDOT (Conjugative Transposon from strain *Bacteroidetes thetaiotaomicron* DOT) conjugative transposons are the central carriers of *erm*(F) among gut-associated Bacteroidales species (70), some other mobility mechanism could explain the presence of identical or highly similar *erm*(F) gene sequences within highly variable genetic contexts among the different environmental Bacteroidales hosts, even within species, as described here for *Seramator thermalis*. Similarly, invertible promoter-regulated ARGs located in integrative conjugative elements (ICEs) are highly enriched within Bacteroidales, particularly among the gut–associated species (70, 99, 104). Although previous work has reported a lower prevalence of these mobile invertible promoter-associated ARGs among environmental Bacteroidota lineages (105), wastewater was not included among the environments explored in this study by Jiang and colleagues (105), leaving their occurrence among wastewater Bacteroidales families unexplored.

It may also be that the acquisition and later chromosomal integration of *erm*(F) by diverse Bacteroidales species has occurred in the distant evolutionary past, and the selective conditions present in sludge promote the growth of *erm*(F) carrying strains. Altogether our results regarding the observed increase in the abundance of *erm*(F) hosting taxa in the sludge supported the view that the high abundance of this gene, particularly through its enrichment in the sludge, noted also by previous studies (75, 103, 106, 107), is not due to the proliferation of one species by the treatment process but rather multiple taxa that are enriched under the conditions present in the sludge. Furthermore, the geographical differences reported for the *erm*(F) prevalence in sludge (103) might reflect the differences in wastewater treatment systems, mirrored as the Bacteroidales composition differences and their interactions facilitating the *erm*(F) carriage and spread. Bacteria of the *Bacteroides fragilis* group have been recognized as resistance gene reservoirs before (99). However, our results suggest that this characteristic is not exclusive to the gut-associated Bacteroidales but is likewise observed in environmental lineages of Bacteroidales, which might serve as environmental reservoirs of *erm*(F) for further transmission to the clinically relevant species.

Taken together, by linking ARGs to their broader genetic contexts and hosts at the microbial community level using bacteria-specific methylation signals, this study addresses one of the key challenges in the field hindering the environmental resistome risk assessment (7, 12). Another recent approach to this task is high–throughput single–cell metagenomic sequencing, which uses individually barcoded cells for sequencing (108). While promising, this approach has certain limitations, including laborious sample preparation, dependency on specific reagents and equipment, and possible bias against gram-negative bacteria (108). A strictly computational strategy for coupling taxonomy, ARGs, and their genetic context draws upon alignment-free methods with CARD k-mers (109). However, the main weakness of this method is its reliance on established ARGs in a certain database and the assumption that these ARGs occur in expected context sequences, and especially among pathogens, while latent ARGs and any ARGs within new contexts are missed. In contrast, by the characterization of newly described ARG hosts and contexts for both latent and established ARGs without the dependency of prior knowledge, we show that the approach presented in our study mitigates this limitation.

The methylation-based binning here enabled the description of several putative emerging ARGs and their hosts and confirmed the intermediate host roles of strictly environmental and understudied taxa. Even though the obtained genome bins represented predominantly partial genomes rather than fully complete MAGs, we were able to assign the taxonomic identity and set the ARG into its wider genetic context to evaluate its mobility potential. One limitation of our approach, however, was the loss of data, as not all contigs displayed distinct methylation profiles or no modifications at all, which led to fewer data available for downstream analyses, namely the genome bin curation. Technical factors such as the lower sensitivity for detecting certain base modifications (110, 111) or simply lower coverage, as in the fewer mapping reads, and their attached modification data of certain strains (23) may have affected this. Another technical bottleneck arose from the manual work required for refining the genome bins of clustered contigs.

Importantly, the data-loss limitation did not introduce false positives but rather gaps in the dataset, and the potential biases introduced by our approach would be different from those of more traditional binning algorithms. For instance, our findings regarding the multiple *erm*(F) *Seramator thermalis* hosts showing diverse methylation profiles and genetic contexts indicate the concurrent presence of multiple strains of this species and their distinct identification by methylation analysis in complex communities. Previous observations on strain- (111, 112) or serovar- (113) level differences in methylation profiles supported our findings. This variation is explained by the transcriptional regulation roles of epigenetic variation mediated by methylation (27, 111, 112) as well as more frequent HGT of restriction modification systems between closely related taxa than more distant lineages (114). Additionally, our approach circumvented the common bias towards the high-abundance taxa that is descriptive for traditional binning methods (9, 25). This is of high importance when aiming to understand the ARG dynamics often mediated by rare taxa (9, 25) within highly complex environmental microbiomes.

Finally, despite considerable research efforts (23, 115), the task of assigning the host-plasmid linkages in metagenomic data has remained without a definitive resolution, particularly in complex communities. Given that the use of methylation profiles ignores the variation in coverage and sequence composition, which currently constrains the correct binning of sequences rich in MGEs, holds great potential for this (22, 23). Building on the valuable work carried out on simpler communities (23), our study demonstrates that methylation-based binning can be used to resolve plasmid origins also in highly complex communities, although with critical limitations. Although we could assign the bacterial hosts for multiple plasmids, unfortunately, none of them represented ARG-encoding plasmids, which are more likely to be those categorized as conjugative or mobilizable plasmids (116). Despite their multi-host history, they are considered to acquire the methylation motif profile of their current host rapidly (22, 23), owing to the evolutionary arms race between plasmids and host restriction-modification systems (116, 117). However, the simultaneous carriage of similar plasmids by different host bacteria may confuse the methylation profile detection by our approach, as reads coming from different strains and hence with mixed methylation signatures are aligned to the same sequence. Given that these most frequently horizontally moving broad-host range plasmids are also the most important drivers of ARG carriage and spread (118), this highlights a critical limitation of our method, in the context of data with the level of complexity examined here. Alternatively, erroneous assembly (21) might explain the poor detectability of these plasmids and their methylation-resolved origins because their mosaic architecture, shaped by embedded MGEs, is prone to fragmentation into smaller contigs (118). This, in turn, would lead to a decreased likelihood of capturing unique methylation profiles for the plasmid sequences to match with those of their host genomes (23).

## Conclusions

Our work sheds light on the previously hidden intermediate hosts and mobility mechanisms of established and newly emerging resistance threats in highly diverse wastewater samples. Furthermore, these results highlight the importance of the culture-independent investigations of ARGs and their carriers. Further investigations are needed to examine the broader occurrence of the latent beta-lactamases and, most importantly, to verify the resistance phenotypes of these ARGs, as their description here relied solely on computational predictions.

## Data Availability

All data produced in the present work are contained in the manuscript

## Acknowledgements and study funding

This work was supported by the Research Council of Finland funding for the Multidisciplinary Center of Excellence in Antimicrobial Resistance Research (V.M. 364234, 346128, M.V. 364231, 346125). M.M. received funding from the MBDP doctoral program at the University of Helsinki. We would like to acknowledge CSC, IT Center for Science, Finland, for providing the computational resources for the study, and the DNA Sequencing and Genomics Laboratory (supported by HiLIFE and Biocenter Finland funding), Institute of Biotechnology, University of Helsinki, for sequencing. Open access was funded by the Helsinki University Library.

## Author contributions

Conceptualization, A.K., M.M., V.M. Methodology — H.P. developed the methylation–detection algorithm under the supervision of V.M., and D.K. — M.M. performed the application of the methylation–detection algorithm, curation of bins of connected contigs, comparative genomics investigation, and all associated bioinformatic analyses under the supervision of A.K. Data acquisition, A.K. Writing — original draft, M.M. Writing — review and editing, all authors. Supervision, A.K., M.V., and V.M. Funding acquisition, A.K., M.V., and V.M.

## References

1. Hutchings M, Truman A, Wilkinson B. 2019. Antibiotics: past, present and future. Curr Opin Microbiol 51:72–80.

2. Naghavi M, Vollset SE, Ikuta KS, Swetschinski LR, Gray AP, Wool EE, Robles Aguilar G, Mestrovic T, Smith G, Han C, Hsu RL, Chalek J, Araki DT, Chung E, Raggi C, Gershberg Hayoon A, Davis Weaver N, Lindstedt PA, Smith AE, Altay U, Bhattacharjee N V., Giannakis K, Fell F, McManigal B, Ekapirat N, Mendes JA, Runghien T, Srimokla O, Abdelkader A, Abd-Elsalam S, Aboagye RG, Abolhassani H, Abualruz H, Abubakar U, Abukhadijah HJ, Aburuz S, Abu-Zaid A, Achalapong S, Addo IY, Adekanmbi V, Adeyeoluwa TE, Adnani QES, Adzigbli LA, Afzal MS, Afzal S, Agodi A, Ahlstrom AJ, Ahmad A, Ahmad S, Ahmad T, Ahmadi A, Ahmed A, Ahmed H, Ahmed I, Ahmed M, Ahmed S, Ahmed SA, Akkaif MA, Al Awaidy S, Al Thaher Y, Alalalmeh SO, AlBataineh MT, Aldhaleei WA, Al-Gheethi AAS, Alhaji NB, Ali A, Ali L, Ali SS, Ali W, Allel K, Al-Marwani S, Alrawashdeh A, Altaf A, Al-Tammemi AB, Al-Tawfiq JA, Alzoubi KH, Al-Zyoud WA, Amos B, Amuasi JH, Ancuceanu R, Andrews JR, Anil A, Anuoluwa IA, Anvari S, Anyasodor AE, Apostol GLC, Arabloo J, Arafat M, Aravkin AY, Areda D, Aremu A, Artamonov AA, Ashley EA, Asika MO, Athari SS, Atout MMW, Awoke T, Azadnajafabad S, Azam JM, Aziz S, Azzam AY, Babaei M, Babin FX, Badar M, Baig AA, Bajcetic M, Baker S, Bardhan M, Barqawi HJ, Basharat Z, Basiru A, Bastard M, Basu S, Bayleyegn NS, Belete MA, Bello OO, Beloukas A, Berkley JA, Bhagavathula AS, Bhaskar S, Bhuyan SS, Bielicki JA, Briko NI, Brown CS, Browne AJ, Buonsenso D, Bustanji Y, Carvalheiro CG, Castañeda-Orjuela CA, Cenderadewi M, Chadwick J, Chakraborty S, Chandika RM, Chandy S, Chansamouth V, Chattu VK, Chaudhary AA, Ching PR, Chopra H, Chowdhury FR, Chu DT, Chutiyami M, Cruz-Martins N, da Silva AG, Dadras O, Dai X, Darcho SD, Das S, De La Hoz FP, Dekker DM, Dhama K, Diaz D, Dickson BFR, Djorie SG, Dodangeh M, Dohare S, Dokova KG, Doshi OP, Dowou RK, Dsouza HL, Dunachie SJ, Dziedzic AM, Eckmanns T, Ed-Dra A, Eftekharimehrabad A, Ekundayo TC, El Sayed I, Elhadi M, El-Huneidi W, Elias C, Ellis SJ, Elsheikh R, Elsohaby I, Eltaha C, Eshrati B, Eslami M, Eyre DW, Fadaka AO, Fagbamigbe AF, Fahim A, Fakhri-Demeshghieh A, Fasina FO, Fasina MM, Fatehizadeh A, Feasey NA, Feizkhah A, Fekadu G, Fischer F, Fitriana I, Forrest KM, Fortuna Rodrigues C, Fuller JE, Gadanya MA, Gajdács M, Gandhi AP, Garcia-Gallo EE, Garrett DO, Gautam RK, Gebregergis MW, Gebrehiwot M, Gebremeskel TG, Geffers C, Georgalis L, Ghazy RM, Golechha M, Golinelli D, Gordon M, Gulati S, Gupta R Das, Gupta S, Gupta VK, Habteyohannes AD, Haller S, Harapan H, Harrison ML, Hasaballah AI, Hasan I, Hasan RS, Hasani H, Haselbeck AH, Hasnain MS, Hassan II, Hassan S, Hassan Zadeh Tabatabaei MS, Hayat K, He J, Hegazi OE, Heidari M, Hezam K, Holla R, Holm M, Hopkins H, Hossain MM, Hosseinzadeh M, Hostiuc S, Hussein NR, Huy LD, Ibáñez-Prada ED, Ikiroma A, Ilic IM, Islam SMS, Ismail F, Ismail NE, Iwu CD, Iwu-Jaja CJ, Jafarzadeh A, Jaiteh F, Jalilzadeh Yengejeh R, Jamora RDG, Javidnia J, Jawaid T, Jenney AWJ, Jeon HJ, Jokar M, Jomehzadeh N, Joo T, Joseph N, Kamal Z, Kanmodi KK, Kantar RS, Kapisi JA, Karaye IM, Khader YS, Khajuria H, Khalid N, Khamesipour F, Khan A, Khan MJ, Khan MT, Khanal V, Khidri FF, Khubchandani J, Khusuwan S, Kim MS, Kisa A, Korshunov VA, Krapp F, Krumkamp R, Kuddus M, Kulimbet M, Kumar D, Kumaran EAP, Kuttikkattu A, Kyu HH, Landires I, Lawal BK, Le TTT, Lederer IM, Lee M, Lee SW, Lepape A, Lerango TL, Ligade VS, Lim C, Lim SS, Limenh LW, Liu C, Liu X, Liu X, Loftus MJ, Amin HIM, Maass KL, Maharaj SB, Mahmoud MA, Maikanti-Charalampous P, Makram OM, Malhotra K, Malik AA, Mandilara GD, Marks F, Martinez-Guerra BA, Martorell M, Masoumi-Asl H, Mathioudakis AG, May J, McHugh TA, Meiring J, Meles HN, Melese A, Melese EB, Minervini G, Mohamed NS, Mohammed S, Mohan S, Mokdad AH, Monasta L, Moodi Ghalibaf AA, Moore CE, Moradi Y, Mossialos E, Mougin V, Mukoro GD, Mulita F, Muller-Pebody B, Murillo-Zamora E, Musa S, Musicha P, Musila LA, Muthupandian S, Nagarajan AJ, Naghavi P, Nainu F, Nair TS, Najmuldeen HHR, Natto ZS, Nauman J, Nayak BP, Nchanji GT, Ndishimye P, Negoi I, Negoi RI, Nejadghaderi SA, Nguyen QAP, Noman EA, Nwakanma DC, O’Brien S, Ochoa TJ, Odetokun IA, Ogundijo OA, Ojo-Akosile TR, Okeke SR, Okonji OC, Olagunju AT, Olivas-Martinez A, Olorukooba AA, Olwoch P, Onyedibe KI, Ortiz-Brizuela E, Osuolale O, Ounchanum P, Oyeyemi OT, Padukudru PAM, Paredes JL, Parikh RR, Patel J, Patil S, Pawar S, Peleg AY, Peprah P, Perdigão J, Perrone C, Petcu IR, Phommasone K, Piracha ZZ, Poddighe D, Pollard AJ, Poluru R, Ponce-De-Leon A, Puvvula J, Qamar FN, Qasim NH, Rafai CD, Raghav P, Rahbarnia L, Rahim F, Rahimi-Movaghar V, Rahman M, Rahman MA, Ramadan H, Ramasamy SK, Ramesh PS, Ramteke PW, Rana RK, Rani U, Rashidi MM, Rathish D, Rattanavong S, Rawaf S, Redwan EMM, Reyes LF, Roberts T, Robotham J V., Rosenthal VD, Ross AG, Roy N, Rudd KE, Sabet CJ, Saddik BA, Saeb MR, Saeed U, Saeedi Moghaddam S, Saengchan W, Safaei M, Saghazadeh A, Saheb Sharif-Askari N, Sahebkar A, Sahoo SS, Sahu M, Saki M, Salam N, Saleem Z, Saleh MA, Samodra YL, Samy AM, Saravanan A, Satpathy M, Schumacher AE, Sedighi M, Seekaew S, Shafie M, Shah PA, Shahid S, Shahwan MJ, Shakoor S, Shalev N, Shamim MA, Shamshirgaran MA, Shamsi A, Sharifan A, Shastry RP, Shetty M, Shittu A, Shrestha S, Siddig EE, Sideroglou T, Sifuentes-Osornio J, Silva LMLR, Simões EAF, Simpson AJH, Singh A, Singh S, Sinto R, Soliman SSM, Soraneh S, Stoesser N, Stoeva TZ, Swain CK, Szarpak L, Sree Sudha TY, Tabatabai S, Tabche C, Taha ZMA, Tan KK, Tasak N, Tat NY, Thaiprakong A, Thangaraju P, Tigoi CC, Tiwari K, Tovani-Palone MR, Tran TH, Tumurkhuu M, Turner P, Udoakang AJ, Udoh A, Ullah N, Ullah S, Vaithinathan AG, Valenti M, Vos T, Vu HTL, Waheed Y, Walker AS, Walson JL, Wangrangsimakul T, Weerakoon KG, Wertheim HFL, Williams PCM, Wolde AA, Wozniak TM, Wu F, Wu Z, Yadav MKK, Yaghoubi S, Yahaya ZS, Yarahmadi A, Yezli S, Yismaw YE, Yon DK, Yuan CW, Yusuf H, Zakham F, Zamagni G, Zhang H, Zhang ZJ, Zielińska M, Zumla A, Zyoud SHH, Zyoud SH, Hay SI, Stergachis A, Sartorius B, Cooper BS, Dolecek C, Murray CJL. 2024. Global burden of bacterial antimicrobial resistance 1990–2021: a systematic analysis with forecasts to 2050. The Lancet 404:1199–1226.

3. Ebmeyer S, Kristiansson E, Larsson DGJ. 2025. Unraveling the origins of mobile antibiotic resistance genes using random forest classification of large-scale genomic data. Environ Int 198:109374.

4. Jiang X, Ellabaan MMH, Charusanti P, Munck C, Blin K, Tong Y, Weber T, Sommer MOA, Lee SY. 2017. Dissemination of antibiotic resistance genes from antibiotic producers to pathogens. Nat Commun 8.

5. Gholipour S, Lee D, Tokuriki N. 2025. Molecular Evolution and Origins of Antibiotic Resistance Genes. Biochemistry 64:2516–2528.

6. Ebmeyer S, Kristiansson E, Larsson DGJ. 2021. A framework for identifying the recent origins of mobile antibiotic resistance genes. Commun Biol 4.

7. Klümper U, Fang P, Li B, Xia Y, Frigon D, Hamilton KA, Quon H, Berendonk TU, de la Cruz Barron M. 2025. Towards the integration of antibiotic resistance gene mobility into environmental surveillance and risk assessment. npj Antimicrobials and Resistance 3:81.

8. Larsson DGJ, Flach CF. 2022. Antibiotic resistance in the environment. Nat Rev Microbiol 20:257–269.

9. Inda-Díaz JS, Lund D, Parras-Moltó M, Johnning A, Bengtsson-Palme J, Kristiansson E. 2023. Latent antibiotic resistance genes are abundant, diverse, and mobile in human, animal, and environmental microbiomes. Microbiome 11.

10. Mlynarcik P, Zdarska V, Kolar M. 2025. Are Putative Beta-Lactamases Posing a Potential Future Threat? Antibiotics 14:1174.

11. Karkman A, Do TT, Walsh F, Virta MPJ. 2018. Antibiotic-Resistance Genes in Waste Water. Trends Microbiol 26:220–228.

12. Bengtsson-Palme J, Larsson DGJ. 2015. Antibiotic resistance genes in the environment: Prioritizing risks. Nat Rev Microbiol 13:396.

13. Sundqvist M. 2014. Reversibility of antibiotic resistance. Ups J Med Sci 119:142–148.

14. Markkanen M, Pezzutto D, Virta M, Karkman A. 2025. Sulfonamide resistance gene *sul4* is hosted by common wastewater sludge bacteria and found in various newly described contexts and hosts. Microbiol Spectr 14.

15. Parras-Moltó M, Lund D, Ebmeyer S, Larsson DGJ, Johnning A, Kristiansson E. 2025. The transfer of antibiotic resistance genes between evolutionarily distant bacteria. mSphere 10.

16. Cantón R, María González-Alba J, Galán JC, Stefani S. 2012. CTX-M enzymes: origin and diffusion. Front Microbiol 3:110.

17. Berglund F, Ebmeyer S, Kristiansson E, Larsson DGJ. 2023. Evidence for wastewaters as environments where mobile antibiotic resistance genes emerge. Commun Biol 6.

18. Martínez JL, Coque TM, Baquero F. 2014. What is a resistance gene? Ranking risk in resistomes. Nat Rev Microbiol 13:116–123.

19. Zhang A-N, Gaston JM, Dai CL, Zhao S, Poyet M, Groussin M, Yin X, Li L-G, Van Loosdrecht MCM, Topp E, Gillings MR, Hanage WP, Tiedje JM, Moniz K, Alm EJ, Zhang T. 2021. An omics-based framework for assessing the health risk of antimicrobial resistance genes. Nat Commun 4765.

20. Quince C, Walker AW, Simpson JT, Loman NJ, Segata N. 2017. Shotgun metagenomics, from sampling to analysis. Nat Biotechnol 35:833–844.

21. Abramova A, Karkman A, Bengtsson-Palme J. 2024. Metagenomic assemblies tend to break around antibiotic resistance genes. BMC Genomics 25:959.

22. Heidelbach S, Mølvang Dall S, Støtt Bøjer J, Nissen J, der Maas van, Sereika M, Kirkegaard RH, Jensen SI, Just S, Thorlacius-Ussing O, Hose K, Dyhre Nielsen T. Nanomotif: Identification and Exploitation of DNA Methylation Motifs in Metagenomes 1 using Oxford Nanopore Sequencing 2 3 10.1101/2024.04.29.591623.

23. Beaulaurier J, Zhu S, Deikus G, Mogno I, Zhang XS, Davis-Richardson A, Canepa R, Triplett EW, Faith JJ, Sebra R, Schadt EE, Fang G. 2018. Metagenomic binning and association of plasmids with bacterial host genomes using DNA methylation. Nat Biotechnol 36:61–69.

24. Frost LS, Leplae R, Summers AO, Toussaint A. 2005. Mobile genetic elements: The agents of open source evolution. Nat Rev Microbiol 3:722–732.

25. Diebold PJ, Rhee MW, Shi Q, Trung NV, Umrani F, Ahmed S, Kulkarni V, Deshpande P, Alexander M, Thi Hoa N, Christakis NA, Iqbal NT, Ali SA, Mathad JS, Brito IL. 2023. Clinically relevant antibiotic resistance genes are linked to a limited set of taxa within gut microbiome worldwide. Nat Commun 14.

26. Ghosh D, Veeraraghavan B, Elangovan R, Vivekanandan P. 2020. Antibiotic resistance and epigenetics: More to it than meets the eye. Antimicrob Agents Chemother 64.

27. Seong HJ, Han SW, Sul WJ. 2021. Prokaryotic DNA methylation and its functional roles. The journal of microbiology 59:242–248.

28. Tuomala H, Holtel J, Markkanen M, Patpatia S, Kaansalo K, Rolland C, Bayfield OW, Ranta K, Skurnik M, Wittmann J, Kiljunen S. 2025. The Host R-M Systems Change the Host Range of Staphylococcus Phage EBHT. Microbiologyopen 14.

29. Seong HJ, Roux S, Hwang CY, Sul WJ. 2022. Marine DNA methylation patterns are associated with microbial community composition and inform virus-host dynamics. Microbiome 10.

30. Tourancheau A, Mead EA, Zhang XS, Fang G. 2021. Discovering multiple types of DNA methylation from bacteria and microbiome using nanopore sequencing. Nat Methods 18:491–498.

31. Wilbanks EG, Doré H, Ashby MH, Heiner C, Roberts RJ, Eisen JA. 2022. Metagenomic methylation patterns resolve bacterial genomes of unusual size and structural complexity. ISME Journal 16:1921–1931.

32. Hiraoka S, Okazaki Y, Anda M, Toyoda A, Nakano S ichi, Iwasaki W. 2019. Metaepigenomic analysis reveals the unexplored diversity of DNA methylation in an environmental prokaryotic community. Nat Commun 10.

33. Li T, Zhang X, Luo F, Wu FX, Wang J. 2020. MultiMotifMaker: A multi-thread tool for identifying DNA methylation motifs from Pacbio reads. IEEE/ACM Trans Comput Biol Bioinform 17:220–225.

34. Partanen V, Dekić Rozman S, Karkman A, Muurinen J, Hiltunen T, Virta M. 2025. Tracking horizontal gene transfer of antimicrobial resistance genes in microbial community with sequence barcodes. ISME J 5.

35. Hogle SL, Tamminen M, Hiltunen T. 2024. Complete genome sequences of 30 bacterial species from a synthetic community. Microbiol Resour Announc 13.

36. Feng X, Cheng H, Portik D, Li H. 2022. Metagenome assembly of high-fidelity long reads with hifiasm-meta. Nat Methods 19:671–674.

37. Mölder F, Jablonski KP, Letcher B, Hall MB, Tomkins-Tinch CH, Sochat V, Forster J, Lee S, Twardziok SO, Kanitz A, Wilm A, Holtgrewe M, Rahmann S, Nahnsen S, Köster J. 2021. Sustainable data analysis with Snakemake. F1000Res 10:33.

38. Pacific Biosciences. ipdsummary -a tool to detect dna base-modifications from kinetic signatures. https://www.pacb.com/support/software-downloads/. Pacific Biosciences.

39. Pedregosa F, Varoquaux G, Gramfort A, Michel V, Thirion B, Grisel O, Blondel M, Prettenhofer P, Weiss R, Dubourg V, Vanderplas J, Passos A, Cournapeau D. 2011. Scikit-learn: Machine Learning in Python. Journal of Machine Learning Research 12:2825–2830.

40. McInnes L, Healy J, Melville J. 2018. UMAP: Uniform Manifold Approximation and Projection for Dimension Reduction 10.48550/arXiv.1802.03426.

41. Chklovski A, Parks DH, Woodcroft BJ, Tyson GW. 2023. CheckM2: a rapid, scalable and accurate tool for assessing microbial genome quality using machine learning. Nat Methods 20:1203–1212.

42. Parks DH, Chuvochina M, Rinke C, Mussig AJ, Chaumeil PA, Hugenholtz P. 2022. GTDB: An ongoing census of bacterial and archaeal diversity through a phylogenetically consistent, rank normalized and complete genome-based taxonomy. Nucleic Acids Res 50:D785–D794.

43. Schneider TD, Stephens RM. Sequence logos: a new way to display consensus sequences. Nucleic Acids Res 18:6097.

44. Bortolaia V, Kaas RS, Ruppe E, Roberts MC, Schwarz S, Cattoir V, Philippon A, Allesoe RL, Rebelo AR, Florensa AF, Fagelhauer L, Chakraborty T, Neumann B, Werner G, Bender JK, Stingl K, Nguyen M, Coppens J, Xavier BB, Malhotra-Kumar S, Westh H, Pinholt M, Anjum MF, Duggett NA, Kempf I, Nykäsenoja S, Olkkola S, Wieczorek K, Amaro A, Clemente L, Mossong J, Losch S, Ragimbeau C, Lund O, Aarestrup FM. 2020. ResFinder 4.0 for predictions of phenotypes from genotypes. J Antimicrob Chemother 75:3491–3500.

45. Berglund F, Österlund T, Boulund F, Marathe NP, Larsson DGJ, Kristiansson E. 2019. Identification and reconstruction of novel antibiotic resistance genes from metagenomes. Microbiome 7.

46. Camacho C, Coulouris G, Avagyan V, Ma N, Papadopoulos J, Bealer K, Madden TL. 2009. BLAST+: architecture and applications. BMC Bioinformatics 10:421.

47. Shen W, Le S, Li Y, Hu F. 2016. SeqKit: A cross-platform and ultrafast toolkit for FASTA/Q file manipulation. PLoS One 11.

48. Camargo AP, Roux S, Schulz F, Babinski M, Xu Y, Hu B, Chain PSG, Nayfach S, Kyrpides NC. 2023. Identification of mobile genetic elements with geNomad. Nat Biotechnol 42:1303–1312.

49. Shao M, Ying N, Liang Q, Ma N, Leptihn S, Yu Y, Chen H, Liu C, Hua X. 2023. Pdif-mediated antibiotic resistance genes transfer in bacteria identified by pdifFinder. Brief Bioinform 24.

50. Schwengers O, Jelonek L, Dieckmann MA, Beyvers S, Blom J, Goesmann A. 2021. Bakta: Rapid and standardized annotation of bacterial genomes via alignment-free sequence identification. Microb Genom 7.

51. Rognes T, Flouri T, Nichols B, Quince C, Mahé F. 2016. VSEARCH: A versatile open source tool for metagenomics. PeerJ 2016.

52. Katoh K, Misawa K, Kuma K-I, Miyata T. 2002. MAFFT: a novel method for rapid multiple sequence alignment based on fast Fourier transform. Nucleic Acids Res 30:3059–66.

53. Stamatakis A. 2014. RAxML version 8: A tool for phylogenetic analysis and post-analysis of large phylogenies. Bioinformatics 30:1312–1313.

54. Letunic I, Bork P. 2021. Interactive tree of life (iTOL) v5: An online tool for phylogenetic tree display and annotation. Nucleic Acids Res 49:293–296.

55. Shimoyama Y. 2024. pyGenomeViz: A genome visualization python package for comparative genomics. https://github.com/moshi4/pyGenomeViz.

56. Siguier P, Perochon J, Lestrade L, Mahillon J, Chandler M. 2006. ISfinder: the reference centre for bacterial insertion sequences. Nucleic Acids Res 34.

57. Pereira MB, Wallroth M, Kristiansson E, Axelson-Fisk M. 2016. HattCI: Fast and Accurate attC site Identification Using Hidden Markov Models. J Comput Biol 23:891–902.

58. Jumper J, Evans R, Pritzel A, Green T, Figurnov M, Ronneberger O, Tunyasuvunakool K, Bates R, Žídek A, Potapenko A, Bridgland A, Meyer C, Kohl SAA, Ballard AJ, Cowie A, Romera-Paredes B, Nikolov S, Jain R, Adler J, Back T, Petersen S, Reiman D, Clancy E, Zielinski M, Steinegger M, Pacholska M, Berghammer T, Bodenstein S, Silver D, Vinyals O, Senior AW, Kavukcuoglu K, Kohli P, Hassabis D. 2021. Highly accurate protein structure prediction with AlphaFold. Nature 596:583–589.

59. van Kempen M, Kim SS, Tumescheit C, Mirdita M, Lee J, Gilchrist CLM, Söding J, Steinegger M. 2024. Fast and accurate protein structure search with Foldseek. Nat Biotechnol 42:243–246.

60. Shaw J, Yu YW. 2025. Rapid species-level metagenome profiling and containment estimation with sylph. Nat Biotechnol 43:1348–1359.

61. Kruchten N, Seier A, Parmer C. 2025. An interactive, open-source, and browser-based graphing library for Python. https://github.com/plotly/plotly.py.

62. RStudio Team. Inc. Boston. 2016. Integrated development for R. RStudio. http://www.rstudio.com/.

63. McMurdie PJ, Holmes S. 2013. Phyloseq: An R Package for Reproducible Interactive Analysis and Graphics of Microbiome Census Data. PLoS One 8.

64. Wickham H. 2016. ggplot2: Elegant graphics for data analysis. https://ggplot2.tidyverse.org.

65. Oksanen J, Blanchet FG, Friendly M, Kindt R, Legendre P, Mcglinn D, Minchin PR, O’hara RB, Simpson GL, Solymos P, Henry M, Stevens H, Szoecs E, Wagner H. 2020. vegan: Community Ecology Package. https://cran.r-project.org/package=vegan.

66. On SLW. 2025. International Committee on Systematics of Prokaryotes (ICSP) Subcommittee on the Taxonomy of Campylobacter and related bacteria: minutes of the closed meetings, 17 January and 31 January 2024. Int J Syst Evol Microbiol 75.

67. Jain C. 2008. The E. coli RhlE RNA helicase regulates the function of related RNA helicases during ribosome assembly. RNA 14:381–389.

68. Poolman B;, Royer TJ, Mainzer SE, Schmidt BF. 1990. Carbohydrate Utilization in Streptococcus thermophilus: Characterization of the Genes for Aldose 1-Epimerase (Mutarotase) and UDPglucose 4-Epimerase. J Bacteriol 172:4037–3047.

69. Romsang A, Atichartpongkul S, Trinachartvanit W, Vattanaviboon P, Mongkolsuk S. 2013. Gene expression and physiological role of Pseudomonas aeruginosa methionine sulfoxide reductases during oxidative stress. J Bacteriol 195:3299–3308.

70. Jiang X, Hall AB, Xavier RJ, Alm EJ. 2019. Comprehensive analysis of chromosomal mobile genetic elements in the gut microbiome reveals phylum-level niche-adaptive gene pools. PLoS One 14.

71. Guo Z, Qin X, Yue M, Wu L, Li N, Su J, Jiang M. 2025. IS26 carrying blaKPC−2 mediates carbapenem resistance heterogeneity in extensively drug-resistant Klebsiella pneumoniae isolated from clinical sites. Mob DNA 16.

72. Targant H, Doublet B, Aarestrup FM, Cloeckaert A, Madec JY. 2010. IS6100-mediated genetic rearrangement within the complex class 1 integron In104 of the Salmonella genomic island 1. Journal of Antimicrobial Chemotherapy 65:1543–1545.

73. Sewunet T, Razavi M, Tellapragada C, Giske CG. 2025. Silent spread of mcr-9 in ESBL-producing Enterobacteriaceae clinical isolates, Jimma, Ethiopia. PLoS One 20.

74. Schmid S, Berger B, Haas D. 1999. Target joining of duplicated Insertion Sequence IS21 is assisted by IstB protein in vitro. J Bacteriol 181:2286–2289.

75. Díaz-García C, Sánchez-Osuna M, Serra-Compte A, Karakatsanidou I, Gómez-Sánchez I, Fidalgo B, Barbuzana-Armas C, Fittipaldi M, Rosselli R, Vinyoles J, González S, Pich OQ, Espasa M, Yáñez MA. 2025. Mapping antimicrobial resistance landscape at a city scale sewage network. Science of the Total Environment 974.

76. Dekić Rozman S, Puljko A, Karkman A, Virta M, Udiković-Kolić N. 2024. Bacterial hosts of clinically significant beta-lactamase genes in Croatian wastewaters. FEMS Microbiol Ecol 100.

77. Hultman J, Tamminen M, Pärnänen K, Cairns J, Karkman A, Virta M. 2018. Host range of antibiotic resistance genes in wastewater treatment plant influent and effluent. FEMS Microbiol Ecol 94.

78. Buzzanca D, Kerkhof PJ, Alessandria V, Rantsiou K, Houf K. 2023. Arcobacteraceae comparative genome analysis demonstrates genome heterogeneity and reduction in species isolated from animals and associated with human illness. Heliyon 9.

79. Pishgar R, Dominic JA, Sheng Z, Tay JH. 2019. Denitrification performance and microbial versatility in response to different selection pressures. Bioresour Technol 281:72–83.

80. Buzzanca D, Chiarini E, Alessandria V. 2024. Arcobacteraceae: An Exploration of Antibiotic Resistance Featuring the Latest Research Updates. Antibiotics 13.

81. Heo H, Nguyen-Dinh T, Jung MY, Greening C, Yoon S. 2025. Hydrogen-dependent dissimilatory nitrate reduction to ammonium enables growth of Campylobacterota isolates. ISME Journal 19.

82. Jia B, Raphenya AR, Alcock B, Waglechner N, Guo P, Tsang KK, Lago BA, Dave BM, Pereira S, Sharma AN, Doshi S, Courtot M, Lo R, Williams LE, Frye JG, Elsayegh T, Sardar D, Westman EL, Pawlowski AC, Johnson TA, Brinkman FSL, Wright GD, McArthur AG. 2017. CARD 2017: Expansion and model-centric curation of the comprehensive antibiotic resistance database. Nucleic Acids Res 45:D566–D573.

83. Balalovski P, Grainge I. 2020. Mobilization of pdif modules in Acinetobacter: A novel mechanism for antibiotic resistance gene shuffling? Mol Microbiol 114:699–709.

84. Mindlin S, Beletsky A, Mardanov A, Petrova M. 2019. Adaptive dif modules in permafrost strains of acinetobacter lwoffiiand their distribution and abundance among present day acinetobacter strains. Front Microbiol 10.

85. Blackwell GA, Hall RM. 2017. The tet39 determinant and the msrE-mphE genes in Acinetobacter plasmids are each part of discrete modules flanked by inversely oriented pdif (XerC-XerD) sites. Antimicrob Agents Chemother 61.

86. Girlich D, Bonnin RA, Bogaerts P, De Laveleye M, Huang DT, Dortet L, Glaser P, Glupczynski Y, Naas T. 2017. Chromosomal amplification of the blaOXA-58 carbapenemase gene in a Proteus mirabilis clinical isolate. Antimicrob Agents Chemother 61.

87. Tempel S, Bedo J, Talla E. 2022. From a large-scale genomic analysis of insertion sequences to insights into their regulatory roles in prokaryotes. BMC Genomics 23.

88. Liu H, Moran RA, Chen Y, Doughty EL, Hua X, Jiang Y, Xu Q, Zhang L, Blair JMA, McNally A, Van Schaik W, Yu Y. 2021. Transferable Acinetobacter baumannii plasmid pDETAB2 encodes OXA-58 and NDM-1 and represents a new class of antibiotic resistance plasmids. Journal of Antimicrobial Chemotherapy 76:1130–1134.

89. Geneva: World Health Organization; 2024. WHO Bacterial Priority Pathogens List, 2024: Bacterial pathogens of public health importance to guide research, development and strategies to prevent and control antimicrobial resistance.

90. Lu S, Ryu SH, Chung BS, Chung YR, Park W, Jeon CO. 2007. Simplicispira limi sp. nov., isolated from activated sludge. Int J Syst Evol Microbiol 57:31–34.

91. Peng C, Gao Y, Fan X, Peng P, Huang H, Zhang X, Ren H. 2019. Enhanced biofilm formation and denitrification in biofilters for advanced nitrogen removal by rhamnolipid addition. Bioresour Technol 287.

92. Liu W, Liu X, Liao J, Zhang Y, Liang X. 2010. Identification of bla OXA-128 and bla OXA-129, two novel OXA-type extended-spectrum-β-lactamases in Pseudomonas aeruginosa, in Hunan Province, China. J Basic Microbiol 50.

93. Valiatti TB, Streling AP, Cayô R, Santos FF, Almeida MS, Tonini MAL, Gales AC. 2024. Genomic analysis of a Pseudomonas aeruginosa strain harbouring blaKPC-2 and blaOXA-129 belonging to high-risk clone ST235 in Brazil. J Glob Antimicrob Resist 37:69–71.

94. Gillings MR. 2017. Class 1 integrons as invasive species. Curr Opin Microbiol 38:10–15.

95. Johansson MHK, Petersen TN, Nag S, Lagermann TMR, Birkedahl LEK, Tafaj S, Bradbury S, Collignon P, Daley D, Dougnon V, Fabiyi K, Coulibaly B, Dembélé R, Magloire N, Ouindgueta IJ, Hossain ZZ, Begoum A, Donchev D, Diggle M, Turnbull LA, Lévesque S, Berlinger L, Søgaard KK, Guevara PD, Duarte C, Maikanti P, Amlerova J, Drevinek P, Tkadlec J, Dilas M, Kaasch A, Westh HT, Bachtarzi MA, Amhis W, Salazar CES, Villacis JE, Lúzon MAD, Palau DB, Duployez C, Paluche M, Asante-Sefa S, Møller M, Ip M, Mareković I, Pál-Sonnevend A, Cocuzza CE, Dambrauskiene A, Macanze A, Cossa A, Mandomando I, Nwajiobi-Princewill P, Okeke IN, Kehinde AO, Adebiyi I, Akintayo I, Popoola O, Onipede A, Blomfeldt A, Nyquist NE, Bocker K, Ussher J, Ali A, Ullah N, Khan H, Gustafson NW, Jarrar I, Al-Hamad A, Luvira V, Paveenkittiporn W, Baran I, Mwansa JCL, Sikakwa L, Yamba K, Aarestrup FM. 2025. Investigation of mobile genetic elements and their association with antibiotic resistance genes in clinical pathogens worldwide. PLoS One 20.

96. Johansson MHK, Aarestrup FM, Petersen TN. 2023. Importance of mobile genetic elements for dissemination of antimicrobial resistance in metagenomic sewage samples across the world. PLoS One 18.

97. Lenferink WB, van Alen TA, Jetten MSM, Op den Camp HJM, van Kessel MAHJ, Lücker S. 2024. Genomic analysis of the class Phycisphaerae reveals a versatile group of complex carbon-degrading bacteria. Antonie van Leeuwenhoek, International Journal of General and Molecular Microbiology 117.

98. Hamidian M, Holt KE, Hall RM. 2015. The complete sequence of Salmonella genomic island SGI2. Journal of Antimicrobial Chemotherapy 70:617–619.

99. Yan W, Hall AB, Jiang X. 2022. Bacteroidales species in the human gut are a reservoir of antibiotic resistance genes regulated by invertible promoters. NPJ Biofilms Microbiomes 8.

100. Owusu-Agyeman I, Plaza E, Cetecioglu Z. 2022. Long-term alkaline volatile fatty acids production from waste streams: Impact of pH and dominance of Dysgonomonadaceae. Bioresour Technol 346.

101. Hirakata Y, Mei R, Morinaga K, Katayama T, Tamaki H, Meng X ying, Watari T, Yamaguchi T, Hatamoto M, Nobu MK. 2023. Identification and cultivation of anaerobic bacterial scavengers of dead cells. ISME Journal 17:2279–2289.

102. Ueki A, Akasaka H, Suzuki D, Ueki K. 2006. Paludibacter propionicigenes gen. nov., sp. nov., a novel strictly anaerobic, Gram-negative, propionate-producing bacterium isolated from plant residue in irrigated rice-field soil in Japan. Int J Syst Evol Microbiol 56:39–44.

103. Ma J, Sun H, Li B, Wu B, Zhang X, Ye L. 2024. Horizontal transfer potential of antibiotic resistance genes in wastewater treatment plants unraveled by microfluidic-based mini-metagenomics. J Hazard Mater 465.

104. Wen J, Zhang H, Chu D, Chen X, Feng J, Wang Y, Liu G, Zhang Y, Li Y, Ning K. 2024. Deep learning revealed the distribution and evolution patterns for invertible promoters across bacterial lineages. Nucleic Acids Res 52:12817–12830.

105. Jiang X, Hall AB, Arthur TD, Plichta DR, Covington CT, Poyet M, Crothers J, Moses PL, Tolonen AC, Vlamakis H, Alm EJ, Xavier RJ. 2019. Invertible promoters mediate bacterial phase variation, antibiotic resistance, and host adaptation in the gut. Science (1979) 363:181–187.

106. Hendriksen RS, Munk P, Njage P, van Bunnik B, McNally L, Lukjancenko O, Röder T, Nieuwenhuijse D, Pedersen SK, Kjeldgaard J, Kaas RS, Clausen PTLC, Vogt JK, Leekitcharoenphon P, van de Schans MGM, Zuidema T, de Roda Husman AM, Rasmussen S, Petersen B, Amid C, Cochrane G, Sicheritz-Ponten T, Schmitt H, Alvarez JRM, Aidara-Kane A, Pamp SJ, Lund O, Hald T, Woolhouse M, Koopmans MP, Vigre H, Petersen TN, Aarestrup FM. 2019. Global monitoring of antimicrobial resistance based on metagenomics analyses of urban sewage. Nat Commun 10:1124.

107. Karkman A, Johnson TA, Lyra C, Stedtfeld RD, Tamminen M, Tiedje JM, Virta M. 2016. High-throughput quantification of antibiotic resistance genes from an urban wastewater treatment plant. FEMS Microbiol Ecol 92.

108. Ling M, Szarvas J, Kurmauskaitė V, Kiseliovas V, Žilionis R, Avot B, Munk P, Aarestrup FM. 2024. High throughput single cell metagenomic sequencing with semi-permeable capsules: unraveling microbial diversity at the single-cell level in sewage and fecal microbiomes. Front Microbiol 15.

109. Wlodarski MA, Lau TTY, Alcock BP, Raphenya AR, Ta TE, Maguire F, Beiko RG, McArthur AG. 2025. CARD k-mers: Unmasking the pathogen hosts and genomic contexts of antimicrobial resistance genes in metagenomic sequences 10.1101/2025.09.15.676352.

110. Beaulaurier J, Schadt EE, Fang G. 2019. Deciphering bacterial epigenomes using modern sequencing technologies. Nat Rev Genet 20:157–172.

111. Ghatak S, Armstrong CM, Reed S, He Y. 2020. Comparative Methylome Analysis of Campylobacter jejuni Strain YH002 Reveals a Putative Novel Motif and Diverse Epigenetic Regulations of Virulence Genes. Front Microbiol 11.

112. Fang G, Munera D, Friedman DI, Mandlik A, Chao MC, Banerjee O, Feng Z, Losic B, Mahajan MC, Jabado OJ, Deikus G, Clark TA, Luong K, Murray IA, Davis BM, Keren-Paz A, Chess A, Roberts RJ, Korlach J, Turner SW, Kumar V, Waldor MK, Schadt EE. 2012. Genome-wide mapping of methylated adenine residues in pathogenic Escherichia coli using single-molecule real-time sequencing. Nat Biotechnol 30:1232–1239.

113. Pirone-Davies C, Hoffmann M, Roberts RJ, Muruvanda T, Timme RE, Strain E, Luo Y, Payne J, Luong K, Song Y, Tsai YC, Boitano M, Clark TA, Korlach J, Evans PS, Allard MW. 2015. Genome-wide methylation patterns in Salmonella enterica subsp. enterica Serovars. PLoS One 10.

114. Oliveira PH, Touchon M, Rocha EPC. 2014. The interplay of restriction-modification systems with mobile genetic elements and their prokaryotic hosts. Nucleic Acids Res 42:10618–10631.

115. Zorea A, Moraïs S, Pellow D, Gershoni-Yahalom O, Probst M, Nadler S, Shamir R, Rosental B, Elia N, Mizrahi I. 2026. ProFiT-SPEci-FISH: a novel approach for linking plasmids to hosts in complex microbial communities at the single-cell level. Microbiome 14.

116. Shaw LP, Rocha EPC, Maclean RC. 2023. Restriction-modification systems have shaped the evolution and distribution of plasmids across bacteria. Nucleic Acids Res 51:6806–6818.

117. Dimitriu T, Szczelkun MD, Westra ER. 2024. Various plasmid strategies limit the effect of bacterial restriction–modification systems against conjugation. Nucleic Acids Res 52:12976–12986.

118. Coluzzi C, Rocha EPC. 2025. The Spread of Antibiotic Resistance Is Driven by Plasmids Among the Fastest Evolving and of Broadest Host Range. Mol Biol Evol 42.

