## Supplementary Methods for "Long-read metagenomics and methylation-based binning allow the description of the emerging high-risk antibiotic resistance genes and their hidden hosts in complex communities"

**Synthetic community samples**

The full sample preparation and experimental setup of the synthetic communities used here for the methylation method validation are detailed in (1). Briefly, synthetic communities were composed of 36 different bacterial strains and grown in microcosm bottles for 20 days with maintenance every 96 hours by the addition of 2.5 % of the fresh community (1). The community samples were subjected to treatments involving one of five antibiotics and varying temperatures, and their combinations altogether in five replicates, reflecting the research aims of the study by Partanen and colleagues (1). Among the 75 microcosm experiments, five individual samples were included in this work to serve as a validation for the methylation-based method. These samples were bcAd1023T (0.2 µg/ml tetracycline, 15 °C; tc_0.2_15_5), bcAd1037T (20 µg/ml sulphamethazine, 25 °C; su_20_25_3), bcAd1039T (20 µg/ml sulphamethazine, 25 °C; su_20_25_4), bcAd1046T (0.02 µg/ml tetracycline, 37 °C; tc_0.02_37_3), bcAd1063T (no antibiotic, 37 °C; no_0_37_3). Since the experimental treatments did not play a critical role in our analyses, the samples were randomly selected. According to Partanen *et al.* 2025 (1), the sequencing was performed by using PacBio instrument Revio.

The available whole genome sequence data of the Partanen *et al.* 2025 (1) community members published by Hogle *et al.* 2023 (2) (<https://gitlab.utu.fi/slhogl/hambiLongRead>) was retrieved for the method validation purposes. This meant WGS data for half (18/36) of the species present in our method validation communities. The sequencing of the community members by Hogle *et al.* 2023 (2) was performed before setting up the experiments by Partanen *et al.* 2025 (1). Similarly to the wastewater metagenomes here, the WGS data by Hogle *et al.* 2023 (2) was generated by PacBio Sequel II.

**Primary analyses of the metagenomic contigs**

Assembled metagenomic contigs of the synthetic communities were queried against WGS data of the 18 strains using BLAST (3) to obtain labels for taxa and genetic element (plasmid or chromosome) for the metagenomic contigs based on the a priori knowledge of available genomes. The BLASTN results were parsed by filtering the hits by > 1000 bp and adding the columns of all taxonomic levels (from domain to species) and genetic element (plasmid or chromosome). Additionally, those with no hits were set as ‘NA’.

**Position Weight Matrices (PWM)**

The values in the PWMs represented relative frequencies of nucleotides, normalized against a background model to account for baseline nucleotide frequencies in the contig. The normalized relative frequency $S_{i}^{c}\left( a \right)$ for each nucleotide $a$ at each position $i$ (where $i \in\left[ -20,20 \right]$) within the context sequences for contig $c$ is calculated using the following formula:

$$S_{i}^{c}\left( a \right) = \log\frac{q_{i}^{c}\left( a \right)}{p^{c}\left( a \right)}$$

The term $q_{i}^{c}\left( a \right)$ represents the relative frequency of nucleotide $a$ at position $i$in the contig $c$ calculated using:

$$q_{i}^{c}\left( a \right) = \frac{N_{i}^{c}\left( a \right)+p^{c}\left( a \right)}{s+1}$$

Here, $N_{i}^{c}\left( a \right)$ is the count of nucleotide $a$ at position $i$ in the context sequences, and $s$ is the total number of bases at that position across all context sequences. The $p^{c}\left( a \right)$ is the frequency of the nucleotide $a$. By adding a pseudo count to the denominator, we avoid zero values, which would otherwise lead to undefined or infinite results when computing relative frequencies. This also helps to normalize the contributions of observed counts and background frequencies. As a result, we obtained matrices of 4 x 41 for the three modification types, allowing the analysis of methylated nucleotide positional preferences relative to their distribution throughout the contig. To minimize noise, the methylation type with fewer than 50 detected instances. Finally, the matrices were flattened into one-dimensional arrays, and the sample-wise contigs were concatenated into large files according to the three modification types. In the case of synthetic community data, the labels were attached to these files by matching the contigs’ PWM data with their corresponding taxonomic and genetic element (plasmid or chromosome) identity.

**Random Forest classifier**

A Random forest model was trained on the synthetic community dataset, which was split into 80 % and 20 % training and testing data sets, respectively, and stratified for each species in order to predict the taxonomic labels of the metagenomic contigs using the high-dimensional methylation matrix. The models were implemented with Scikit-learn (v1.4.2) (4) Python library and trained with default parameters, including the construction of 100 decision trees, each trained using bootstrap sampling, with the number of features considered at each node set to the square root of the total number of features. For Random Forest analysis, contigs with less than 10 lines of base modification data were excluded. This filtering resulted in the exclusion of 4 bacterial species from the analysis (from 15 to 11 species). To assess model performance across taxonomic levels, the classification accuracy was evaluated as the target moved from species upward to higher taxonomic ranks.

**UMAP for the method validation**

Metagenomic contigs of the synthetic community data with known taxonomic identity were color-coded to allow the evaluation of the taxa-wise clustering of contigs based on PWMs and projected by UMAP. Additionally, information on the sequence lengths computed by SeqKit (v2.5.1) (5) and the amount of base modification data obtained per contig (ipdSummary (v3.0) (6) output), was added to the preliminary visualization. For visualization purposes, the contig lengths were square root transformed, while a logarithmic transformation was applied for the numbers of detected base modifications.

**Sequence logos**

To further confirm the similar methylation profiles of clustered contigs, sequence logos were created to visualize the nucleotide preferences across the context sequences at each position (7). Sequence logos show the distribution of the four bases at each position within the sequence, aiding in understanding the nucleotide preferences by illustrating the relative frequency and importance of each base (7), in our case, revealing the methylation target motif sequences.

The information content (7) *R_i_* for each position *i* which was calculated using the formula:

$$R_{i} = \log_{2} \left( s \right) - \left( H_{i}+e_{n} \right)$$

where *s* is the number of possible nucleotides, *n* the number of sequences, and *H_i_*

is the uncertainty (or Shannon entropy) at position *i* in the context sequence:

$$H_{i} = -\sum_{b=1}^{t} f_{b,i} \times\log_{2} f_{b,i}$$

and the function *f_b,i_* is the relative frequency of nucleotide *b* in position *i*. The

*e_n_* is the approximation for the small-sample correction calculated with

$$e_{n} = \frac{1}{\text{ln}2} \times\frac{s-1}{2n}$$

In the logo image, the total height of the letters illustrates the information content

of a specific position in the sequences converted into bits:

$$height = f_{b,1} \times R_{i}$$

**Methylation-based binning for wastewater community data**

Contig clusters projected in UMAP were examined by visual inspection using an interactive scatter plot by Plotly (8) for each sample separately (**Fig. S7**). The large cluster of non-disguisable data points in each sample was excluded from the main analysis to allow a more accurate assessment of data points where distinct clustering patterns were evident (**Fig. S8)**. The excluded data represented contigs with inconclusive methylation data, restricting their eligibility by our approach, and thus, they were excluded. The interactive visualization enabled a magnified view of the clusters, which enabled the recognition of related contigs. Contig identities of the clusters were gathered by their coordinates in the UMAP. After the concatenation of the sequences, the quality and taxonomical identities were confirmed using CheckM2 (v 1.0.1) (9). GTDB-Tk (v2.4.1, database release 220) (10) respectively.

**Analysis of ARG mobility potential and genetic context**

Regarding the more detailed description of the hosts and genetic contexts of latent ARGs, predicted by fARGene (11), we decided to focus on the putative beta-lactamases due to their higher likelihood of serving the antibiotic resistance function and thus being mobile in contrast to chromosomal housekeeping genes that are incorrectly assigned as putative resistance genes (39).

The latent class C or D beta-lactamases represented by methylation-resolved genome bins were used to query similar unbinned sequences in the full dataset by BLAST (3). Bakta (v1.11.0) (12) was used for gene annotations. To extract the ARG sequences from their context, a BLAST comparison between the contigs with matches and the ORF sequences predicted by fARGene was performed to identify the sequence locations. For the analysis of the ARG flanking gene region, this location information was used to obtain sequences of 5 or 10 kb up- and downstream of the ARG for further analysis by in-house scripts applying SeqKit tools (v 2.5.1) (5).

For mobility prediction and detailed gene context analysis, flanking regions were dereplicated for the visualization. For that, vsearch (v2.22.1) (13) was run for all binned and unbinned sequences of class D and C beta-lactamases separately using sequence identity thresholds of 0.99 and 0.90 for respectively. For the established *erm*(F) the threshold for the dereplication performed by vsearch (v2.22.1) (13) was 0.50. After dereplication, all sequences that had been assigned to genome bins but were removed during dereplication were reinstated for visualization purposes. Additionally, reference sequences that either matched the predicted genes or represented the closest related established ARGs retrieved from public sequence repositories were included in the visualization along with their flanking regions. The final manual data exclusion step was performed by excluding sequences in which genes of interest were truncated by visual inspection.

In addition to *Acinetobacter* sp*,* (bin INF1-C3f), the predicted class C beta-lactamases were also hosted by *Aquabacterium_A* sp. (bins INF3_C1f, EFF1-C1f, and EFF2-C1f) and *Acinetobacter cellitus* (bin INF2-C5f) (**Table 1**). However, the beta-lactamases in the latter two hosts were dissimilar to those in *Acinetobacter* sp., which had more matching unbinned genes and mobility-associated contexts in the whole data. For this reason, we decided to focus more on the investigation of the latter genes, their contexts, and hosts.

The beta-lactamase and *erm*(F) sequences were oriented consistently (reverse-complemented if needed) for the sequence alignment and gene tree visualization with MAFFT (v7.505) (14), RAxML (v8.2.12) (15) and Interactive Tree of Life (iTOL) (16). For the genetic context visualization, GenBank files (.GBFF) obtained from Bakta (v1.11.0) (12) were converted to a consistent orientation by an in-house Python script if needed before the visualization with pyGenomeViz (v1.6.1) (17) using a BLAST (3) query for highlighting the aligned nucleotide sequences between the compared sequences. The gene colors were set manually according to the Bakta annotation labels.

The analysis for the remaining two established ARGs, *bla*OXA-129 and *sul1-9*, and their contexts was similar to the latent ARGs, except for the dereplication step, which was unnecessary because the number of visualized sequences was sufficiently low. Additionally, no gene trees for the ARGs were drawn as the sequences were identical or differed by only one to two nucleotides from each other.

**References**

1.	Partanen V, Dekić Rozman S, Karkman A, Muurinen J, Hiltunen T, Virta M. 2025. Tracking horizontal gene transfer of antimicrobial resistance genes in microbial community with sequence barcodes. ISME J 5.

2.	Hogle SL, Tamminen M, Hiltunen T. 2024. Complete genome sequences of 30 bacterial species from a synthetic community. Microbiol Resour Announc 13.

3.	Camacho C, Coulouris G, Avagyan V, Ma N, Papadopoulos J, Bealer K, Madden TL. 2009. BLAST+: architecture and applications. BMC Bioinformatics 10:421.

4.	Pedregosa F, Varoquaux G, Gramfort A, Michel V, Thirion B, Grisel O, Blondel M, Prettenhofer P, Weiss R, Dubourg V, Vanderplas J, Passos A, Cournapeau D. 2011. Scikit-learn: Machine Learning in Python. Journal of Machine Learning Research 12:2825–2830.

5.	Shen W, Le S, Li Y, Hu F. 2016. SeqKit: A cross-platform and ultrafast toolkit for FASTA/Q file manipulation. PLoS One 11.

6.	Pacific Biosciences. ipdsummary - a tool to detect dna base-modifications from kinetic signatures. https://www.pacb.com/support/software-downloads/. Pacific Biosciences.

7.	Schneider TD, Stephens RM. Sequence logos: a new way to display consensus sequences. Nucleic Acids Res 18:6097.

8.	Kruchten N, Seier A, Parmer C. 2025. An interactive, open-source, and browser-based graphing library for Python. https://github.com/plotly/plotly.py.

9.	Chklovski A, Parks DH, Woodcroft BJ, Tyson GW. 2023. CheckM2: a rapid, scalable and accurate tool for assessing microbial genome quality using machine learning. Nat Methods 20:1203–1212.

10.	Parks DH, Chuvochina M, Rinke C, Mussig AJ, Chaumeil PA, Hugenholtz P. 2022. GTDB: An ongoing census of bacterial and archaeal diversity through a phylogenetically consistent, rank normalized and complete genome-based taxonomy. Nucleic Acids Res 50:D785–D794.

11.	Berglund F, Österlund T, Boulund F, Marathe NP, Larsson DGJ, Kristiansson E. 2019. Identification and reconstruction of novel antibiotic resistance genes from metagenomes. Microbiome 7.

12.	Schwengers O, Jelonek L, Dieckmann MA, Beyvers S, Blom J, Goesmann A. 2021. Bakta: Rapid and standardized annotation of bacterial genomes via alignment-free sequence identification. Microb Genom 7.

13.	Rognes T, Flouri T, Nichols B, Quince C, Mahé F. 2016. VSEARCH: A versatile open source tool for metagenomics. PeerJ 2016.

14.	Katoh K, Misawa K, Kuma K-I, Miyata T. 2002. MAFFT: a novel method for rapid multiple sequence alignment based on fast Fourier transform. Nucleic Acids Res 30:3059–66.

15.	Stamatakis A. 2014. RAxML version 8: A tool for phylogenetic analysis and post-analysis of large phylogenies. Bioinformatics 30:1312–1313.

16.	Letunic I, Bork P. 2021. Interactive tree of life (iTOL) v5: An online tool for phylogenetic tree display and annotation. Nucleic Acids Res 49:293–296.

17.	Shimoyama Y. 2024. pyGenomeViz: A genome visualization python package for comparative genomics. https://github.com/moshi4/pyGenomeViz.
