## Supplementary Figures (Figures S1-S9) for "Long-read metagenomics and methylation-based binning allow the description of the emerging high-risk antibiotic resistance genes and their hidden hosts in complex communities"

List of Supplementary Figures:

**Fig. S1.** Bacterial composition of the synthetic community

**Fig. S2.** Random Forest classifier predictions

**Fig. S3.** Uniform Manifold Approximation and Projection (UMAP) of synthetic community methylomes.

**Fig. S4.** Sequence logos of contigs included in synthetic community bins and MAGs

**Fig. S5.** PCoA of bacterial composition in wastewater

**Fig. S6.** Bacterial composition of the wastewater community

**Fig. S7.** UMAP ordination of methylomes of wastewater communities

**Fig. S8.** *erm*(F) bins in different wastewater samples

**Fig. S9.** Composition of the Bacteroidales order in wastewater communities

**Supplementary Results**

**Method validation**

To reduce noise in the data, contigs with *a priori* known taxonomical identity (n = 18) had fewer than 10 contigs, and those lacking any methylation signals were excluded from the random forest classifier analysis. By this filtering criterion, 11 out of 18 species remained for the model. Additionally, a threshold for included base modification data per contig was explored (tested values 20, 50, 100, 200, and 500 were tested, **Table S2**) and set to >50 as this threshold cut off the noise without losing any of the eleven species shown as the highest test accuracy values with maximum included species number (**Table S2**). Accuracy test values ranged from 0.88 to 1 from species to phylum, where the number of different taxon labels varied from 11 to 1, respectively (**Table S2**), partially explaining the increasing accuracy when moving towards higher taxa levels.

The PWMs of the synthetic community metagenomes were visualized using Uniform Manifold Approximation and Projection (UMAP) (**Fig. S3A**). To test whether the contig clusters correspond to MAGs or bins, completion and redundancy of some selected species clusters were examined (**Table S3**). Fully complete MAGs containing both chromosomal and plasmid elements expected based on the WGS data were curated for multiple species and samples (**Table S3**). In the sequence logos of selected contigs included in the MAGs, the nucleotide letter is displayed larger at the central position due to the higher information content (**Fig. S4**). However, methylation site surrounding region in the ±20 bases window, variability is observed in the preferred bases for both m6A and m4C (**Fig. S4**). Methylation signatures are highly similar across chromosomal and plasmid contigs of the same species (**Fig. S4**).

For some of these species and samples, only partial bins could be gathered. Finally, some contigs originating from different species are grouped under a miscellaneous group (**Fig. S3A**). These contigs represent those with the lowest amount of methylation data (**Fig. S3B**), suggesting that a certain level of input data is required for obtaining distinctive clusters. However, it may be that some bacterial species, for example, *Cupriavidus oxalaticus* and *Comamonas testostorini_C* in the miscellaneous group, do not biologically (or have too few contigs) demonstrate any uniform methylation profile (**Fig. S2**, **Fig. S3A**)*.* But as this miscellaneous group of contigs is clearly separate from the more defined, distinct clusters, the risk of false positives coming from this group is ruled out (loss of data rather than false positives). Altogether, we conclude that species-wise bins – although not entirely complete bacterial genomes – can be correctly curated by our approach (**Fig. S3A**).

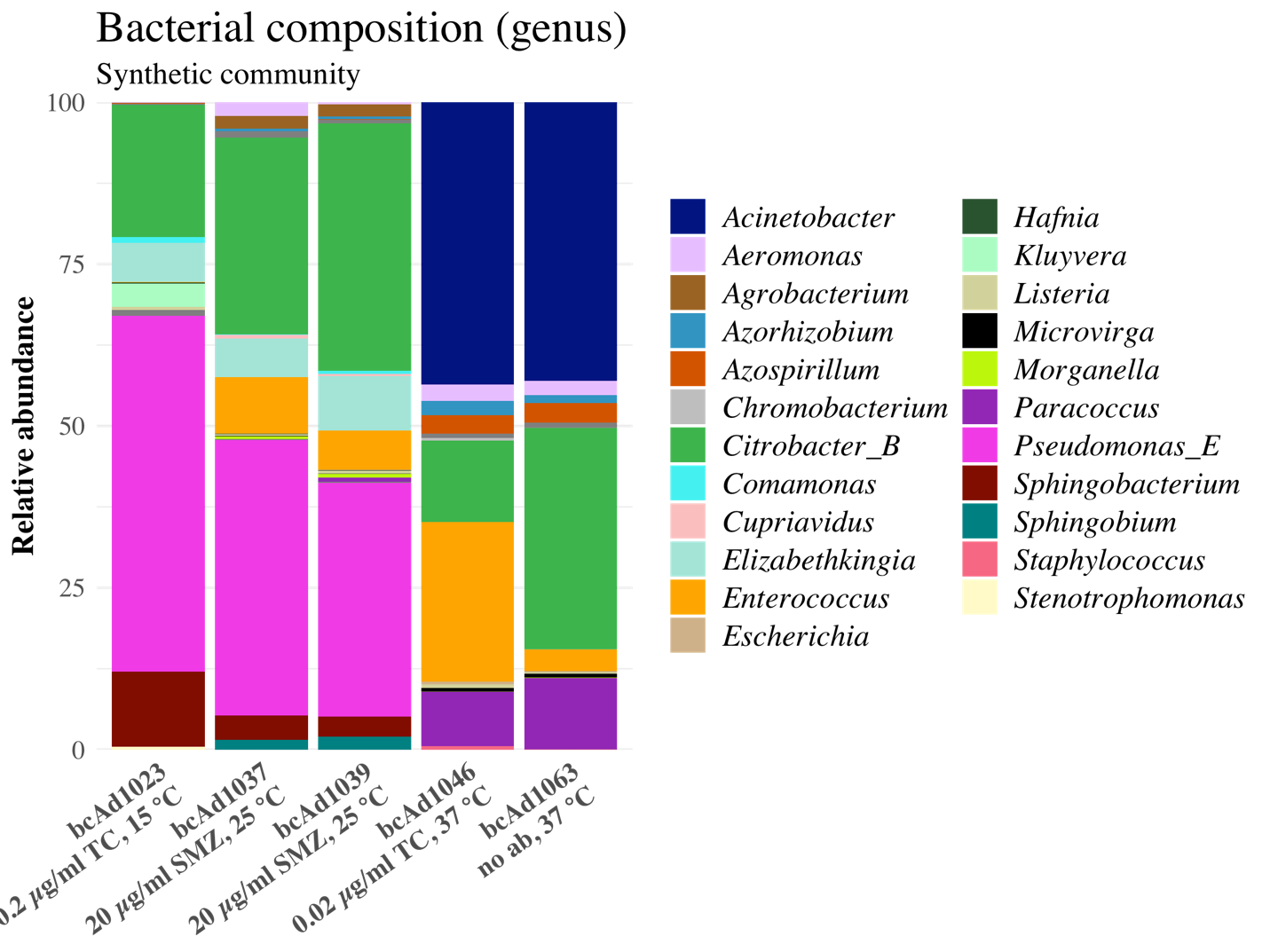
**Fig. S1.** Bacterial composition of the synthetic community. Genus-level taxonomic profiling was performed using Sylph (1).

**A. B.**

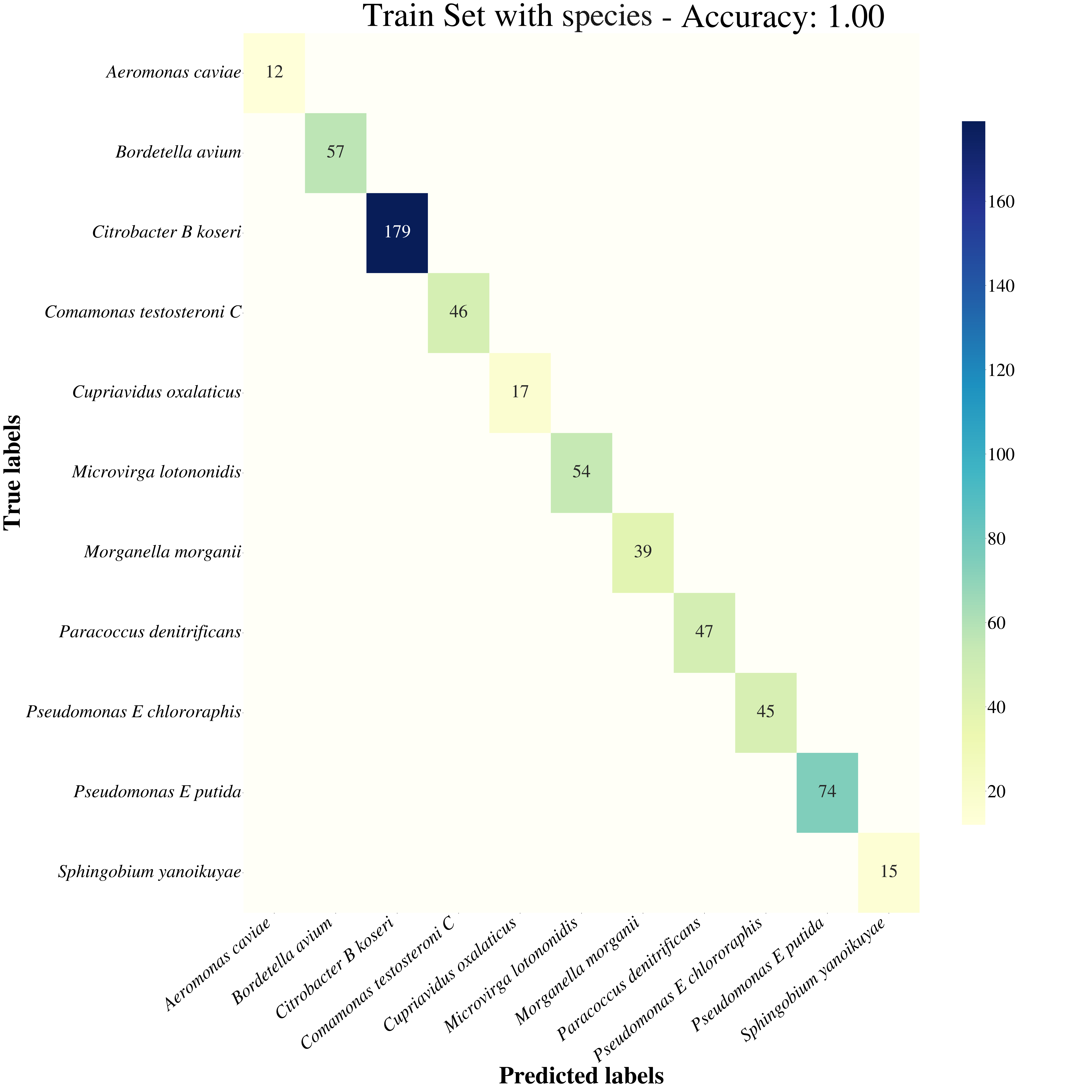

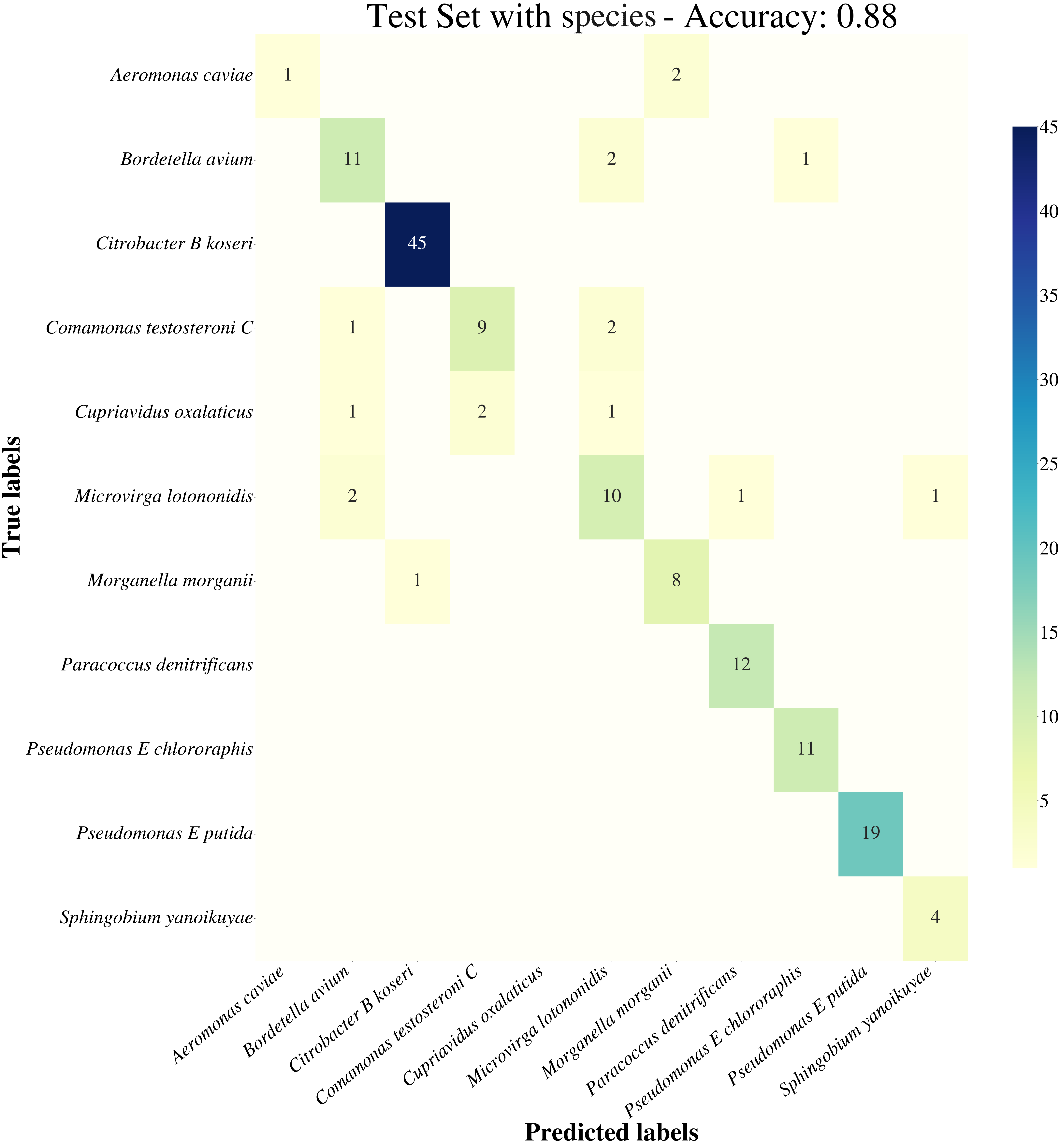

**Fig. S2.** Random Forest classifier predictions for the taxonomic labels of the metagenomic contigs using the high-dimensional methylation matrix. Predictions are shown for **A.** Train Set and **B.** Test Set.

**A.**

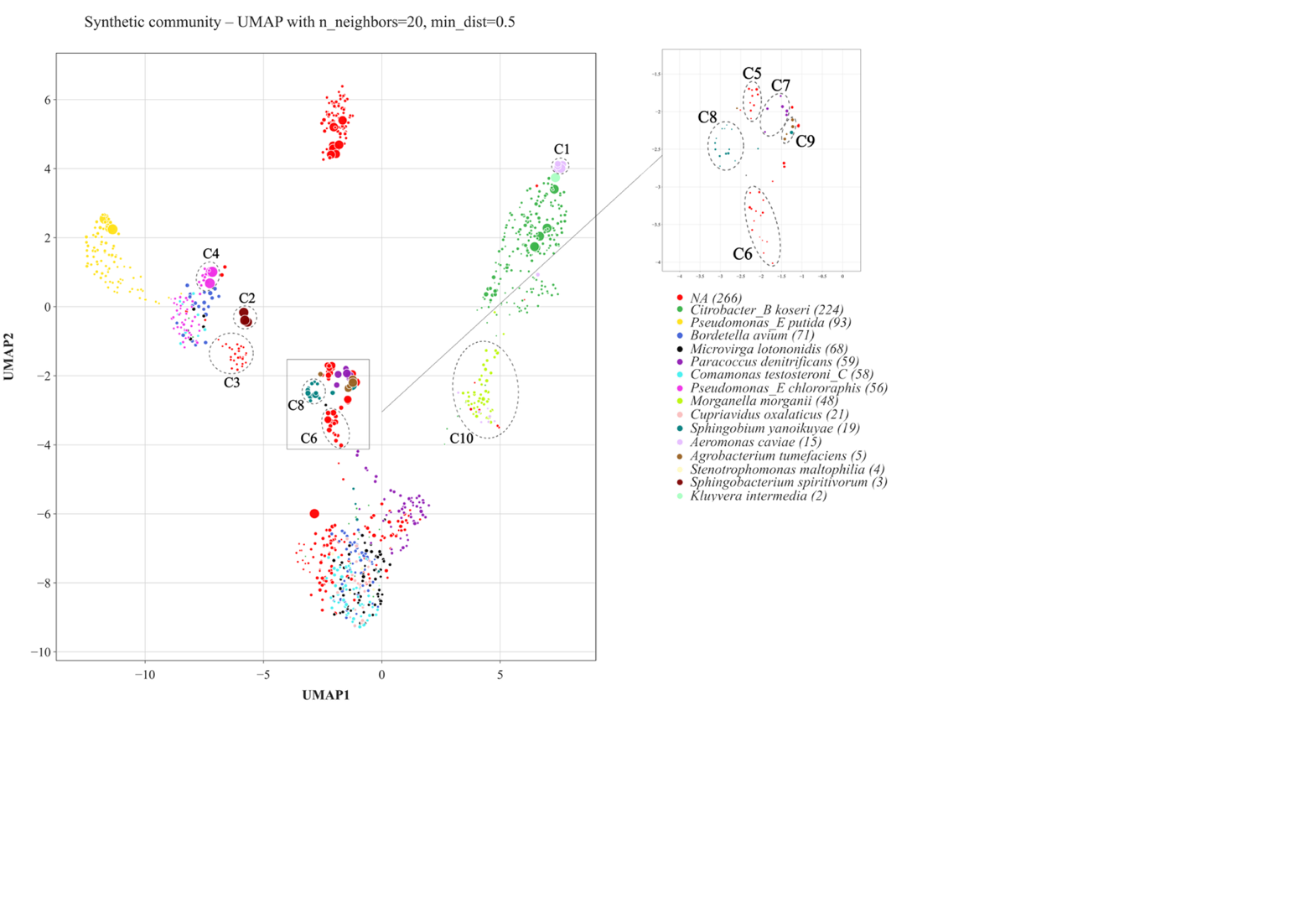

**B.**

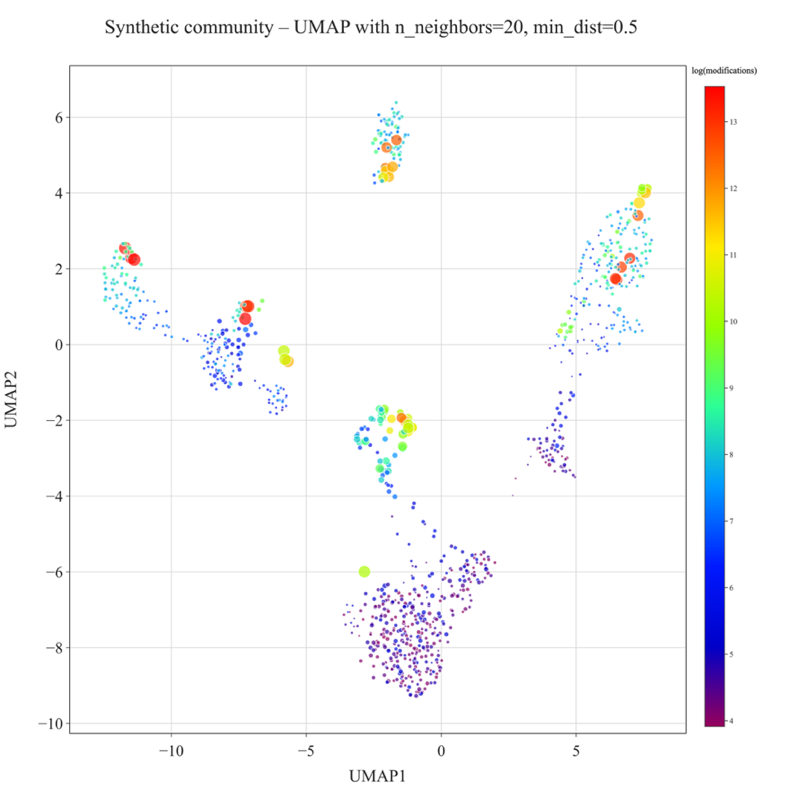

**Fig. S3.** Uniform Manifold Approximation and Projection (UMAP) of the synthetic community methylomes. **A.** The *a priori* known taxonomical identities of the metagenomic contigs are color-coded based on species-level taxa. Data point size denotes the square root transformed contig length. **B.** The amount of base modification data used for downstream analysis is displayed as heatmap colors, where blue denotes for fewest and red for the highest amount of data. The number of detected base modifications is log-transformed for the visualization. Data point size denotes the square root transformed contig length.

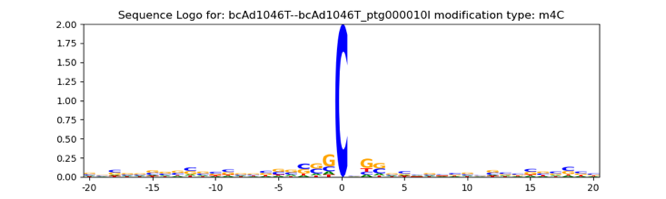

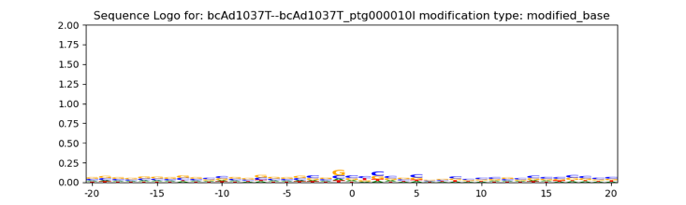

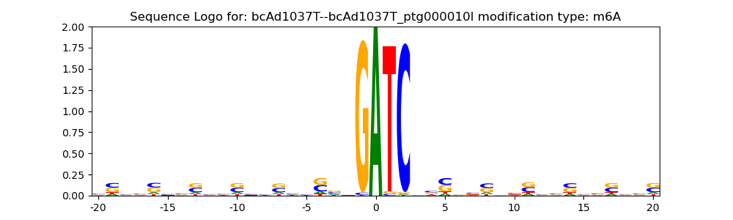

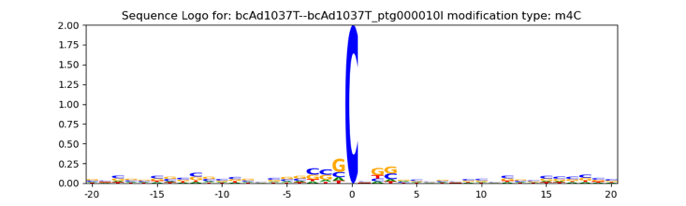

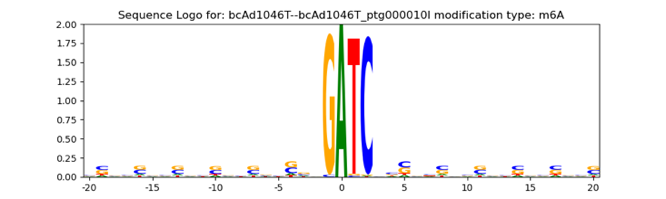

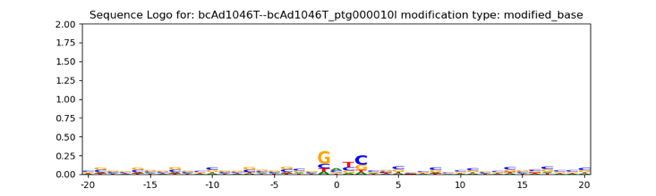
*Aeromonas caviae* (chrom)

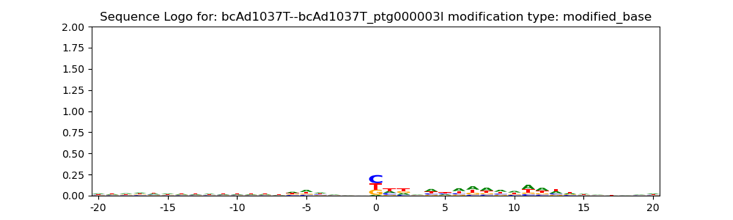

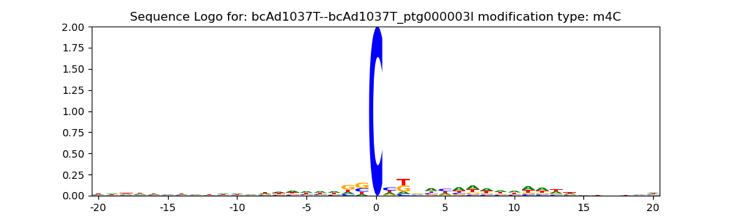

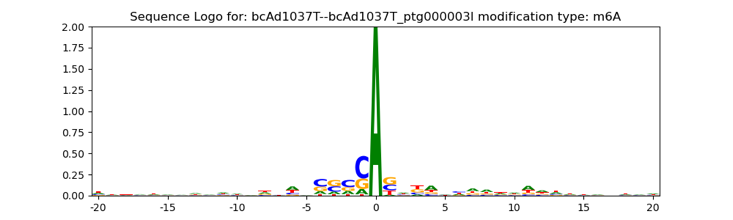

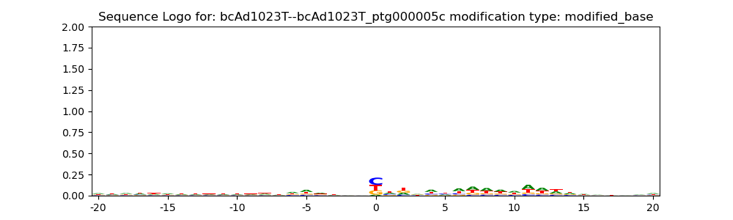

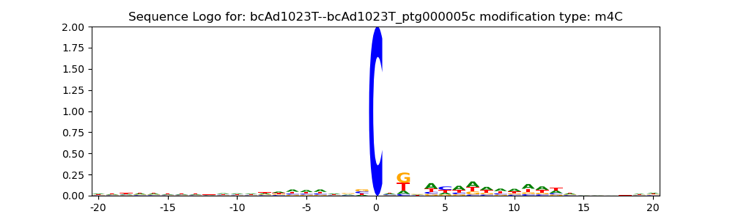

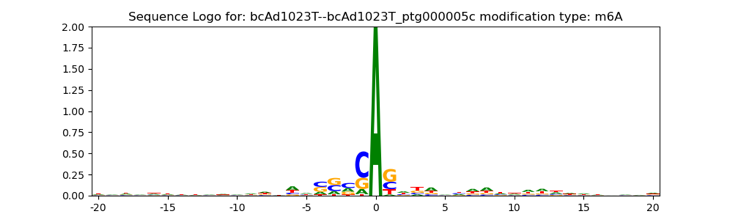
*Sphingobacterium spiritivorum* (chrom)

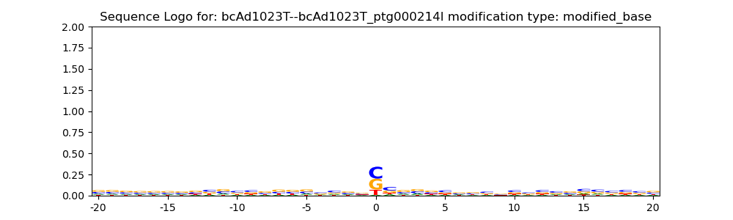

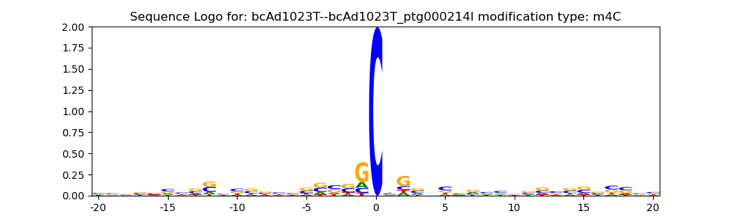

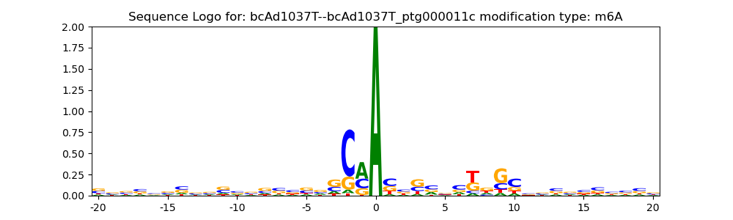

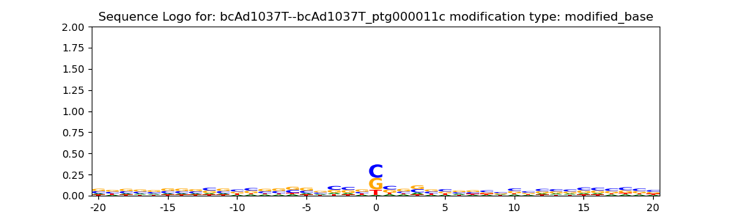

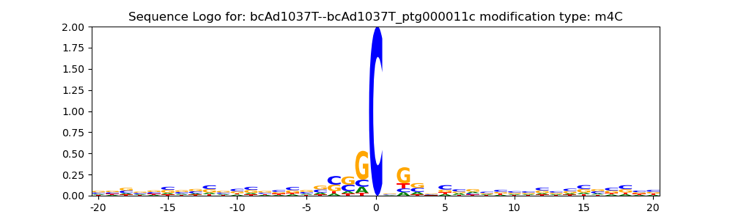

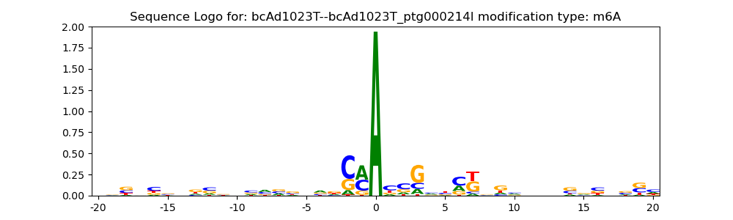
*Pseudomonas_E chlororaphis* (chrom)

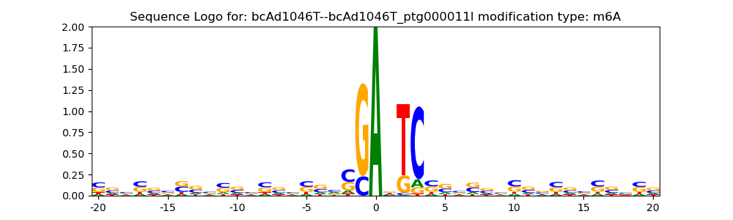

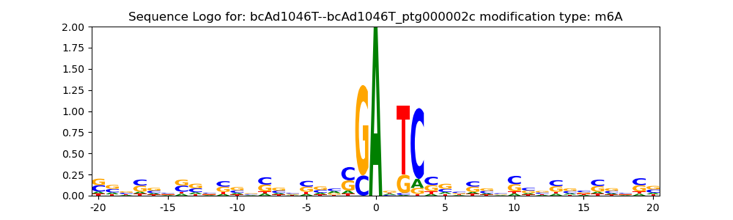
*Azospirillum brasilense_B*

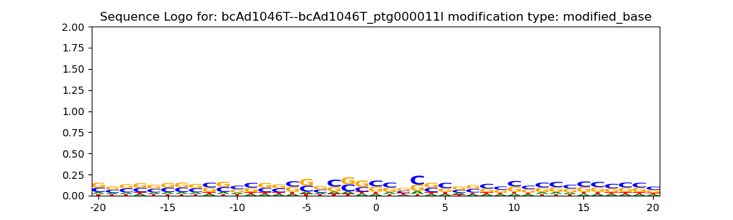

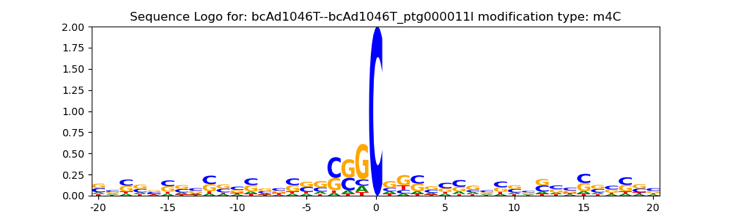

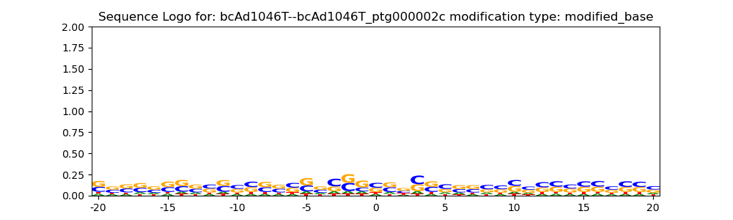

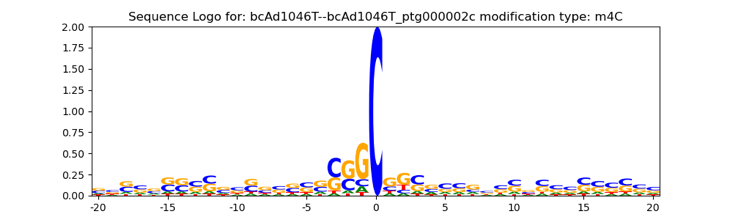

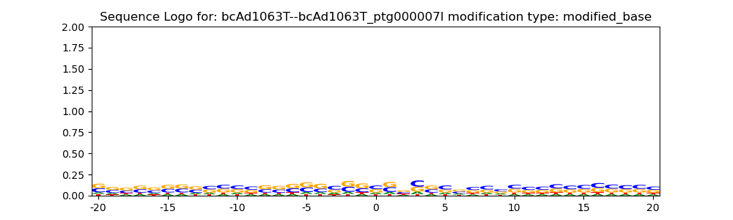

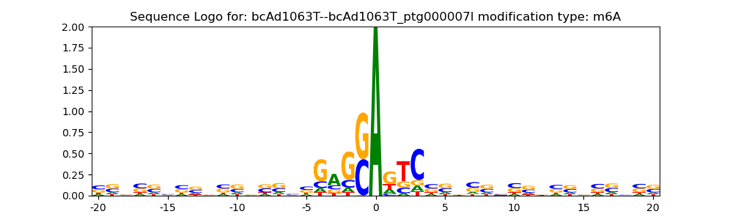

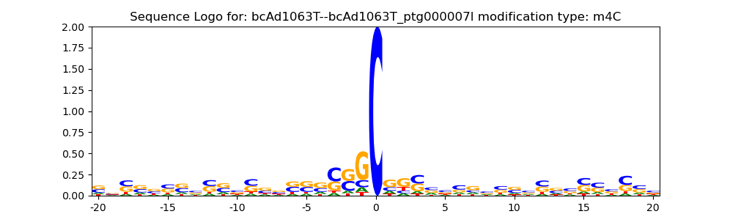

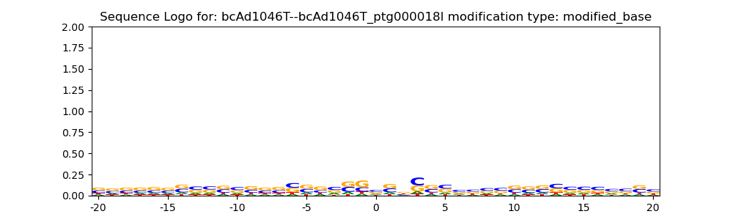

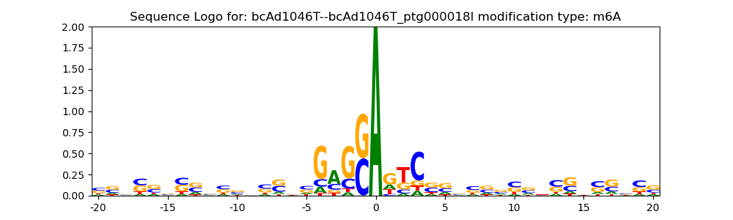

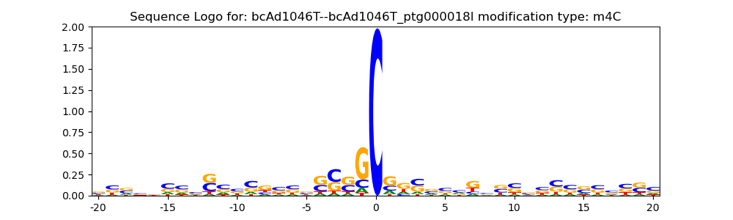
*Azorhizobium caulinodans*

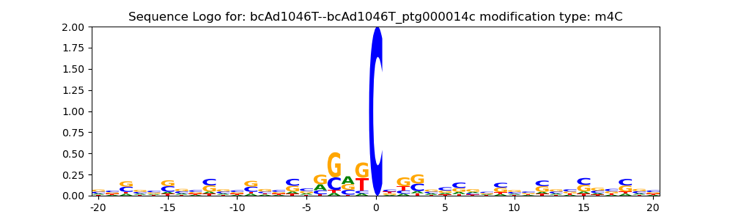

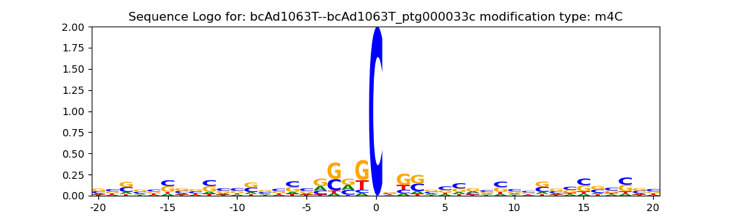

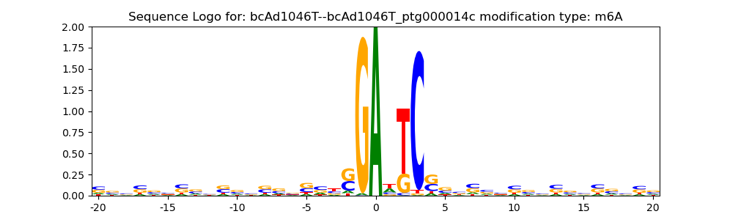

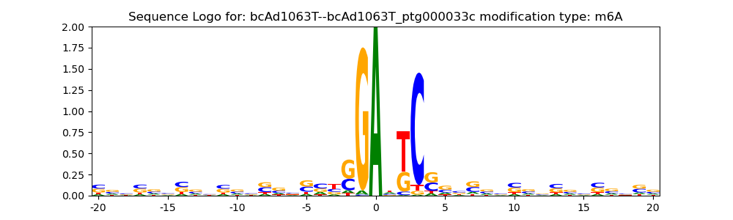
*Paracoccus denitrificans* (chrom, plas)

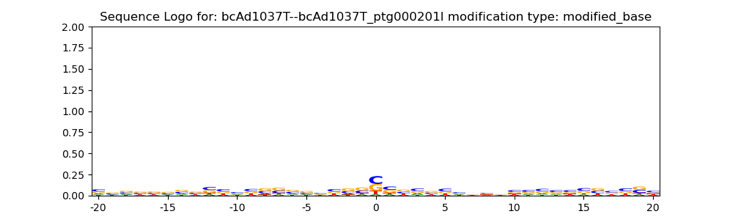

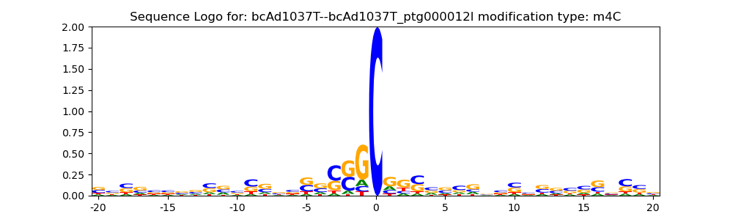

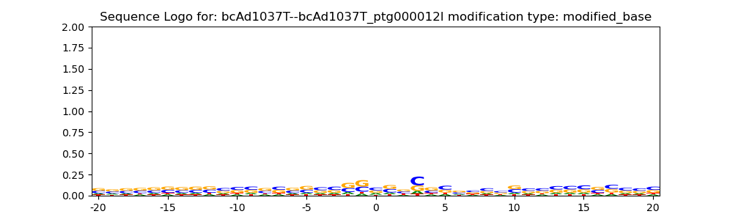

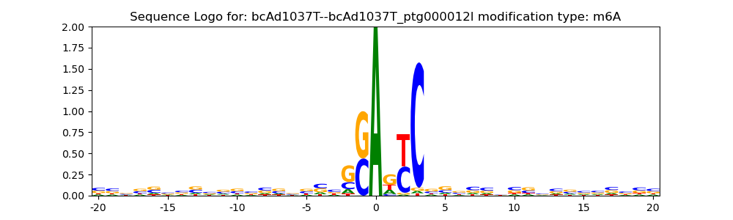

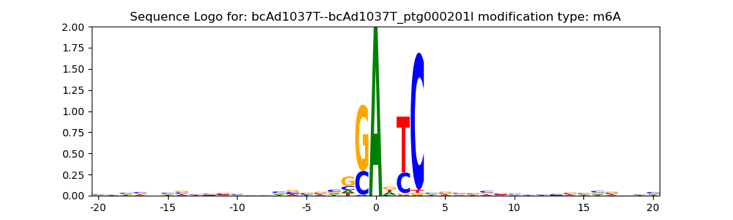

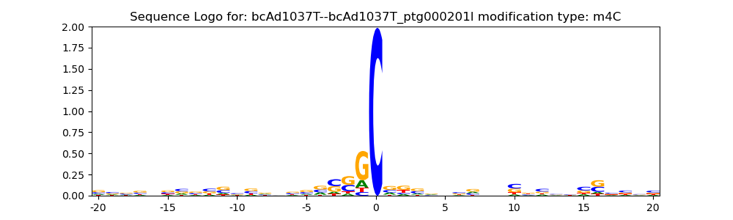
*Sphingobium yanoikuyae* (chrom, plas2)

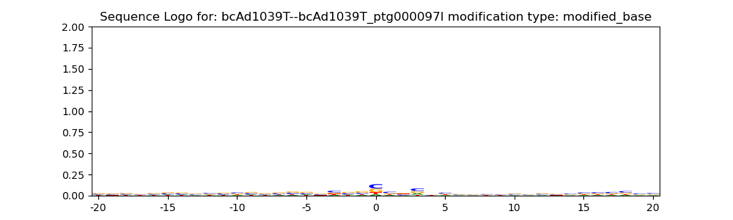

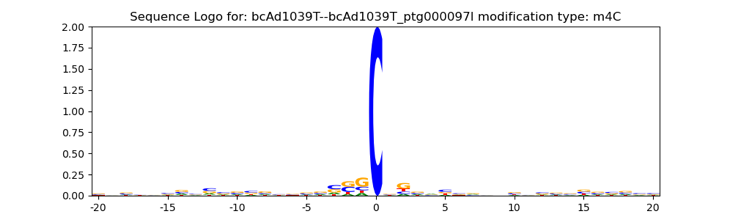

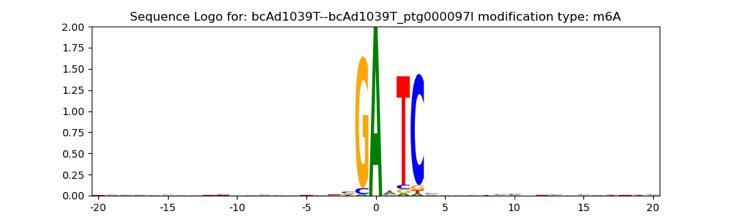

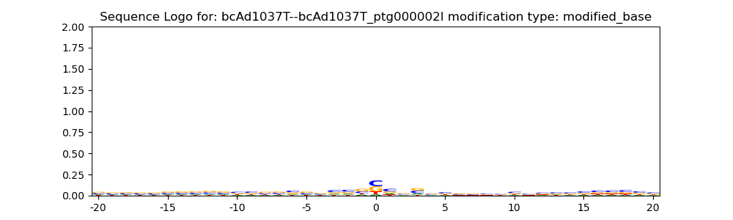

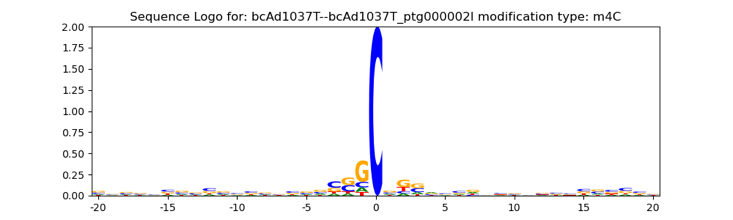

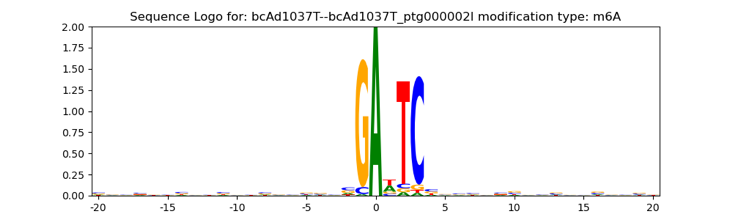
*Agrobacterium tumefaciens* (chrom2, plas)

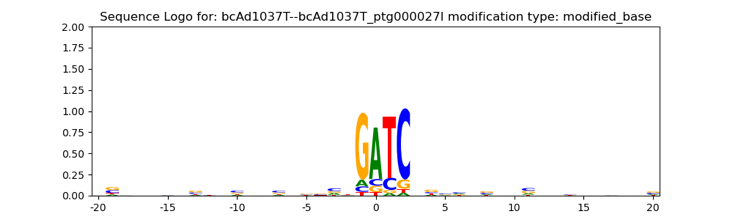

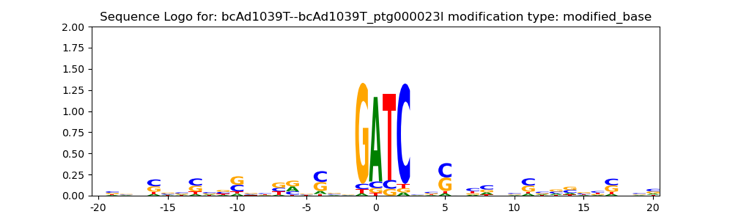
*Morganella morganii* (chrom)

**Fig. S4.** Sequence logos of contigs included in synthetic community bins and MAGs (**Fig. S3**, **Table S3**). Sequence logos of two example contigs are shown. The examples were selected based on the following criteria: contigs from separate samples and different genetic elements, if applicable (chromosomes (chrom1-2) or plasmids (plas1-2)).

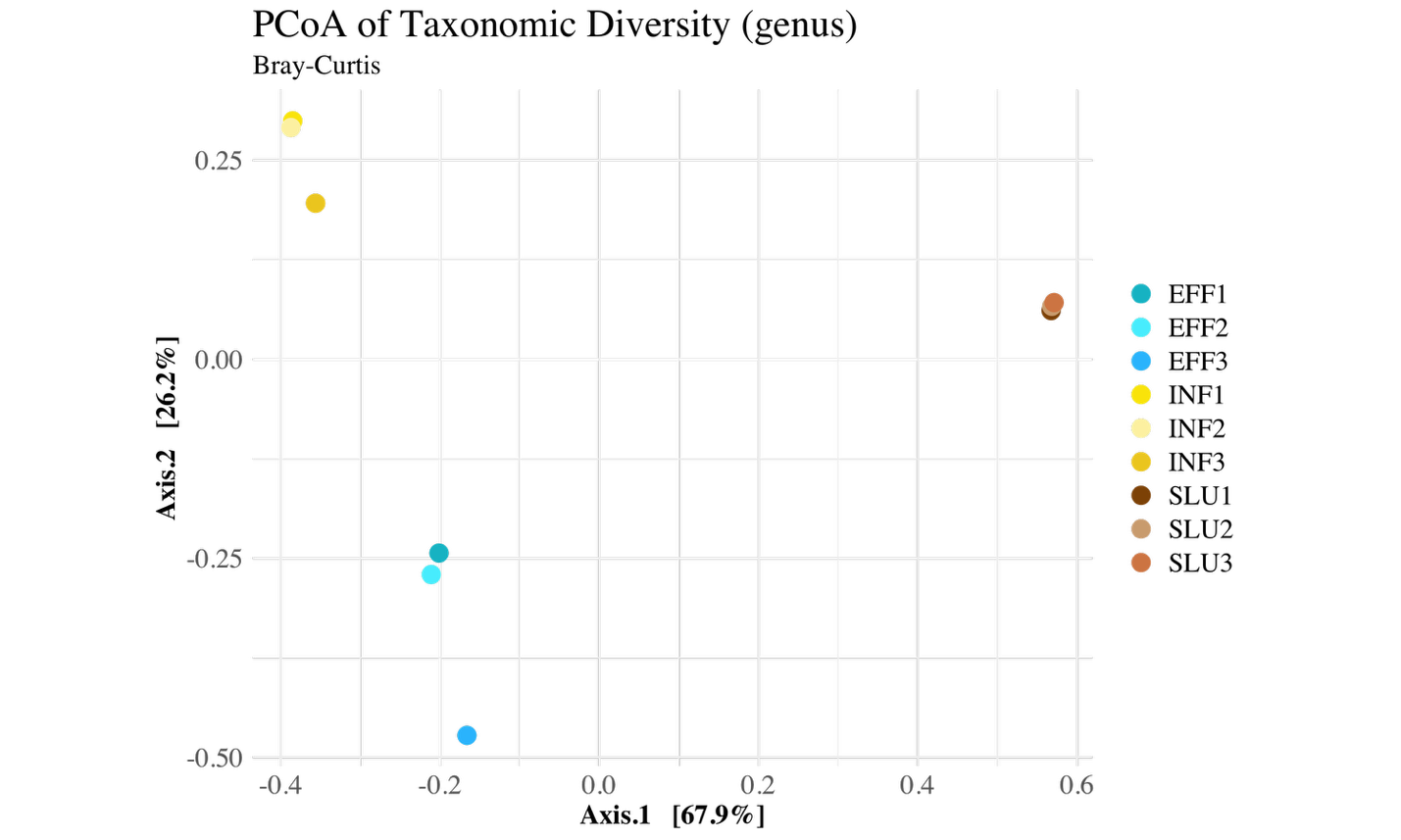

|  | Df | Sumofsqs | R2 | F | Pr(>F) |
| --- | --- | --- | --- | --- | --- |
| Model | 2 | 2.05630 | 0.92591 | 37.493 | 0.005 ** |
| Residual | 6 | 0.16453 | 0.07409 |  |  |
| Total | 8 | 2.22083 | 1.00000 |  |  |

**Fig. S5.** Principal Coordinates Analysis (PCoA) of bacterial composition in wastewater. Ordination was plotted with genus-level relative abundances generated by Sylph (1) using Bray-Curtis dissimilarity by the vegan package (2). Microbial community composition differed significantly between the sample types (p = 0.005, highly significant (p < 0.01)). F = 37.493: A high F-statistic suggested strong separation between groups. About 92.6% of the variation in Bray-Curtis distances is explained by sample type (Permutation test for adonis under reduced model, Permutation: free, Number of permutations: 999).

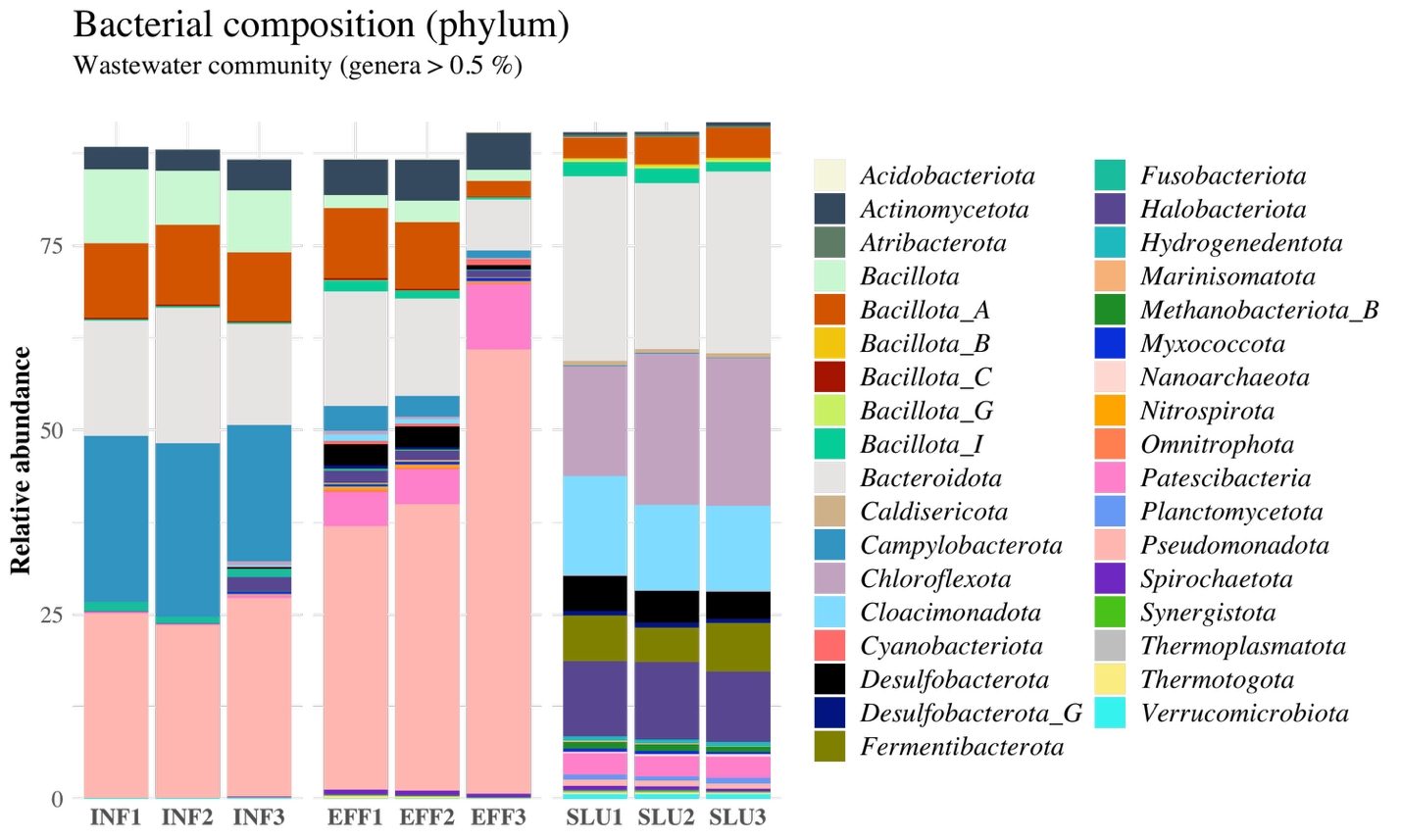

**Fig. S6.** Bacterial composition of wastewater communities. Genera with less than 0.5 % relative abundance computed by Sylph (1) were filtered out from the visualization. Phylum-level taxa are shown.

**A.	B. C.**

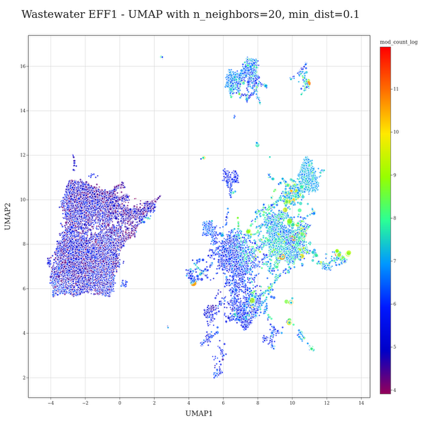

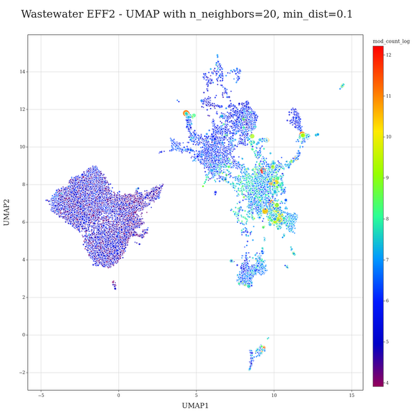

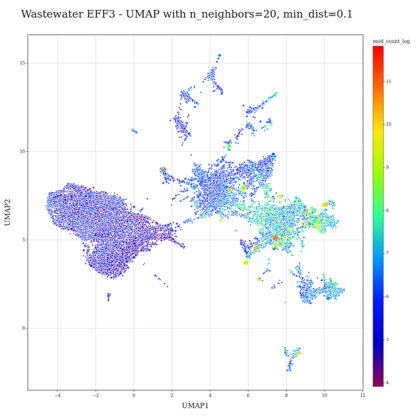

**D. E. F.**

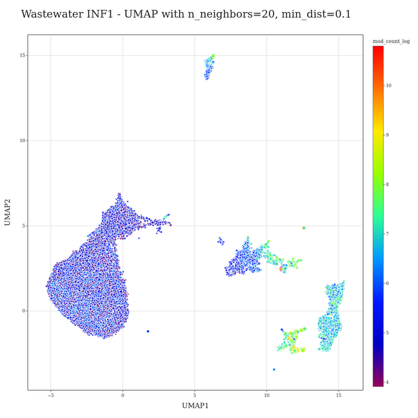

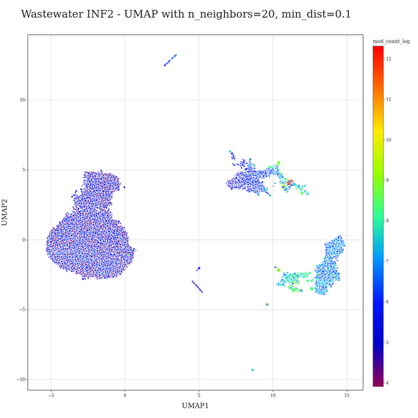

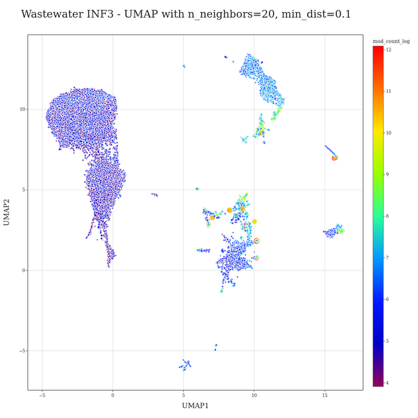

**G. H. I.**

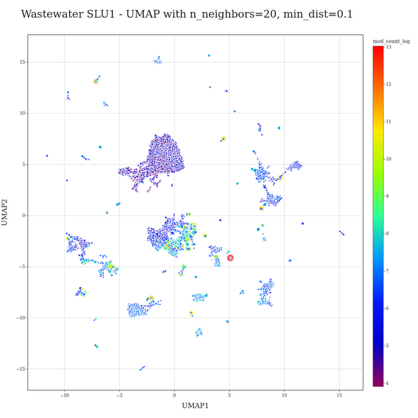

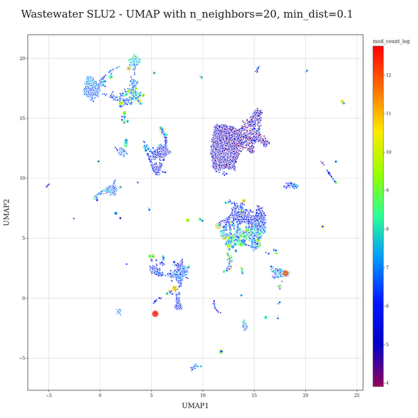

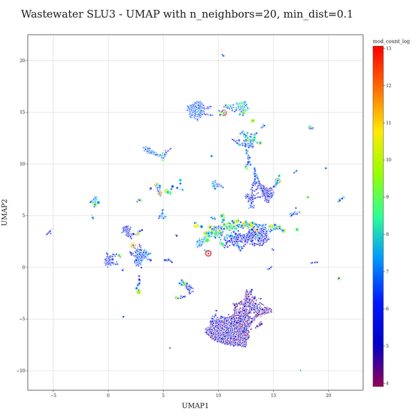

**Fig. S7.** UMAP ordination of methylomes of wastewater communities. The heatmap color encoding denotes the number of detected base modifications per contig. A logarithmic transformation was applied to the modification count data to reduce skewness. The dot size indicates the contig length, which is square root transformed. In **A-F**, the included data is shown with a rectangle with a dotted line, while in **G-I**, all data except those contigs within the cluster a marked with a red solid rectangle.

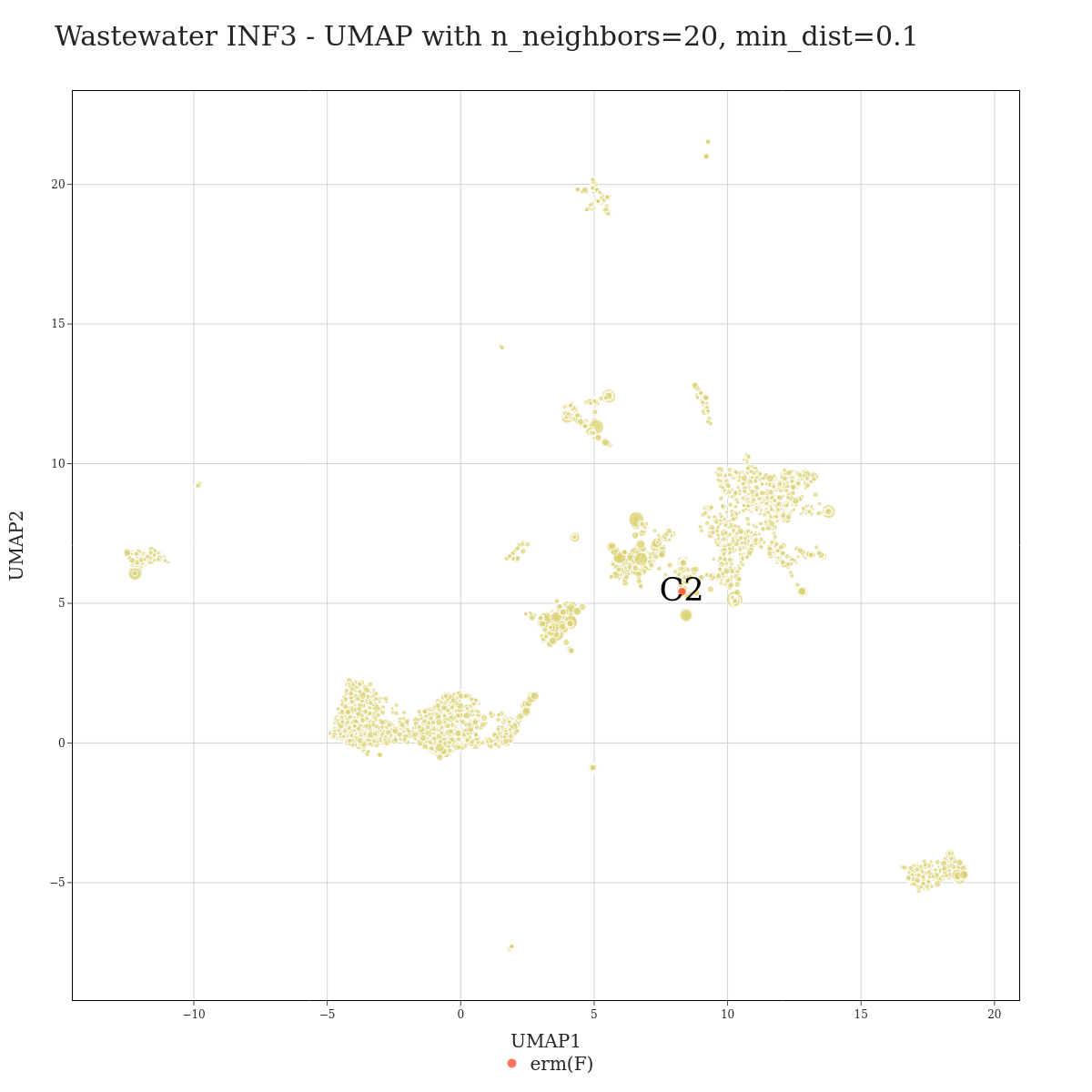

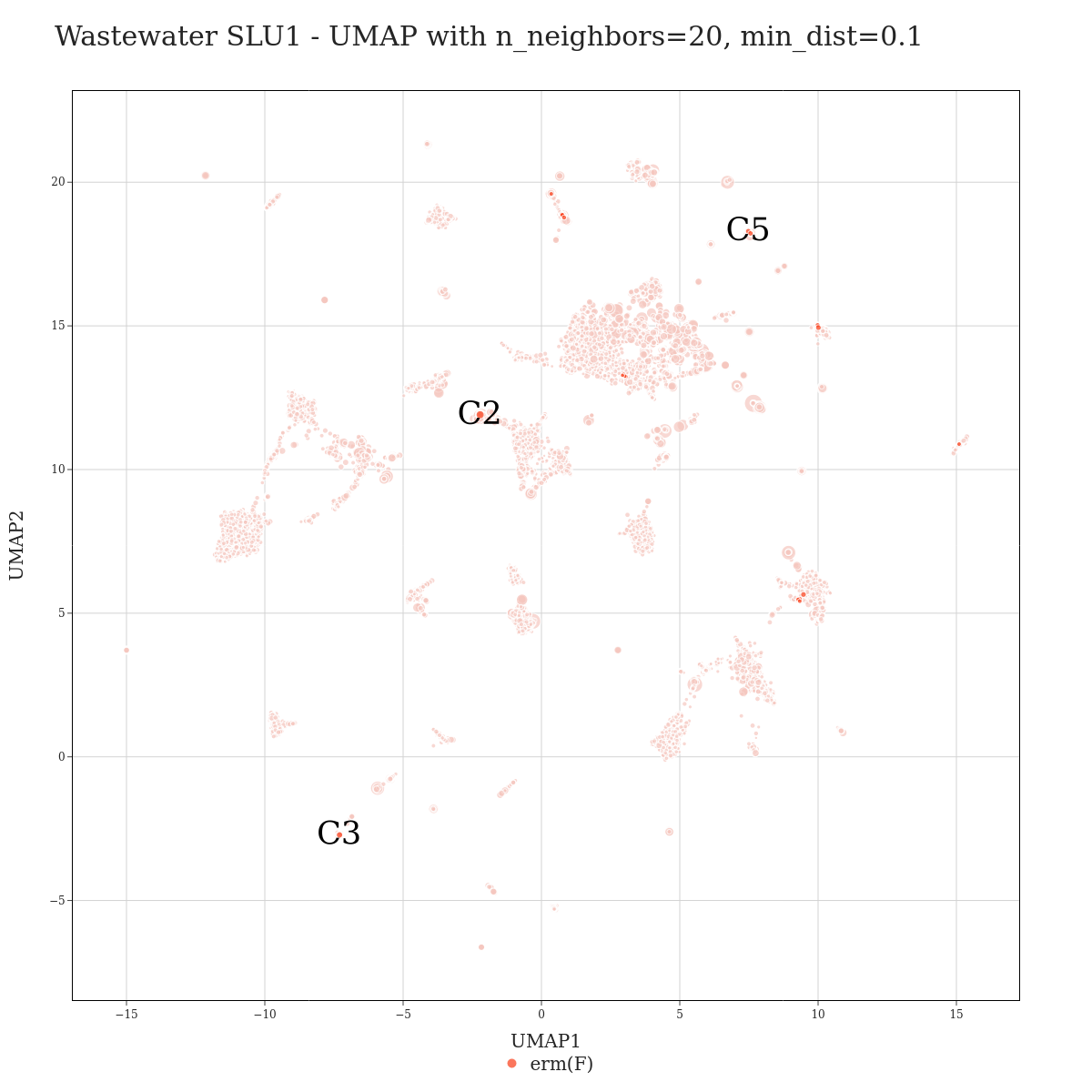

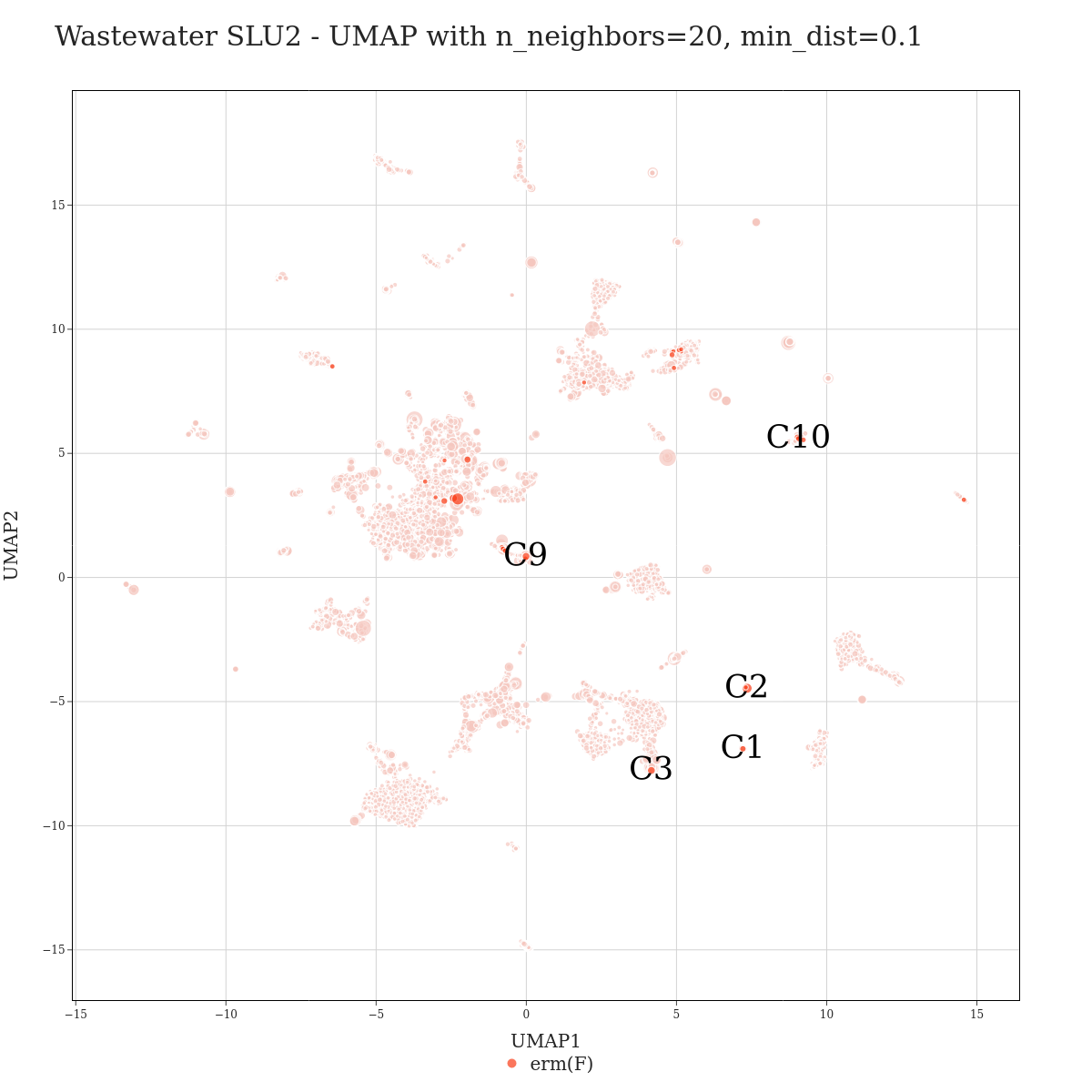

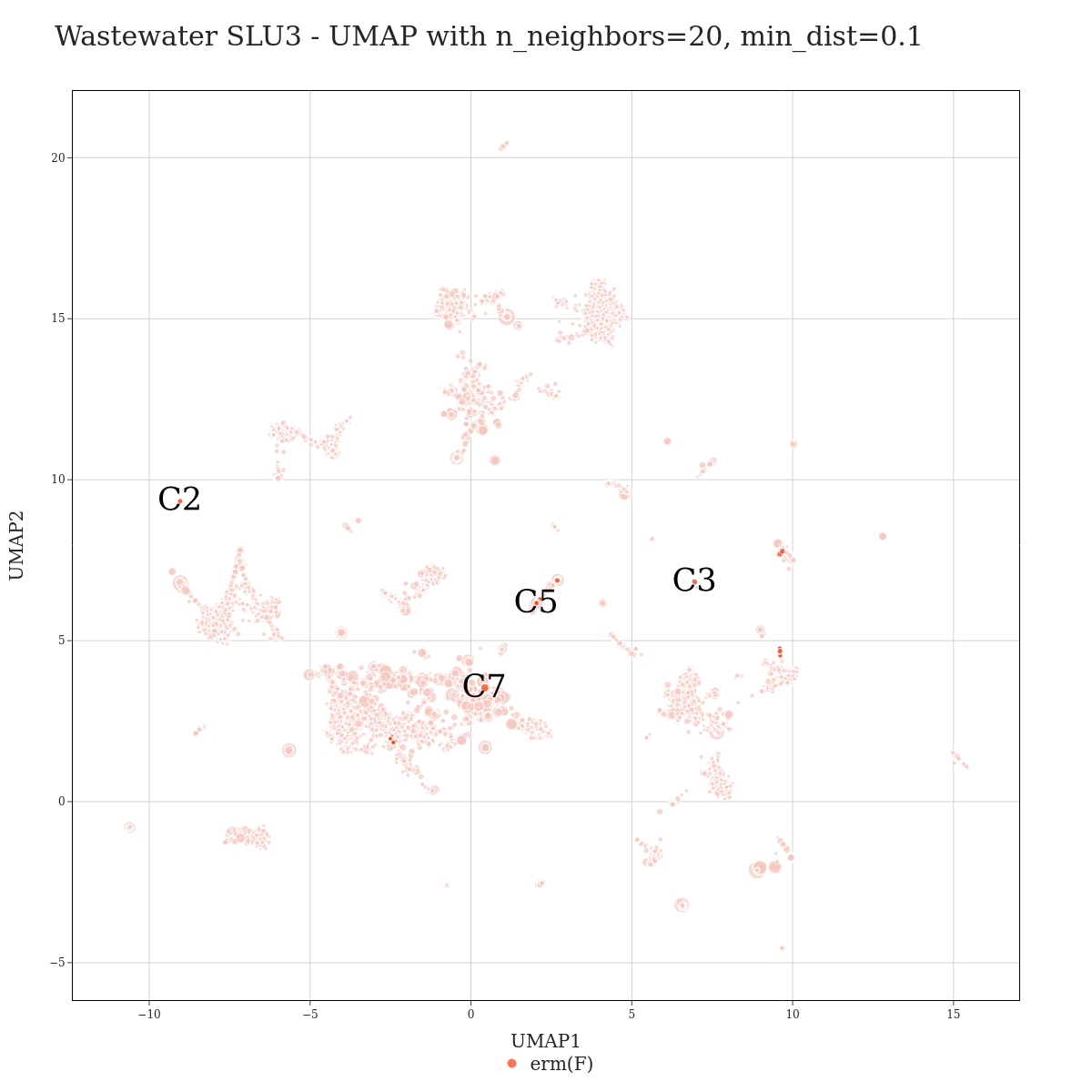

**Fig. S8.** *erm*(F) bins in different wastewater samples. The plot on the top row represents influent sample INF3, while the bottom row plots show the UMAP visualization of the metagenomic contigs of sludge samples SLU1, SLU2, and SLU3. In all plots, the pure red colored data points denote contigs with the *erm*(F) gene. The other dots (pale yellow or pink) represent non-*erm*(F) contigs. Contigs clustering together with each *erm*(F) contigs were combined into bins, and the labels of the bins are displayed on the plots.

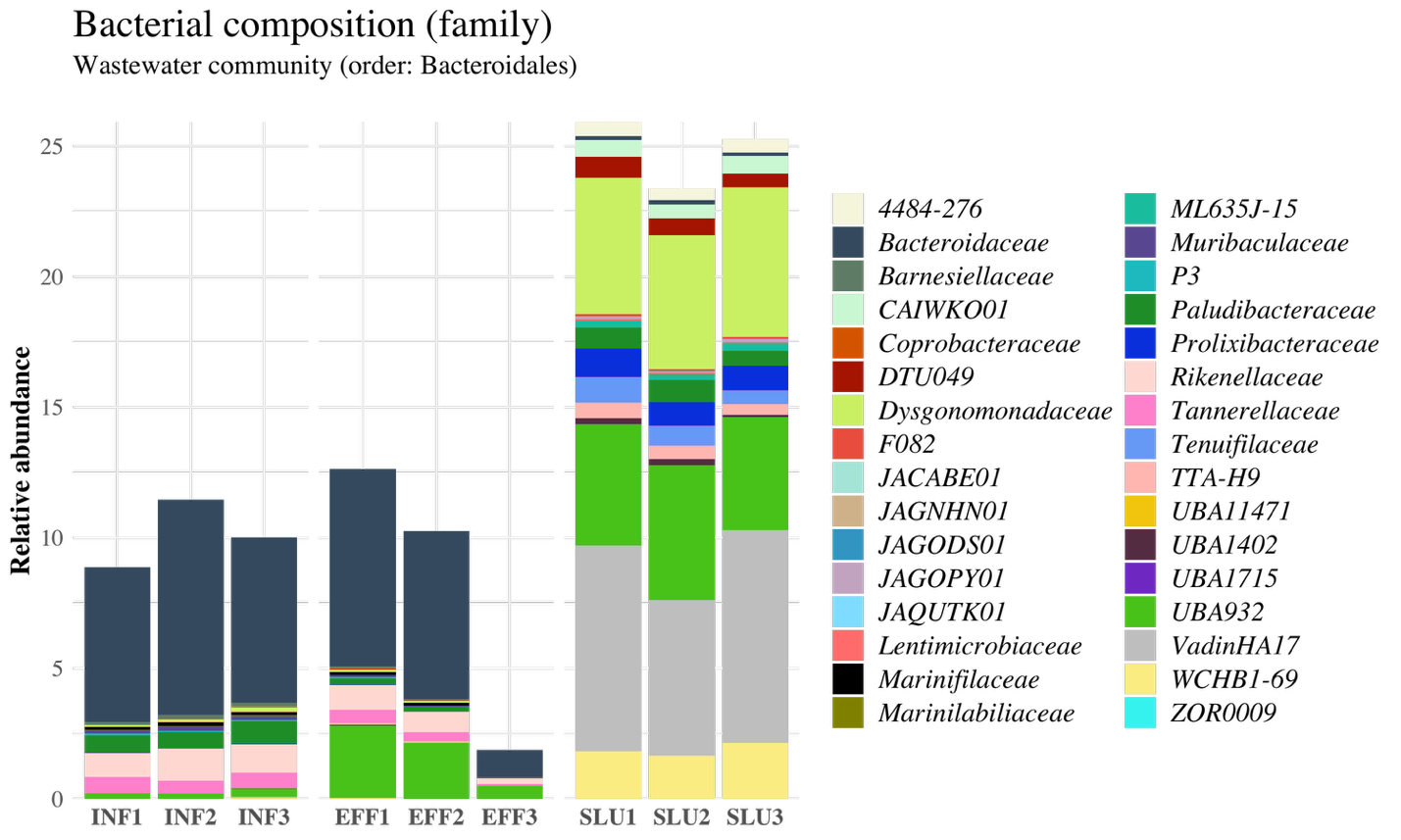

**Fig. S9.** Composition of the Bacteroidales order in wastewater communities computed with Sylph (1). Family-level taxa are shown.

**References**

1.	Shaw J, Yu YW. 2025. Rapid species-level metagenome profiling and containment estimation with sylph. Nat Biotechnol 43:1348–1359.

2.	Oksanen J, Blanchet FG, Friendly M, Kindt R, Legendre P, Mcglinn D, Minchin PR, O’hara RB, Simpson GL, Solymos P, Henry M, Stevens H, Szoecs E, Wagner H. 2020. vegan: Community Ecology Package. https://cran.r-project.org/package=vegan.
